# Reconstructing age-specific social contact patterns across 28 EU/EEA countries during the COVID-19 pandemic: linking non-pharmaceutical interventions to social mixing, 2020–2022

**DOI:** 10.64898/2026.11.09.26367661

**Authors:** Anastasia Chatzilena, Ellen Brooks-Pollock, Ewan Colman, Disa Hansson, Nikolaos Demiris, Bastian Franklin, Leon Danon

## Abstract

In response to the COVID-19 pandemic, most EU/EEA countries sequentially introduced non-pharmaceutical interventions (NPIs) to reduce social contacts and mitigate transmission. The CoMix study captured real-time changes in social mixing, but substantial spatiotemporal gaps remain, with some countries lacking pandemic contact data. We developed a model-based framework to estimate time-varying changes in age-specific contact patterns and relate these changes to NPIs recorded in the ECDC-JRC Response Measures Database. This relationship was used to predict contacts during periods without CoMix observations. For countries without CoMix data, we reconstructed temporal age-specific mixing patterns using estimates from countries with pandemic contact data selected through a country-matching process based on socio-demographic characteristics and similarity in NPI implementation. We generated age-specific contact matrices for 28 EU/EEA countries spanning 2020-2022. Reconstructed patterns captured substantial temporal and cross-country variation in social mixing and its evolving association with NPIs across diverse European contexts. Our framework provides estimates that extend incomplete pandemic contact data across both time and countries, yielding a consistent set of age-specific contact matrices for the EU/EEA. These estimates address important gaps in empirical social-contact data and provide valuable insights into the interplay of NPIs and social mixing patterns which is of critical importance to improving epidemic modelling and policy making.

## 1 Introduction

Social contact patterns are essential for characterising and predicting infectious disease transmission. However during epidemics, individuals change their behaviour, including their social contacts, in response to the risk of infection, either voluntarily or due to restrictions [1–4]. During the COVID-19 pandemic, many EU/EEA countries implemented population-wide nonpharmaceutical interventions (NPIs) to reduce transmission ranging from social distancing recommendations, closures of schools and other public places, recommendations for teleworking to large-scale lockdowns. These control strategies changed over time, with countries implementing varying action plans to reflect policy choices, social mechanisms, healthcare system capacity and transmission dynamics. In order to monitor social restrictions, the European Centre for Disease Prevention and Control (ECDC) and the Joint Research Centre (JRC) of the European Commission constructed the joint Response Measures Database (ECDC-JRC RMD) constituting an archive of NPIs used in 30 EU/EEA countries from 1 January 2020 to 30 September 2022 [5].

A key aspect of the transmission for respiratory viruses such as SARS-CoV-2 is that the infection dynamics are determined by the patterns of social mixing behaviour [6]. Since infection can be acquired by close contact, it is reasonable to assume that the intensity of transmission for each age group is proportional to the number of contacts made with another or the same age group [7]. Therefore, data on social mixing patterns is key for epidemiological modelling, yet obtaining real-time data poses a considerable challenge. Social contact data can also offer valuable insight into the effectiveness of implemented NPIs.

The impact of implemented population-wide NPIs during the COVID-19 pandemic was captured through contemporaneous social contact surveys. The CoMix contact surveys were conducted intermittently in more than 20 countries during 2020-2022 measuring the number of daily contacts between participants of different age groups [2, 4, 8]. These surveys offer a snapshot of behaviour, however, linking observed changes to NPIs is challenging because of the spatiotemporal gaps in the data, response delays and changing effectiveness.

Quantifying the changes in the social mixing patterns and their link to NPIs during the COVID-19 pandemic for most EU/EEA countries over the whole time period from 2020-2022 is of critical importance to learn from the COVID-19 pandemic and improve pandemic preparedness planning. This work uses a model-based approach to generate daily social contact patterns between 2020 and 2022 for 28 EU/EEA countries.

## 2 Methods

### 2.1 Overview of approach

We use available CoMix data for Austria, Belgium, Croatia, Denmark, Estonia, Finland, France, Greece, Hungary, Italy, Lithuania, Netherlands, Poland, Portugal, Slovakia, Slovenia and Spain to link pandemic-era social contact patterns to pre-pandemic social contact patterns through a time-varying effect model. The estimated changes in social contact patterns were then related to the implementation of NPIs, for which data are available during the whole COVID-19 pandemic period. Informed by the NPIs, out-of-sample predictions of the change in contacts were generated and consequently, the average number of contacts. To overcome the lack of CoMix surveys in certain EU/EEA countries (Czech Republic, Bulgaria, Cyprus, Ireland, Latvia, Luxembourg, Malta, Romania, Sweden, Iceland, Germany), we propose a methodology based on clustering and NPI similarity and generate the average number of contacts between age groups using the out-of-sample predictions from the countries with CoMix contact data. We implement our suggested modelling framework in Stan using the No-U-Turn sampler variant of Hamiltonian Monte Carlo.

### 2.2 Data sources

#### Pre-pandemic contact patterns – POLYMOD

The POLYMOD study is a landmark research project aimed at understanding social contact patterns and how they relate to the transmission of infectious diseases. POLYMOD is a large-scale survey conducted in 2005/2006 in eight European countries (Belgium, Germany, Finland, United Kingdom, Italy, Luxembourg, Netherlands and Poland) using a paper diary where participants documented their physical and non-physical contacts for a single day along with basic demographic information [9]. In a subsequent study, POLYMOD contact data was projected to 152 countries by synthesising multiple data sources [10]. The POLYMOD contact data have been used in many studies to incorporate the heterogeneity of social contacts that can lead to transmission [11, 12]. For our analysis, we utilized the country-specific synthetic contact matrices developed by Prem et al. [10], which refine contact patterns by incorporating nation-specific demographic and socioeconomic factors.

#### Pandemic contact patterns – CoMix

The CoMix study, conducted during the COVID-19 pandemic between 2020 and 2022, collected social contact data in more than 20 European countries through a longitudinal online survey. It started in March 2020, with survey data first being collected in the United Kingdom (UK), Belgium, and the Netherlands and extended to another 17 European countries in October 2020. The structure of the CoMix survey was based on POLYMOD [9, 10, 13], with additional questions related to group interactions, work and school attendance, household composition, use of public transportation, risk perception, and a variety of other questions. A CoMix “wave” refers to one period of survey data collection, running from the point when invitations are sent to the point when the predefined targets are reached and the survey is closed [8, 14]. The surveys in all countries were consistent in study design, making detailed comparisons between countries and inference possible. Respondents reported basic demographic information (e.g. age, gender) and were asked about their social contacts from the previous day. Characteristics of those contacts included: age of contact, gender, frequency of social interaction, whether physical contact was involved, the location where the contact took place (home/ work/ school/ transport/ leisure/ other/ house/ worship/ supermarket/ shop) and duration of the contact. The CoMix data undergo thorough cleaning and validation procedures following a specific data management protocol and are available through a public Zenodo-based repository (www.socialcontactdata.org/data).

#### Non-pharmaceutical interventions - ECDC-JRC Response Measure Database (RMD)

This database was initiated to collect information on non-pharmaceutical interventions across all EU/EEA countries [5] from 1 January 2020 to 30 September 2022. It is organised in a hierarchical way, with information at three levels of granularity. The coarsest level (Level 1) contains information on measures concerning case management and quarantine, general measures (including communications), hygiene and safety, domestic travel, international travel and physical distancing. NPIs are further categorised by two additional levels (Levels 2 and 3 with 41 and 51 categories, respectively). In our analysis, we used Level 2 categories under physical distancing that might have a direct effect on the expected change in contacts. Visualisations of ECDC-JRC response measures data for 30 countries along with CoMix mean contacts and COVID-19 related deaths [15] where available are presented in Supplementary Material. Based on the intensity of implementation (none, partial, full) we formed categorical variables that can take on 3 values. Level 2 NPIs indicate closure of educational institutions (CEI), workplace measures (W), stay at home orders (SH), public gathering restrictions (PbG), private gathering restrictions (PrG), closure of public spaces (CPS) and measures for special populations (SpP).

Table 1 categorises countries according to the availability of social contact data: 17 countries had some social contact data covering the COVID-19 pandemic period, together with data related to pre-pandemic. A further 11 countries had some form of social contact data related to pre-pandemic and 2 countries had neither pre-pandemic contact data nor CoMix data available in the datasets used. All countries are included in the ECDC–JRC RMD.

**Table 1:** Countries in the EU/EEA categorised by social contact data availability.

| **Category** | **Countries** |
|---|---|
| Countries with POLYMOD and CoMix sampled <Oct 2020 to >Oct 2021 | Belgium, Netherlands |
| Countries with POLYMOD and CoMix sampled between Oct 2020 and Oct 2021 | Finland, Italy, Poland |
| Countries with synthetic POLYMOD and CoMix sampled between Oct 2020 and Oct 2021 | Austria, Croatia, Denmark, Estonia, France, Greece, Hungary, Lithuania, Portugal, Slovakia, Slovenia, Spain |
| Countries with synthetic POLYMOD but without CoMix | Bulgaria, Cyprus, Czech Republic, Iceland, Ireland, Latvia, Luxembourg, Malta, Romania, Sweden |
| Countries with POLYMOD but without CoMix | Germany<sup>a</sup> |
| Countries without synthetic POLYMOD or CoMix | Norway, Liechtenstein |

<sup>a</sup> Alternative social contact data are available from the COVIMOD survey [16].

### 2.3 Construction of age-specific CoMix contact matrices

CoMix contact survey data were used to construct age-specific mixing matrices across six modelling age groups: 0–9, 10–19, 20–24, 25–44, 45–64, and 65+ years of age. The CoMix questionnaire records the ages of participants and their reported contacts in predefined age bands (e.g. <1, 1–4, 5–9, 10–14, 15–19, 20–24, 25–34, 35–44, 45–54, 55–64, 65–69, 70–74, 75–79, 80–84, 85+ years of age). To map these finer questionnaire bands to our six broader modelling age groups while preserving the underlying population age structure, imputation was used to allocate each participant, or contact of a participant, to one of these age groups. Specifically, an exact age was allocated by choosing randomly from the range provided in the data, with probability weighted by the population age distribution according to World Population Prospect estimates for 2020 [17]. Participant and contact ages were then regrouped into the six categories listed above.

A weight was applied to contacts to adjust for the bias that arises from different levels of participation on weekends (compared to weekdays) when contact behaviour is expected to also be different. The weighting factor was calculated such that the number of participants providing data for a weekend day, when multiplied by the weekend weighting factor, equals 2/7 of the total weight of participants. A similar calculation is made for the weekday weighting factor such that 5/7 of the weight of participants comes from those who participated on a weekday. A further adjustment is applied to ensure symmetry in the amount of contact between age groups. For example, the total number of contacts between groups *a* and *b* (calculated as the number of individuals in the population in group *a* multiplied by the mean number of contacts they have with individuals in *b*) is the same as the number of contacts between groups *b* and *a*. To ensure this is the case we use the population weighted average used in [18].

The timing and age coverage of CoMix data varied considerably across countries (Figure 1). Most data were collected for adults, and even though there were survey waves collecting children’s data, asking parents to answer on behalf of their child, the survey timings and frequencies were different in most countries. Although adult contact data were collected consistently throughout the study period, children’s contact data were collected during fewer waves and at different time points. This created a methodological challenge when constructing complete age-specific contact matrices. To address this data structure limitation, first, for each country, we identified the available children’s survey waves and their corresponding collection dates. Due to the limited availability of children’s data, we assumed that children’s contact patterns remained constant between consecutive children’s data collection points. Then, for each adult survey wave, we matched it with the most recent valid children’s data available at that time point. If no prior children’s data existed for a country, we used the first available children’s wave data retrospectively. Using the matched data, we constructed complete age-specific contact matrices for each adult survey wave.

**Figure 1:**
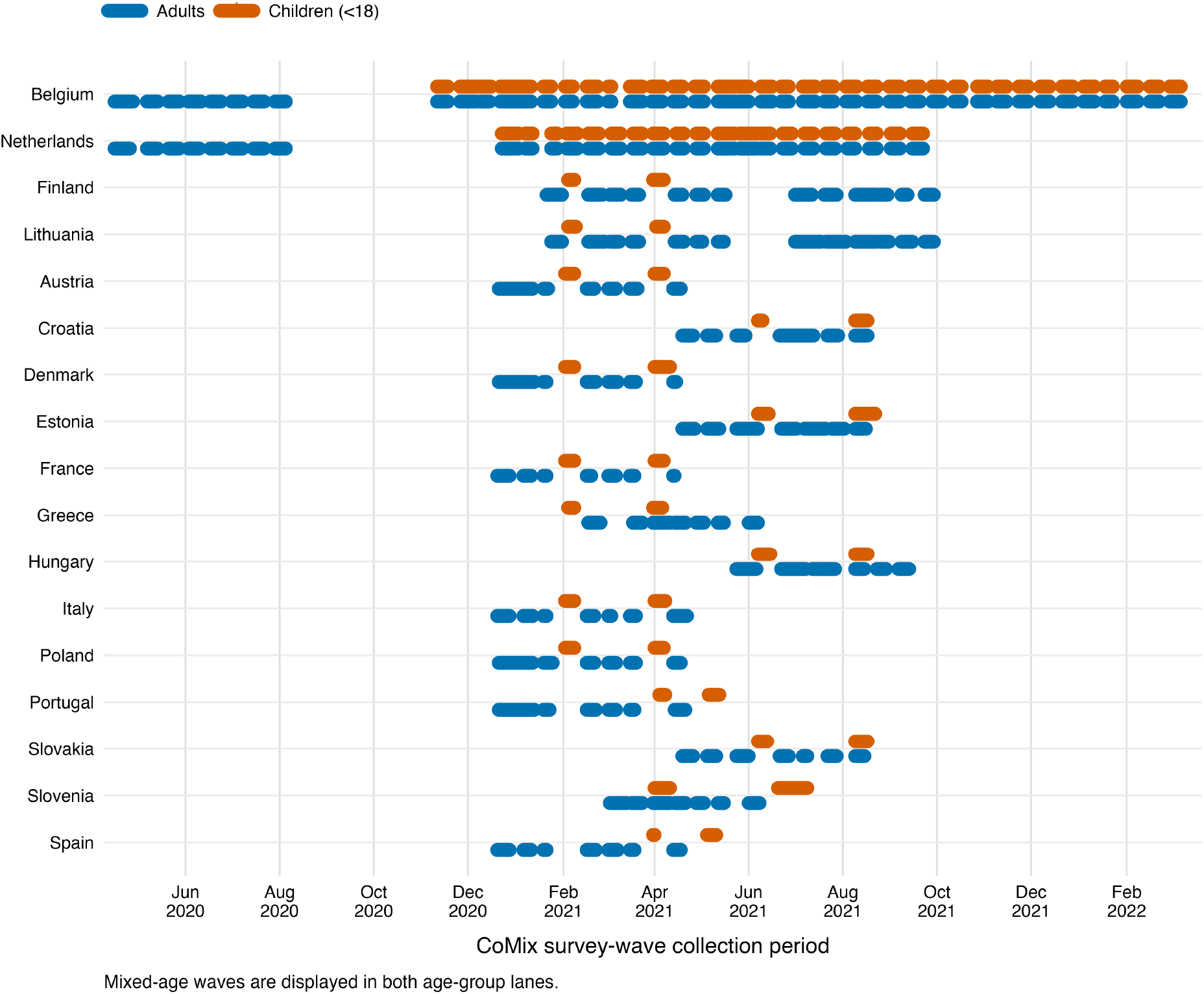
Timing of CoMix survey waves by country and age group. Horizontal segments indicate the start and end dates of each survey wave. Adult and child participants (<18 years) are displayed in separate vertically offset lanes within each country.

### 2.4 Modelling framework

Our modelling approach is structured in two interconnected levels. At the first level, we relate pre-pandemic contact patterns to pandemic contact patterns through a time-varying model, estimating how mixing in different settings evolves over time. At the second level, we link these estimated time-varying changes to non-pharmaceutical interventions (NPIs), which allows us to generate predictions for countries or periods without direct survey data. Together, these levels form a hierarchical framework in which information flows from observed survey data to NPI-informed predictions. A schematic link between the directed acyclic graphs of the two levels of the modelling framework is presented in Figure 2.

**Figure 2:**
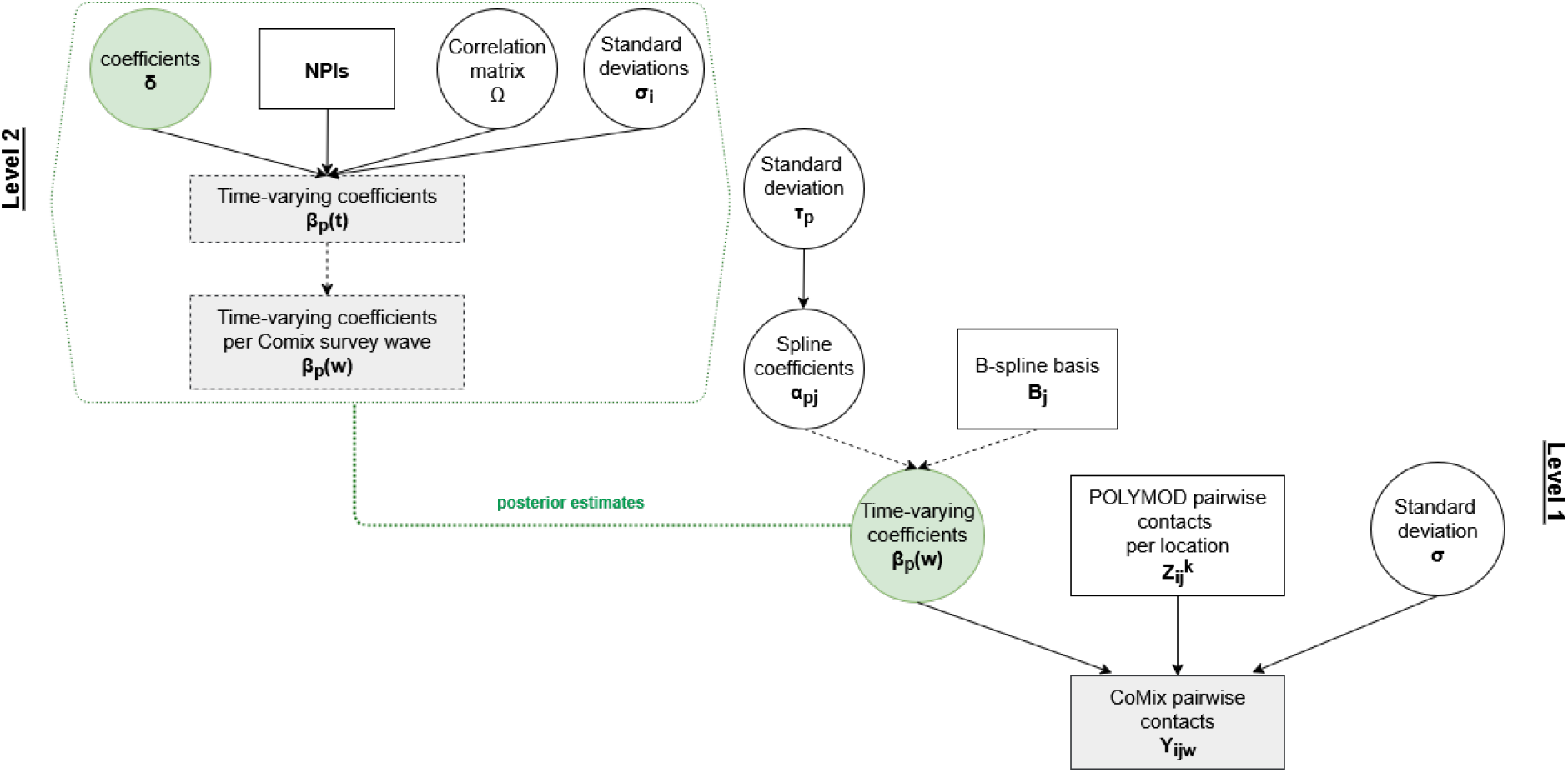
Schematic representation of the two-level modelling framework. In each level, square nodes denote observable quantities and black circles denote latent quantities. In Level 2, grey square nodes correspond to posterior estimates of latent quantities obtained from Level 1. Solid arrows represent stochastic dependencies, while dashed arrows represent deterministic dependencies.

#### 2.4.1 Relate pre-pandemic contact patterns to pandemic contact patterns for EU/EEA countries with data for both time periods

We developed a Bayesian approach based on the model proposed by Lajot et al. [19] to estimate the relationship between pre-pandemic contact patterns (synthetic POLYMOD) and pandemic contact patterns (CoMix) for EU/EEA countries with data for both time periods.

We employed a time-varying effect model (TVEM) [20] where the pandemic contacts are described as a linear combination of the pre-pandemic contacts. The outcome variable *Y*_*ijw*_ is the average number of contacts for individuals in age group *j* with individuals of age group *i* at time *w*, where *w* denotes the adult waves of the CoMix survey, with children’s contact data treated as constant between collection points and matched to these adult waves. The independent variables 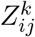 denote the pre-pandemic average number of contacts for individuals in age group *j* with individuals of age group *i* at location *k* where *k* corresponds to home, work, school and other places. We considered 6 age groups [0,10), [10,20), [20,25), [25,45), [45,65) and 65+ years. Then, for each country *Y*_*ijw*_ can be described as:

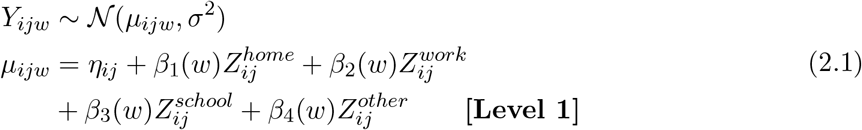

where *η*_*ij*_ is a fixed, pair-specific term, *β*_*p*_(*w*) (*p* = 1, 2, 3, 4) are smooth coefficient functions of time *w* and *σ*^2^ is the variance parameter.

The intercept *η*_*ij*_ is a fixed, pair-specific term representing the time-constant component of pandemic contacts between age groups that cannot be attributed to any linear combination of the four pre-pandemic POLYMOD location matrices. It is precomputed from the data as the timeaverage of the per-wave residuals obtained after projecting the observed CoMix contact matrix onto the subspace spanned by the four POLYMOD location-specific matrices. Epidemiologically, *η*_*ij*_ captures a systematic structural shift in mixing between age groups that persisted throughout the pandemic period and is not representable as a rescaling of pre-pandemic contact patterns across any single setting. This could reflect genuine changes in mixing behaviour during the pandemic, but may also partly reflect differences in sampling design and population coverage between the CoMix and POLYMOD surveys.

The *β*_*p*_(*w*) quantify the time-varying sensitivity of pandemic contacts to pre-pandemic contacts at location *p*, i.e. the expected change in *E*[*Y*_*ijw*_] per unit increase in 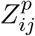 conditional on other locations, after removing the systematic structural shift.

Spline-based methods represent the most commonly used approach in varying-coefficient models. We utilised B-spline basis functions since they can form sufficiently complex models to reveal nuanced changes. The number of knots and the knot positions are tuning parameters which may have an important impact on the estimated shape of a spline function. Thus, we approximate the coefficient functions via:

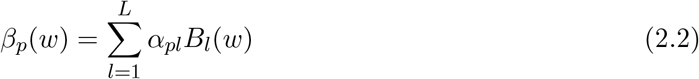

where *L* is the number of B-spline basis functions, *α*_*pl*_ denote the coefficients associated with each basis function and *B*_*l*_(*w*) is a B-spline basis function evaluated at wave index *w*.

A large number of knots result in high flexibility but may also result in overfitting while a small number of knots may result in an “oversmooth” estimate and underfitting. Employing a Bayesian approach can alleviate the problems associated with the choice of number/locations of the knots [21]. For any given set of knots, and any B-spline order,∑ _*l*_ *B*_*l*_(*w*) = 1. Thus, if the B-spline coefficients *α*_*pl*_ are all equal, then the resulting spline *β*_*p*_(*w*) is a constant function, and if the coefficients of nearby B-splines are close to each other, we will have less local variability. By employing a random-walk prior on the coefficients *α*_*pl*_, we reduce the risk of a prior choice of a large number of knots resulting in overfitting. Therefore, we used a random-walk prior as follows:

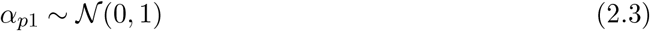

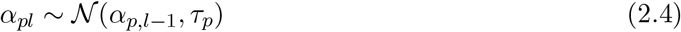

We estimated the model by choosing a priori 10 knots for all coefficient functions and placed the knots at equally spaced quantiles of the wave index *w* in each country. We conducted a sensitivity analysis examining various prior choices for the number of knots, including extreme cases, which confirmed that model fit remained robust across different knot configurations with minimal impact on smoothing quality. To complete the model specification, we considered a half-Cauchy prior for the standard deviation *σ, σ ∼ C*^+^(0, 5). Finally, a half-Cauchy prior was assigned for the standard deviation of the random-walk *τ*_*p*_ *∼ C*^+^(0, 5) for *p* = 1, 2, 3, 4.

Half-Cauchy priors were chosen for the scale parameters (*σ* and *τ*_*p*_) as weakly informative priors with positive support and heavy tails, providing mild regularisation while allowing large values when supported by the data. We fitted our model on country level in Stan using the No-U-Turn Sampler (NUTS) for all countries with CoMix data.

#### 2.4.2 Fill in gaps in countries with pandemic data with incomplete time coverage by relating changes in social contact patterns to non-pharmaceutical interventions

The estimated coefficients *β*_*p*_(*w*) (*p* = 1, 2, 3, 4) from Level 1 express the time-varying expected change in the contacts between two age groups during the pandemic, resulting from a change of one contact between those age groups at different locations before the pandemic. This expected change in contacts can be linked to the implemented NPIs in each country during the different time periods using the ECDC–JRC RMD [5], forming the basis of Level 2 of our modelling approach (Figure 2). By establishing a mapping between posterior percentiles of the time-varying change in contacts between two age groups and NPIs we can then generate the average number of contacts between age groups during the pandemic in sampling points where there are available measurements of NPIs, but where contact data from the CoMix surveys are not available.

The large set of categorical variables constructed from ECDC–JRC RMD reported NPIs related to physical distancing measures are highly correlated. Since the effects of sequentially introduced NPIs are highly interdependent, making it hard to disentangle their individual contribution, we implemented Multiple correspondence analysis (MCA) to identify systematic patterns of variations within the data and reduce their dimension. The factor dimensions of the MCA represent linear combinations of the original categorical variables [22]. These dimensions capture the main patterns and associations in the categorical NPI variables. In most countries, the first five dimensions explain at least 60% of the variability of the data, with the first dimension mostly consisting of measures related to closure of public spaces and educational institutions. For each CoMix country, we used the first 5 factor dimensions of the MCA on all the NPIs under the physical distancing category as summaries of these NPIs. Therefore, for each country and time point *t*, we denote by Dim_*t*_ = (Dim_*t*1_, …, Dim_*t*5_) the first five MCA factor dimensions which are used as time-varying predictors in Level 2 of our modelling approach.

The posterior estimates of the change in contacts between two age groups from Level 1 were obtained for each CoMix survey wave while data on the NPIs were collected daily. In order to translate the posterior percentiles of the coefficients *β*_*p*_(*w*) for each CoMix wave *w* into coefficients *β*_*p*_(*t*) for each day *t*, we linearly interpolated each of the four coefficient functions *β*_*p*_(*·*) independently for each location p between waves, assuming that changes to contact patterns occur gradually.

By definition, the expected change in contacts as expressed by the estimated *β*_*p*_(*t*) is not independent among the different locations, therefore, we will naturally assume that there is a cross-equation error correlation among locations. So, we run the following multivariate regression for each country:

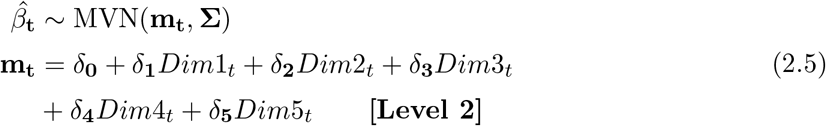

Where 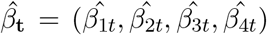. The vectors of regression coefficients are denoted by *δ*_**q**_, *q* = 0, 1, 2, 3, 4, 5, and the covariance matrix is represented by **σ**. We can rewrite **σ** in terms of the correlation matrix Ω as, **σ** = *D*_*σ*_Ω*D*_*σ*_, where *D*_*σ*_ denotes a diagonal matrix with diagonal elements *σ*_*i*_, *i* = 1, 2, 3, 4. We specified an LKJ correlation matrix distribution [23] for Ω. The parameterization of LKJ distribution used in Stan allows to sample matrices depending on a shape parameter. The shape parameter determines whether we expect to sample posterior matrices close to the identity matrix or to general positive definite matrices [24]. In principle, large values of the shape parameter pertain to correlations close to zero, while values less than 1 suggest high probability for non-zero correlations. We use Stan’s implicit parameterization of the LKJ correlation matrix in terms of its Cholesky factorisation. We account for the dependence in our variables by assuming that Ω *∼* LkjCholesky(0.5) which translates to a U-shaped prior over random correlation matrices and assign a half-Cauchy prior on the standard deviations: *σ*_*i*_ *∼ C*^+^(0, 5). For the regression coefficients, we use Normal priors, *δ*_*q*_ *∼ N* (0, 1).

To generate predictions for periods without CoMix observations, we make the strong simplifying assumption that, within each country, the relationship between the recorded NPIs, as represented by the MCA dimensions, and changes in contacts remains stable over time. This assumption may be restrictive because behavioural responses, adherence to measures, and the context in which interventions were implemented may have changed during the pandemic. Under this assumption we can use the estimated regression coefficients, *δ*_*q*_ to generate out-of-sample predictions of the change in the number of contacts between age groups *β*_**t**_ by sampling from a multivariate normal with mean 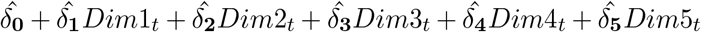 and covariance matrix 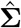.

By generating *β*_**t**_ in sampling points where there are available measurements of NPIs, but where contact data from the CoMix surveys are not available, we can indirectly generate the number of contacts between age groups as a linear combination of the pre-pandemic contacts in different locations (home, work, school and other places) weighted by out-of-sample predicted vector *β*_**t**_.

#### 2.4.3 Infer pandemic contact patterns for EU/EEA countries with pre-pandemic data (but not pandemic data)

To construct social contact data for countries lacking CoMix surveys, we relied on our estimates for the countries with CoMix data. For each target country without pandemic data (non-CoMix country), we assigned one matched country with pandemic data (CoMix country)from which the estimated relationship between NPIs and changes in contacts could be transferred..

To select a matched CoMix country for each target country without CoMix data, we implemented a multi-faceted matching approach. We began by performing hierarchical clustering of all countries based on their NPI implementation patterns using the first 5 factor dimensions from the MCA analysis. Multiple hierarchical clustering methods were employed to identify consistent groupings across different algorithmic approaches (see Supplementary Material). Simultaneously, for each target country without pandemic data, we quantified similarity with potential CoMix country matches using Euclidean distances between their time-varying NPI factor dimensions. This distance-based matching provided a ranked list of potential neighboring countries based on similarity in pandemic response measures (Supplementary Material). To further assess the plausibility of these potential matches, we cross-referenced them with the socio-demographic clusters identified by Prem et al. [10], which grouped countries based on fundamental characteristics including GDP per capita, total fertility rate (TFR), population density, population growth rate, under-five mortality rate (U5MR), health expenditure per capita, life expectancy of males, Internet penetration rate, and urbanization rate. The final matched CoMix country was selected primarily on the basis of NPI similarity, with the clustering results and socio-demographic groupings used as supporting information to assess the plausibility of the selected match and provide a basis for extrapolating social contact patterns.

Importantly, NPI data from the target non-CoMix countries were not used to estimate the relationship between NPIs and changes in contact patterns. This relationship was estimated separately for each CoMix country using its observed contact data and corresponding NPI factor dimensions. NPI data from the target countries were used only to inform the country-matching procedure and, subsequently, as predictors when applying the fitted Level 2 model from the selected matched CoMix country.

For prediction, we assumed that the relationship between NPIs and changes in contact patterns estimated for the matched CoMix country could be transferred to the target country. The target country’s implemented NPIs under the physical distancing category, summarised by their first five MCA factor dimensions from the ECDC-JRC RMD NPI indicators, were then used to generate its predicted time-varying contact coefficients.

Let *c* denote the selected matched CoMix country for a target non-CoMix country *nc*. For each day *t*, we generated the location-specific time-varying coefficient vector for the non-CoMix country, 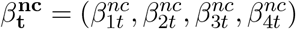, by sampling from multivariate normal with mean 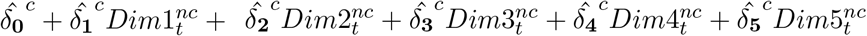 and covariance matrix 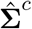 where 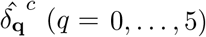 are the estimated marginal effects of the MCA dimensions from country *c* in Level 2, 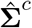 is the corresponding estimated covariance matrix and 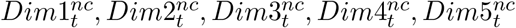 represent the first five factor dimensions of an MCA on all the NPIs under the physical distancing category for the target country *nc* at time *t*.

Given the generated coefficient functions 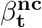, we obtained pandemic contacts for the nonCoMix country by combining these with the pre-pandemic contact matrices as follows:

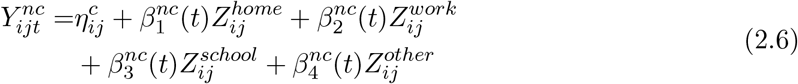

where 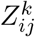 denotes the pre-pandemic average number of contacts for individuals in age group *j* with individuals of age group *i* at location *k*, where *k* corresponds to home, work, school and other places.

The intercept 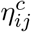 is borrowed from the matched CoMix country *c*, as no CoMix data are available for the target country *nc* from which to compute country-specific projection residuals.

## 3 Results

### Complete social mixing patterns for countries with some available pandemic data: Austria, Belgium, Croatia, Denmark, Estonia, Finland, France, Greece, Hungary, Italy, Lithuania, Netherlands, Poland, Portugal, Slovakia, Slovenia and Spain

We estimated time-varying coefficient functions for 17 EU/EEA countries with CoMix survey data spanning most or part of the pandemic period (2020-2022). The estimated coefficients (Level 1) quantify how the contribution of pre-pandemic contacts in each location (home, work, school, other) changes over pandemic waves, after removing systematic structural shifts. Across countries, several consistent patterns emerge. School-related contacts show pronounced cyclical behaviour consistent with school closures and holiday periods, workplace coefficients tend to decline and remain suppressed in many settings consistent with sustained teleworking measures, and “other places” coefficients exhibit substantial uncertainty, likely reflecting rapid policy changes and heterogeneous adherence. Home coefficients are generally stable or increasing, particularly in countries with stronger stay-at-home measures. Supplementary Figures S2 and S4 show the estimated coefficients and their NPI-predicted trajectories for all countries with CoMix data.

To translate the estimated time-varying coefficients into predicted contact trajectories for countries with CoMix data, we combined the Level 1 and Level 2 models. This yielded reconstructed average numbers of contacts between age groups over time. To illustrate these results across different European regions, Figure 3 presents four countries, Belgium (Western), Poland (Eastern), Italy (Southern), and Finland (Northern). Results for all 17 countries with CoMix data are provided in the Supplementary Figure S5.The reported (dots) average number of age-specific contacts over time are plotted along with the predicted (lines and shaded areas), sampled based on estimates of Level 2. The lines and the dark-shaded areas represent the predicted median and 95% Credible Intervals (CIs) resulting from using the posterior medians of the TVEM coefficients to implement Level 2 and then reconstruct the contacts. The light-shaded areas depict the CIs range of the predicted contacts resulting from using the 2.5% and 97.5% posterior percentiles of the TVEM coefficients to implement Level 2.

**Figure 3:**
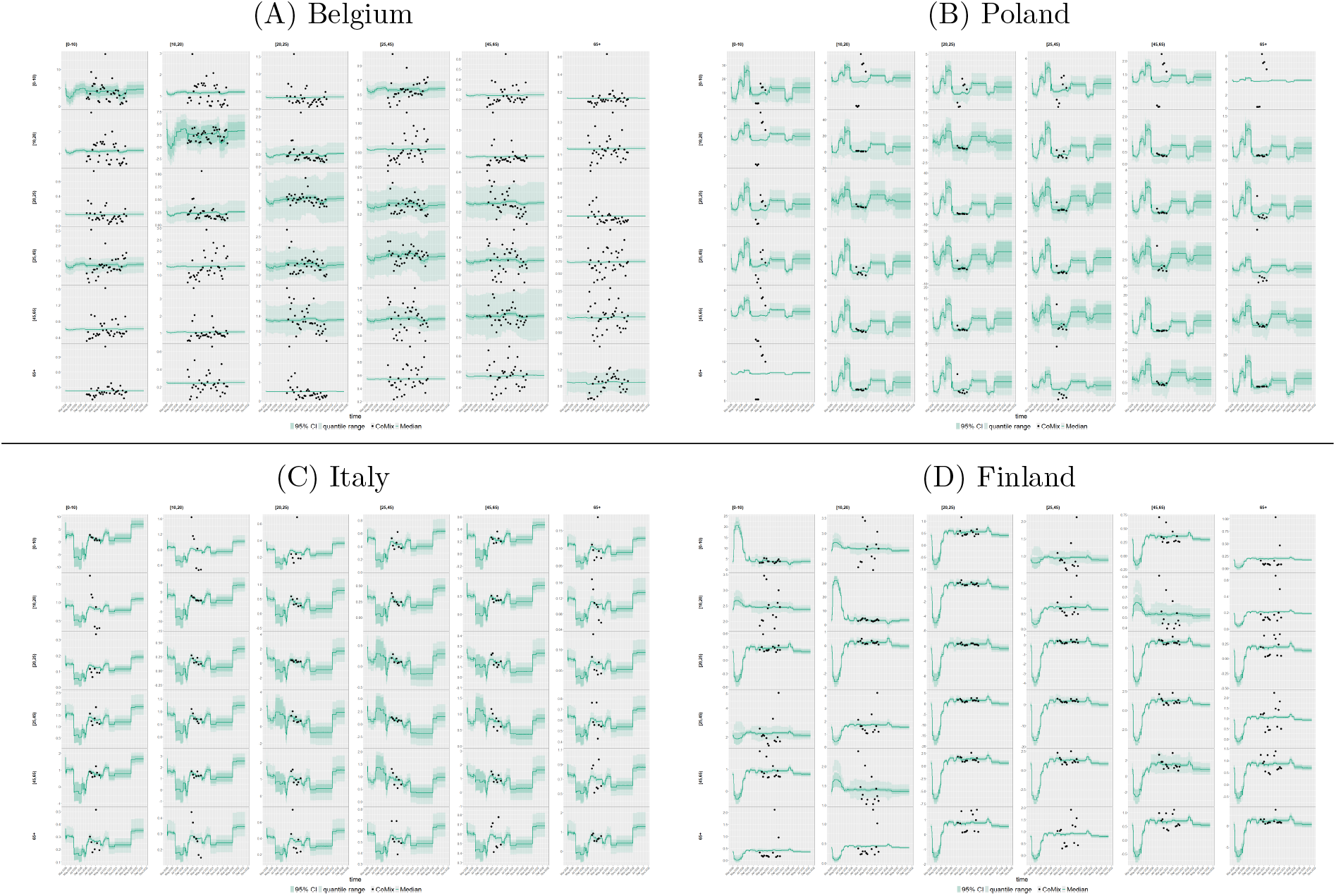
Predicted number of contacts between age groups over time for countries with CoMix survey data, medians (lines) and 95% CI (dark shaded areas) based on posterior medians of *β*_**t**_ along with reported number of contacts from CoMix(dots).The CIs range (light shaded areas) of the predicted contacts resulting from using the 2.5% and 97.5% posterior percentiles of *β*_**t**_ are also presented.

The relationship between contact trajectories and NPI implementation varies across countries (Figure 3, Supplementary Figure S1). Two patterns are common across countries. First, reductions in child-related contacts tend to coincide with the timing of educational measures, although this correspondence may also partly reflect school-calendar and holiday effects, which the modelling framework does not separately identify. Second, a portion of the contact variation is not explained by the recorded NPIs and might be consistent with voluntary behavioural change, differing levels of compliance, seasonality, or other unmeasured time-varying factors. In several countries the resulting seasonal and cyclical variation is comparable in magnitude to the changes associated with policy, indicating that periodic and behavioural factors operate alongside formal interventions.

In Belgium, the predicted age-specific contacts show partial correspondence with the responsemeasure timeline. The clearest changes are observed in age pairs involving children, particularly child–child contacts, where reductions and subsequent rebounds broadly match periods of educational closures and reopening. Contacts among adult age groups change relatively little over time, despite the presence of teleworking measures, gathering restrictions, and closures of restaurants, entertainment venues and public spaces. This suggests that for Belgium, the recorded NPIs capture some important changes in contact behaviour, especially for school-related mixing, but do not fully explain the temporal variation in adult contacts (Figure 3A, Supplementary Figure S1(b)). Poland exhibits significant wave-to-wave variability in predicted contacts, and some sharp changes appear difficult to attribute to the recorded response measures alone, implying that factors beyond policy like behavioural changes, compliance, or other unmeasured timevarying factors may substantially influence contact patterns (Figure 3B, Supplementary Figure S1(x)). In Italy, the largest decreases are observed in child–child contacts, and these broadly align with periods when educational measures were in place, especially the school-closure phases in spring 2020 and again from late 2020 into early 2021. This suggests that the response-measure timeline is translated most clearly into school-related mixing. Contacts among working-age adults also decrease during periods of teleworking and broader restrictions, but because these overlap with stay-at-home orders and closures of public venues, the reductions are better interpreted as the combined effect of multiple measures (Figure 3C, Supplementary Figure S1(p)). Finland maintains comparatively stable predicted contact levels and smaller swings across many age pairs, consistent with a less stringent and more recommendation-based policy approach. Early in the pandemic, school-related measures coincide with reductions in children’s contacts but later variation in child-related panels is less clearly linked to recorded educational measures and may instead reflect school-calendar effects or voluntary behavioural changes. Adult contacts do not show a sustained decrease, consistent with the absence of prolonged workplace restrictions in the response-measure timeline (Figure 3D, Supplementary Figure S1(i)).

Our modelling framework assumes a Gaussian likelihood for the mean number of contacts between age groups, which facilitates a linear, interpretable relationship between pre-pandemic and pandemic contacts. A direct consequence is that the posterior predictive distribution is not constrained to be non-negative, so for combinations of age pairs and time points where the inferred contact intensity is very low, the model can occasionally yield small negative predicted values. These negative predictions are not statistically distinguishable from zero and should be interpreted as an artefact of the Normal approximation, indicating contact levels that are effectively zero rather than substantively meaningful negative contacts.

### Reconstructed social mixing patterns for countries without pandemic social contact data: Czech Republic, Bulgaria, Cyprus, Ireland, Latvia, Luxembourg, Malta, Romania, Sweden, Iceland, Germany

Constructed age-specific contacts for countries without CoMix data are illustrated in Figure 4 for four countries selected to provide examples from different European regions: Luxembourg (Western), Romania (Eastern), Cyprus (Southern), and Sweden (Northern). Reconstructed contacts for all EU/EEA countries without CoMix data but with pre-pandemic contact data are provided in Supplementary Figure S8. Each country was matched to a CoMix country based on clustering of socio-demographic characteristics according to Prem et al. [10] and similarity of NPI implementation as represented by their first five factor dimensions. The full matching of all countries without pandemic social contact data to their selected CoMix counterparts is reported in Supplementary Figure S7.

**Figure 4:**
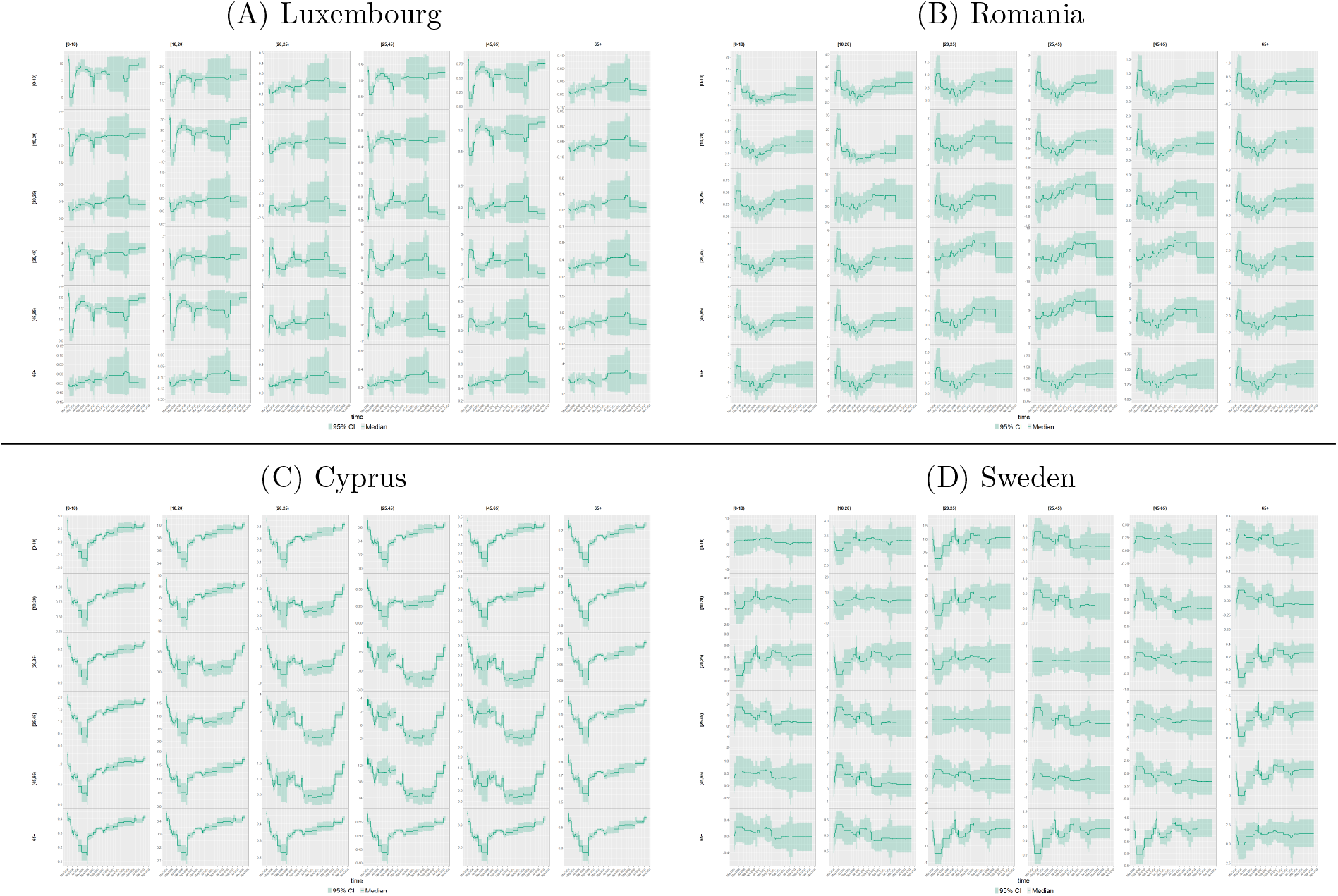
Reconstructed average number of contacts between age groups over time for countries without CoMix survey data, medians (lines) and 95% CI (dark shaded areas). Contact patterns were reconstructed using the NPI-informed predictions from the matched CoMix country.

Because the reconstructed contact patterns for countries without CoMix data are conditional on their observed NPI trajectories, correspondence between reconstructed contacts and the timing of NPIs should be interpreted as a feature of the model-based reconstruction rather than as independent evidence of a causal effect of those interventions.

In Luxembourg, our contact reconstruction approach estimates sharp reductions in child–child and child–adult contacts that coincide with the spring 2020 school closures and national lockdown, while adult–adult contacts are estimated to show a relative increase during this period, coinciding with the absence of workplace closure mandates and the persistence of adult interactions. A similar pattern is estimated in early 2021, coinciding with renewed restrictions and school closures that reduced child-related contacts but left adult mixing largely unchanged. In Romania, the reconstructed contacts show broad reductions after the early pandemic period, with the clearest changes occurring in age pairs involving children. These estimated reductions broadly align with prolonged educational closures and restrictions, although later fluctuations may also reflect school-calendar effects. Estimated contacts among working-age and older adults show less direct correspondence with workplace-related measures. Several adult–adult panels increase around summer 2021 despite continued prolonged restrictions, reflecting possible additional influences such as behavioural adaptation, compliance, seasonality, or unmeasured contextual factors. In Cyprus, the reconstructed contacts fall substantially during 2020, followed by gradual increases during 2021 and 2022. Contacts involving children are estimated to decline during the early phase when educational closures were in place, while during parts of 2021 when educational measures are no longer clearly in place, there is an expected increase suggesting that the rebound is broadly consistent with the removal of school-related restrictions, although school-calendar effects and broader behavioural changes may also contribute. Estimated adult–adult contacts also decline during periods with teleworking, stay-at-home orders, gathering restrictions and closures of public venues. In Sweden, where stringent stay-at-home orders were not implemented, reconstructed contacts remain relatively stable throughout the study period. Educational measures are limited, and the predicted child-related contacts do not show a strong sustained decrease aligned with these measures. Reconstructed contacts among working-age adults show some reductions during periods with workplace adaptation and gathering restrictions. Notably, contacts involving older adults decrease at the beginning of the pandemic, which is consistent with early visiting restrictions and measures for special populations. Estimated contacts involving the 65+ years of age group show temporal variation over the study period, potentially reflecting voluntary protective behaviour among older adults and those interacting with them, rather than a clear response to a specific formal restriction. In general, adult–adult contacts are estimated to remain comparatively stable at higher levels, reflecting Sweden’s reliance on voluntary measures rather than mandatory restrictions.

As described above for the countries with CoMix data, the Gaussian likelihood occasionally yields small negative posterior predictions at age–pair–time combinations with very low inferred contact intensity and these should again be interpreted as effectively zero rather than as meaningful negative contacts.

Constructed contacts for all the EU/EEA countries without CoMix data but with pre-pandemic data are also included in the Supplementary Material.

## 4 Conclusions

In this study, we developed a comprehensive modeling approach for reconstructing age-specific social contact patterns during the COVID-19 pandemic across 28 EU/EEA countries. By combining pre-pandemic contacts, the available survey data, and the non-pharmaceutical interventions recorded in the ECDC–JRC Response Measures Database, we extended contact matrices to the full pandemic period (2020–2022), including for countries in which no contact survey was conducted. Two features of the framework are central to its interpretation. First, the estimated time-varying coefficients summarise how the contribution of each pre-pandemic setting (home, work, school, and other places) to pandemic contacts evolves over time. Second, because these coefficients are driven by the observed behaviour of the population, they reflect the combined influence of NPIs and of the behavioural responses, compliance, and seasonal factors that accompanied them, rather than the effect of NPIs in isolation.

Linking the estimated changes to the ECDC–JRC RMD allowed us to characterise the association between the recorded interventions and social mixing, and to generate predictions for periods and countries without survey data. Consistent with this interpretation, the recorded measures aligned most clearly with school-related mixing, whereas variation in adult contacts was less fully explained, indicating that the contact response to policy is partly mediated by behaviour. Because interventions were introduced in overlapping sequences and coincided with voluntary behavioural change and seasonal effects, the relationship we estimate should be read as the change in mixing accompanying a given set of measures, not as the isolated effect of any single intervention.

The implications of this work are substantial for both retrospective analysis and future pandemic preparedness. The reconstructed contact matrices provide a more realistic representation of social mixing during periods of intervention than point estimates of pre-pandemic matrices, and can be incorporated into transmission models, potentially leading to better forecasting and more effective policy decisions. For the countries without CoMix data, they offer a first approximation where none previously existed, contributing to a more complete picture of mixing across the EU/EEA and supporting retrospective analysis and pandemic-preparedness planning.

Our modeling approach acknowledges several limitations that should be considered when interpreting the results.

First, to infer contact change at time points without CoMix data, we assumed that a given set of NPIs had a consistent effect on contacts over time within each country. While behavioral changes during the pandemic likely modified NPI effectiveness over time, these effects are partially captured by the time-varying coefficients in our model. However, without additional data, disentangling the effect of behavioural changes over time, related to compliance with government policies, is non-trivial. Second, the quantity of available data creates important constraints. Most countries had only around seven CoMix survey waves, frequently with substantial gaps between collections and a particularly limited availability of children’s contact data, which limits the resolution of the estimated coefficient functions and increases the uncertainty of the resulting predictions. Third, the survey data itself is subject to the biases common to self-reported contact surveys, like selection, recall, and reporting effects, which the survey design and weighting procedures mitigate but do not remove. The country-matching procedure adds a further layer of uncertainty for countries without direct survey data. Finally, the absence of independent contact data for these countries limits our ability to validate the reconstructed contacts directly.

Future work should prioritise validation against independent data sources, including comparison of the reconstructed matrices with observed case trajectories, and the development of models that represent behavioural adaptation explicitly rather than absorbing it into the time-varying coefficients. Additionally, extending this approach to countries beyond Europe would provide valuable insights into how cultural and socioeconomic factors influence the relationship between NPIs and social mixing patterns.

## Data Availability

All data used in this work are publicly available. The underlying CoMix social contact data are publicly available from https://socialcontactdata.org/data/. The pre-pandemic contact matrices used in this
study were obtained from Prem et al., available at https://journals.plos.org/ploscompbiol/article?id=10.1371/journal.pcbi.1005697#sec020. The ECDC-JRC Response Measures Database is available from https://www.ecdc.europa.eu/en/publications-data/response-measures-database-rmd.
All data and code supporting this work are publicly available. The analysis code, data
used in the analyses, and generated age-specific contact matrices are available in
the project GitHub repository: https://github.com/EU-ECDC/COVID19SocialPatterns.
The data directory is available at https://github.com/EU-
ECDC/COVID19SocialPatterns/tree/main/data.

https://journals.plos.org/ploscompbiol/article?id=10.1371/journal.pcbi.1005697#sec020

https://www.ecdc.europa.eu/en/publications-data/response-measures-database-rmd

## Supplementary material

Supplementary material containing additional figures is available for this article.

## Data and code availability

All data used in this study can be downloaded from the cited references. The codes used to generate these analyses and the generated age-specific contact matrices are available at https://github.com/EU-ECDC/COVID19SocialPatterns.

## Author contributions

Anastasia Chatzilena: Conceptualization, methodology, formal analysis, visualization, writing – original draft preparation. Ellen Brooks-Pollock: Conceptualization, review and editing. Ewan Colman: Data curation, visualization, review and editing. Disa Hansson: review and editing. Nikolaos Demiris: Methodology, review and editing. Bastian Franklin: Conceptualization, review and editing. Leon Danon: Conceptualization, review and editing. All authors contributed to discussions throughout the project and reviewed and approved the final manuscript.

## Acknowledgements

AC acknowledges support from the Wellcome Trust (227438/Z/23/Z). EBP acknowledges the support of the National Institute for Health Research Health Protection Research Unit (NIHR HPRU) in Evaluations and Behavioural Science. LD is funded by the UK Research and Innovation AI programme of the Engineering and Physical Sciences Research Council, AI for Collective Intelligence Research Hub (EPSRC grant EP/Y028392/1). LD and EBP are funded by the NIHR Health Protection Research Focus Award in Vaccines at the University of Bristol.

