## Supplementary figures for "Reconstructing age-specific social contact patterns across 28 EU/EEA countries during the COVID-19 pandemic: linking non-pharmaceutical interventions to social mixing, 2020–2022"

### Overview of supplementary contents

- **Figure S1.** Visualisations of the ECDC-JRC response measures (NPIs) for 30 EU/EEA countries, shown alongside CoMix mean contacts and COVID-19 deaths where available.
- **Figure S2.** Estimated time-varying coefficient functions ($\beta_p(w)$, Level 1) for home, work, school and other places, for the 17 countries with CoMix data.
- **Figure S3.** Level 1 fitted number of contacts between age groups (medians and 95% CIs) against the reported CoMix contacts, for CoMix countries.
- **Figure S4.** NPI-predicted change in contacts by location over time ($\beta_p(t)$, Level 2), for CoMix countries.
- **Figure S5.** Level 2 predicted contacts based on the posterior medians of $\beta_t$, against reported CoMix contacts.
- **Figure S6.** Level 2 predicted contacts based on the 2.5%, 50% and 97.5% posterior percentiles of $\beta_t$.
- **Figure S7.** Country matching: hierarchical clustering of NPI patterns and distance-based matching.
- **Figure S8.** Reconstructed contacts for countries without CoMix data, each derived from its matched CoMix country.

(a)

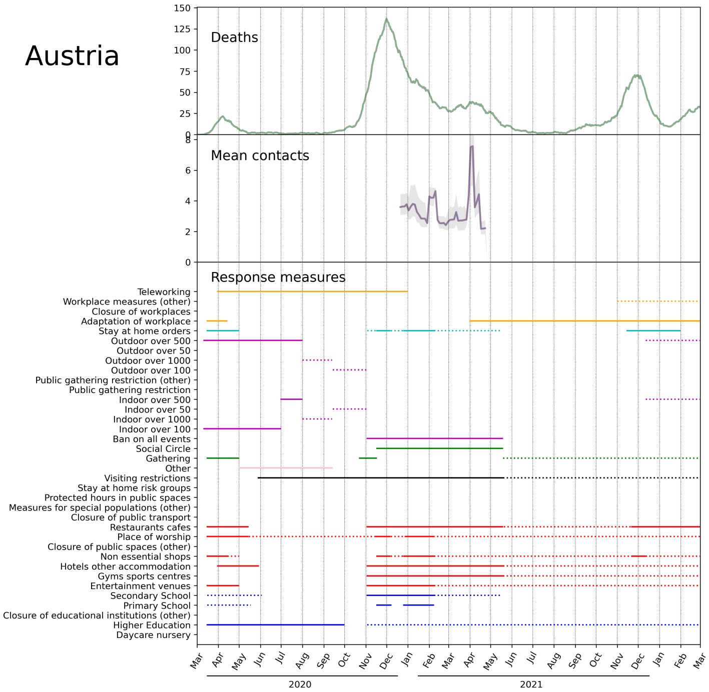


(b)

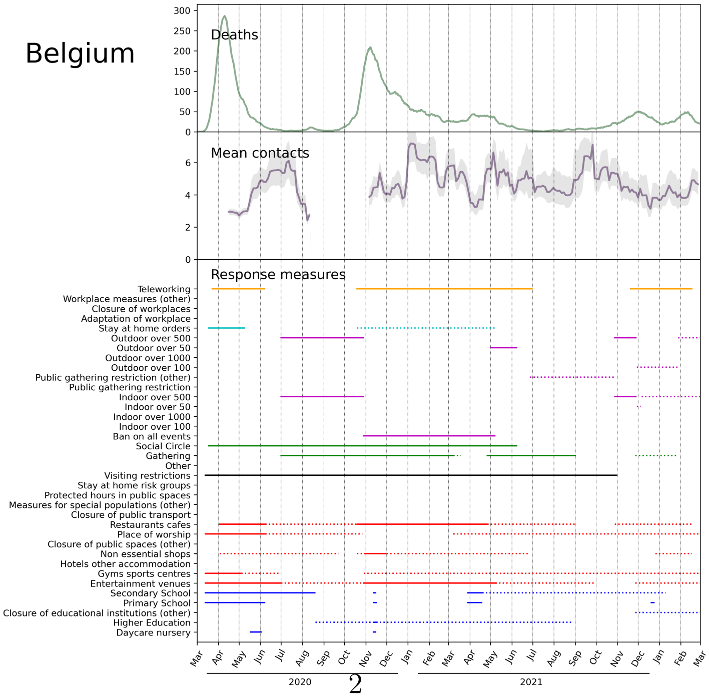




Figure S1: Visualisations of ECDC response measures data for 30 countries along with CoMix mean contacts and COVID-19 deaths where available

(c)

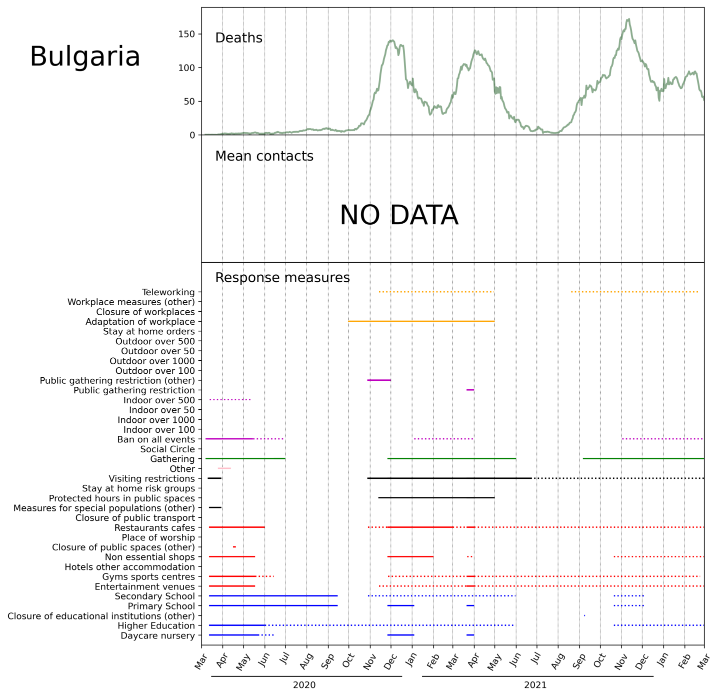


(d)

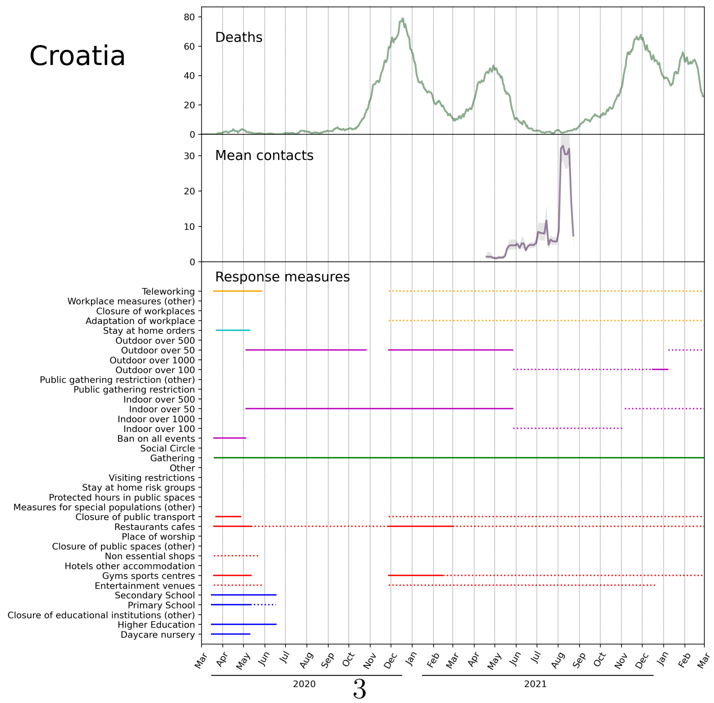




Figure S1: Visualisations of ECDC response measures data (continued)

(e)

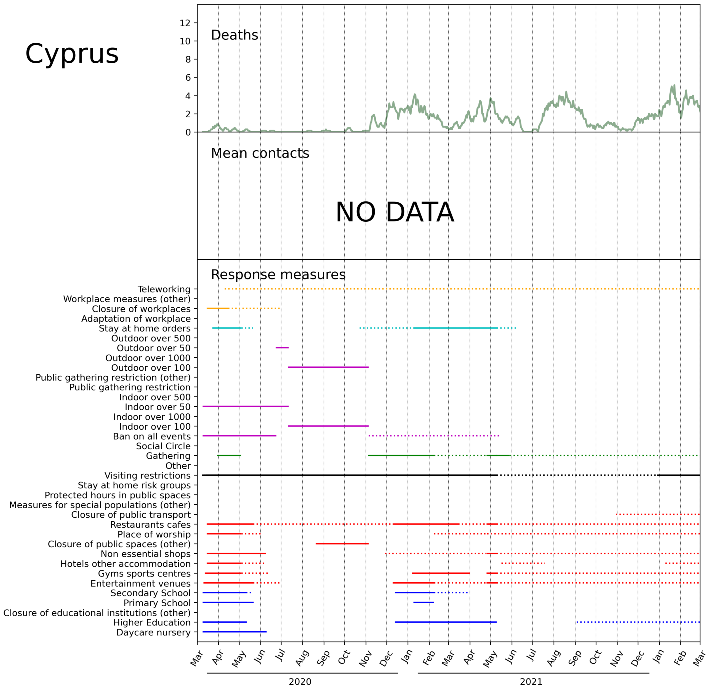


(f)

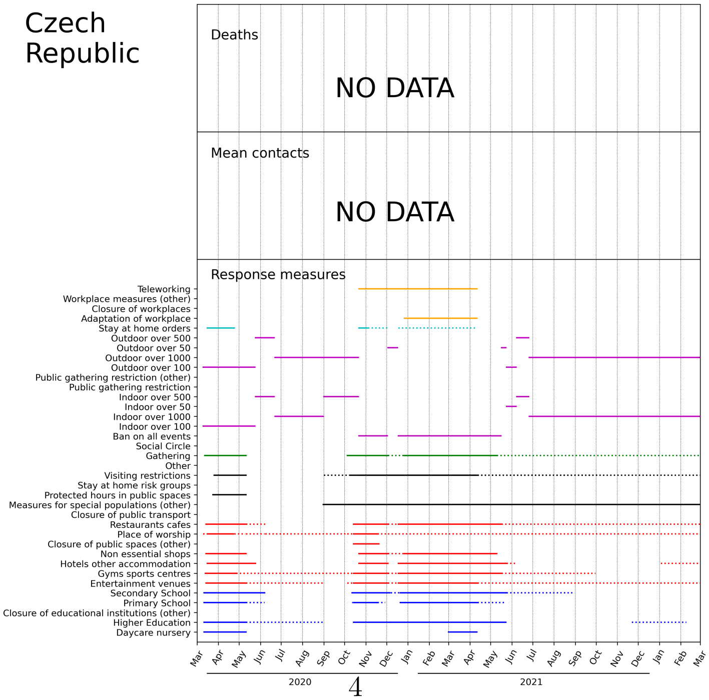




Figure S1: Visualisations of ECDC response measures data (continued)

(g)

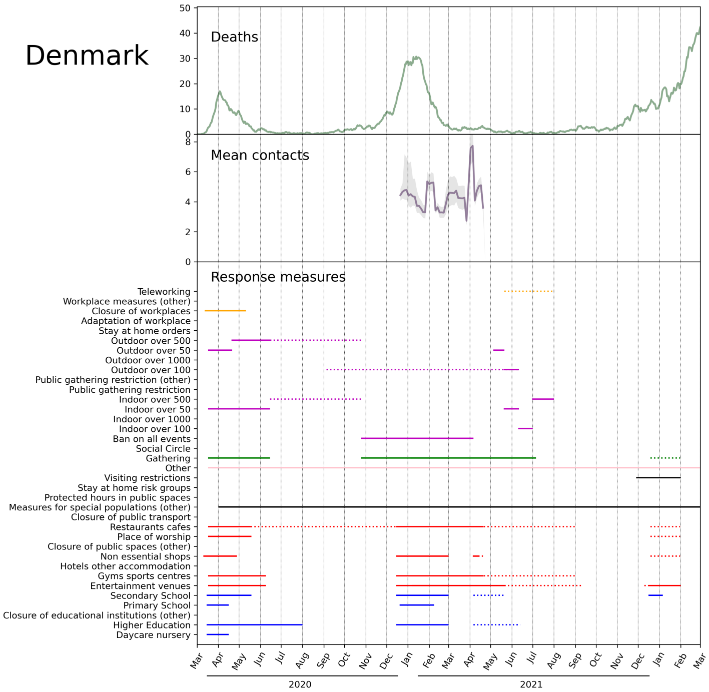


(h)

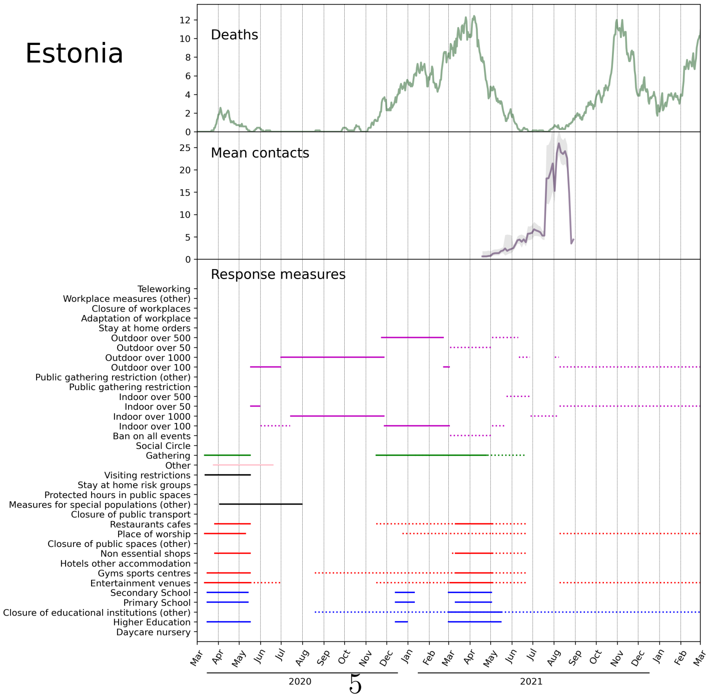




Figure S1: Visualisations of ECDC response measures data (continued)

(i)

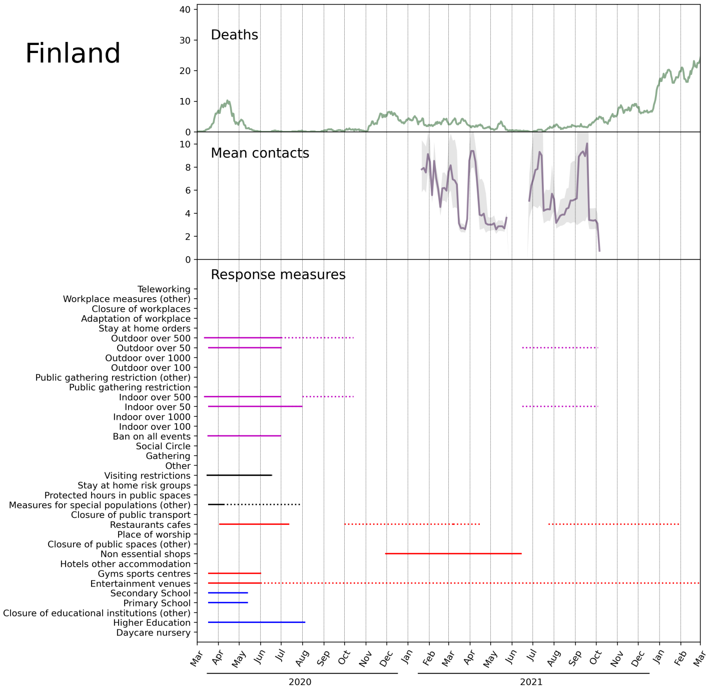


(j)

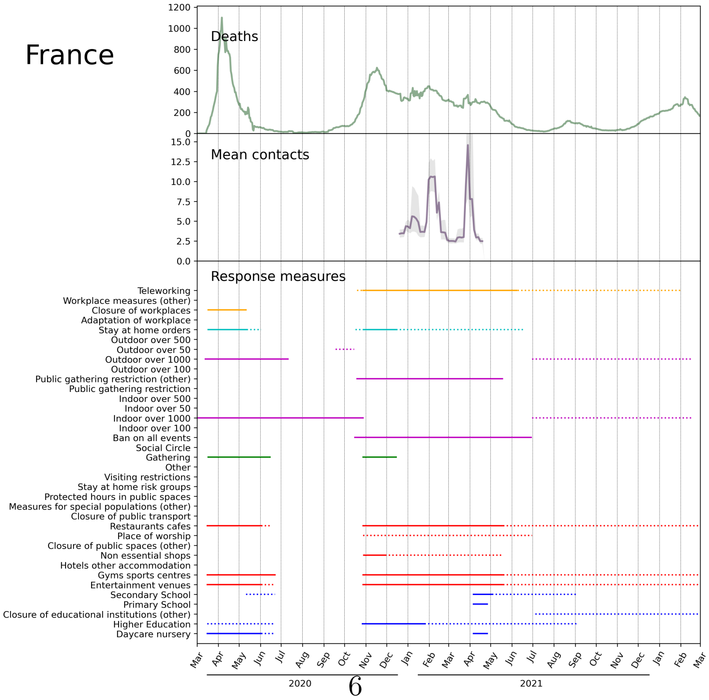




Figure S1: Visualisations of ECDC response measures data (continued)

(k)

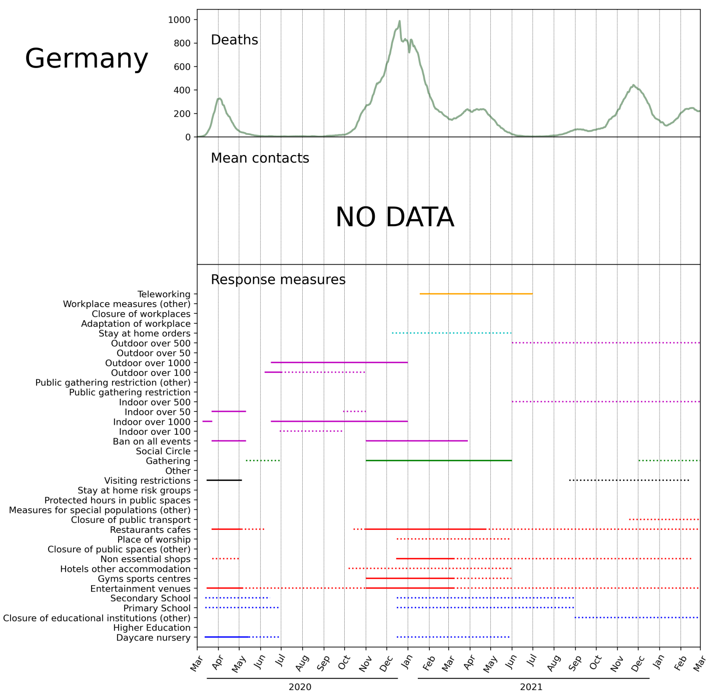


(l)

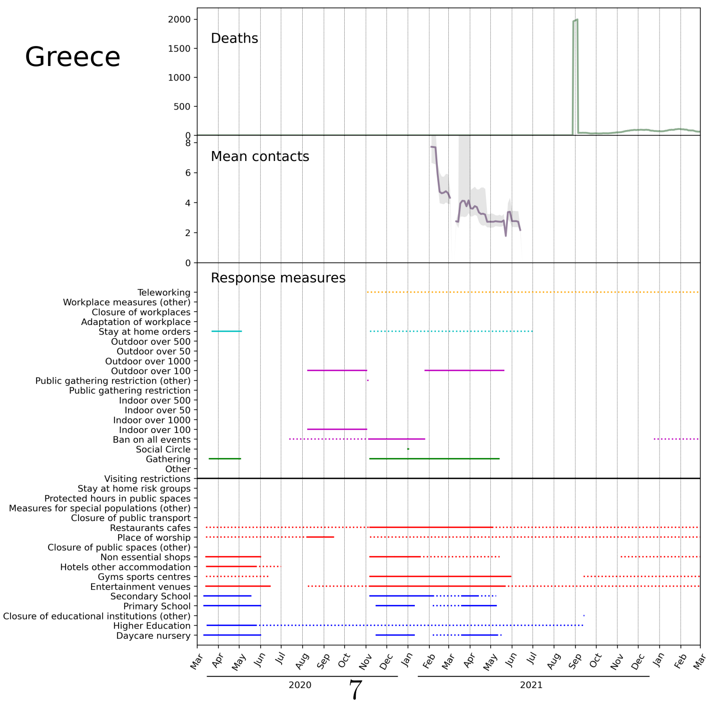




Figure S1: Visualisations of ECDC response measures data (continued)

(m)

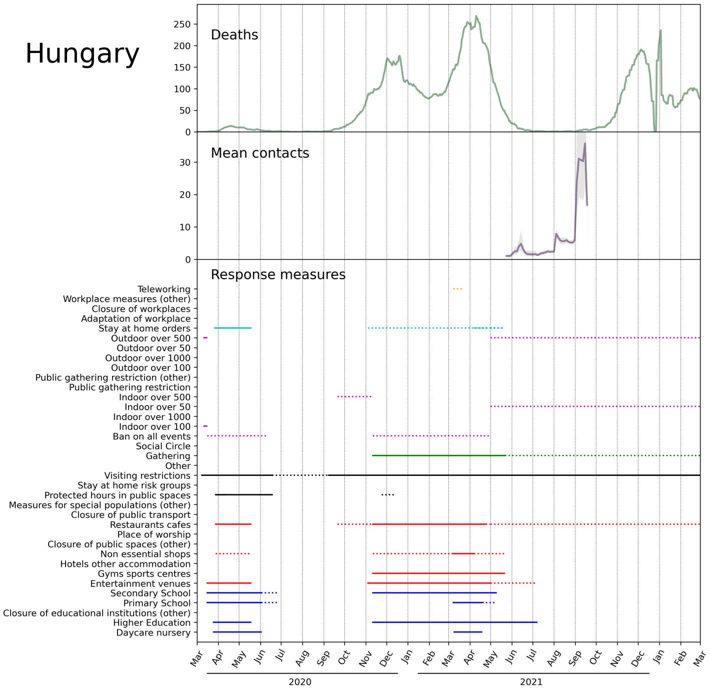


(n)

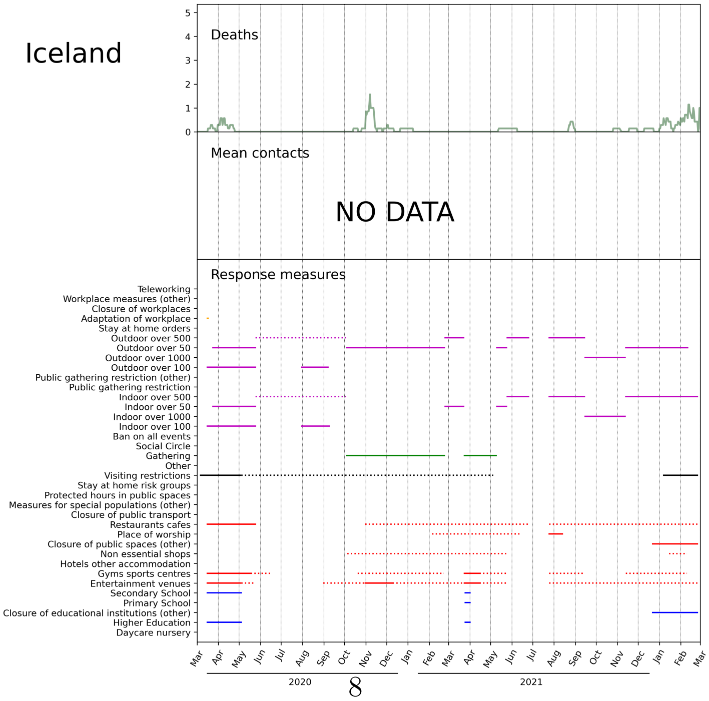




Figure S1: Visualisations of ECDC response measures data (continued)

(o)

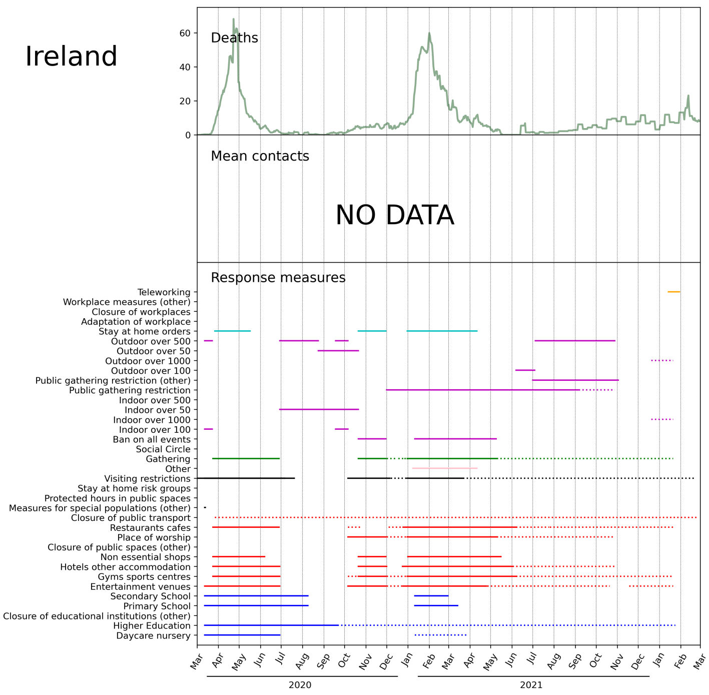


(p)

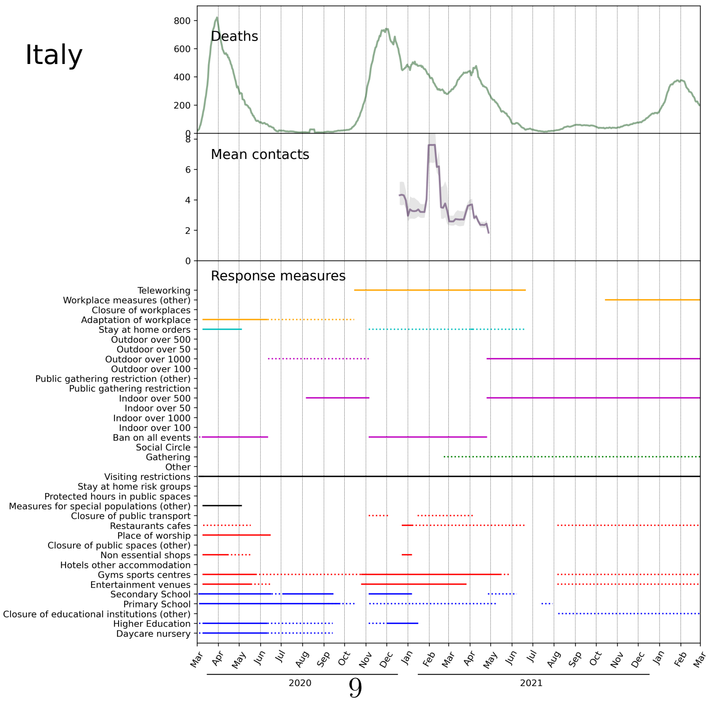




Figure S1: Visualisations of ECDC response measures data (continued)

(q)

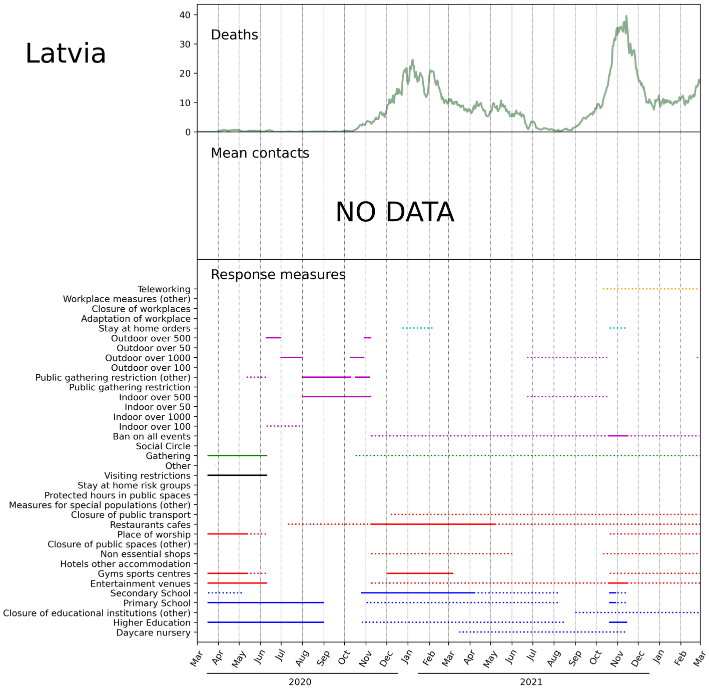


r)

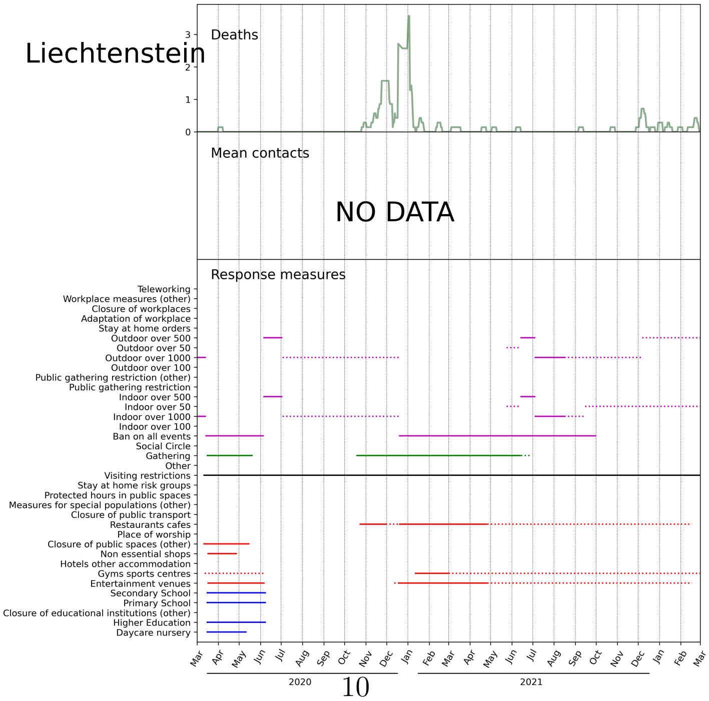




Figure S1: Visualisations of ECDC response measures data (continued)

(s)

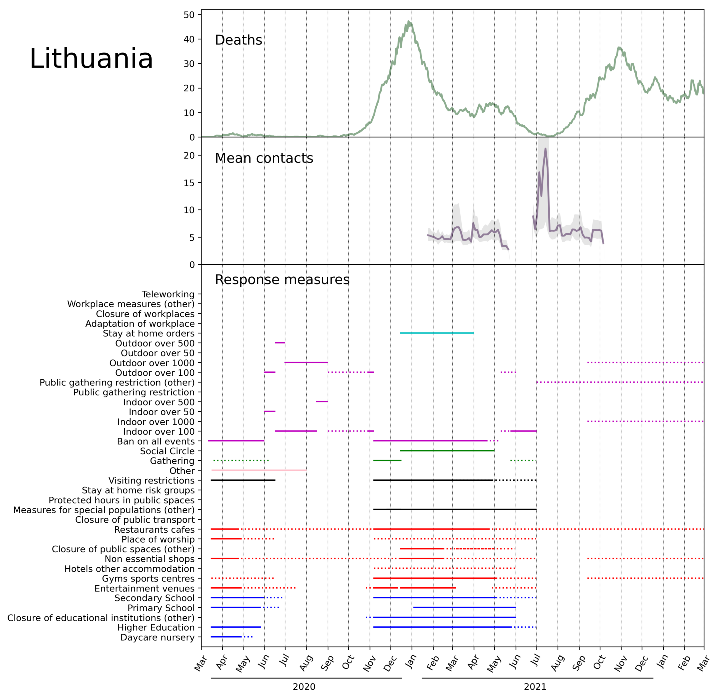


(t)

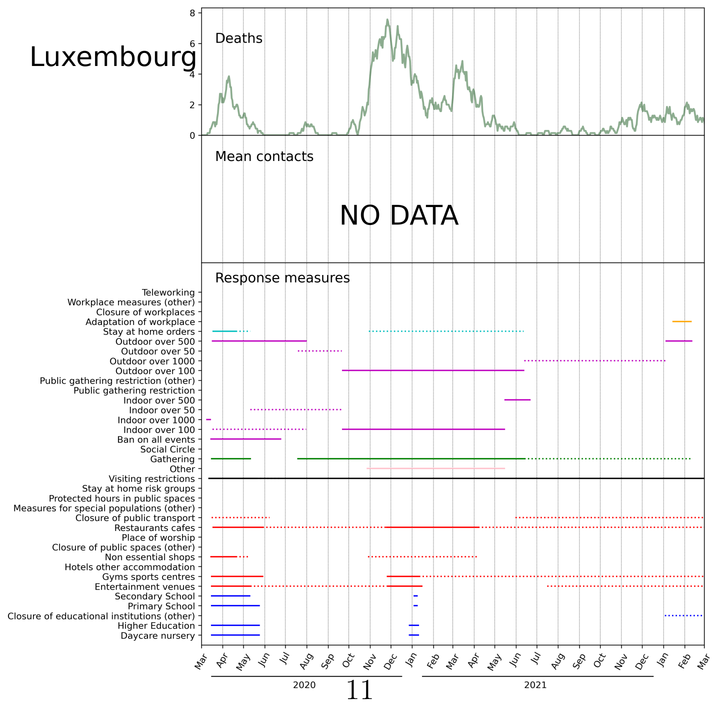




Figure S1: Visualisations of ECDC response measures data (continued)

(u)

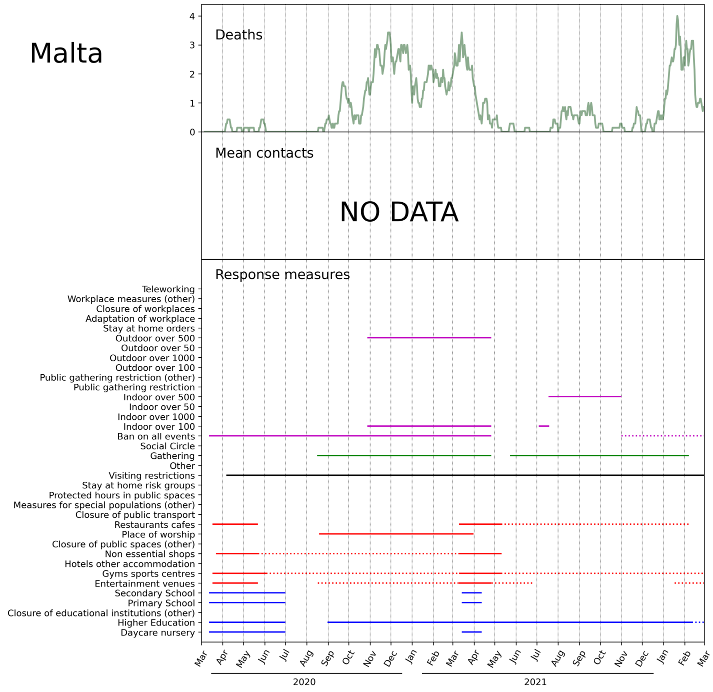


(v)

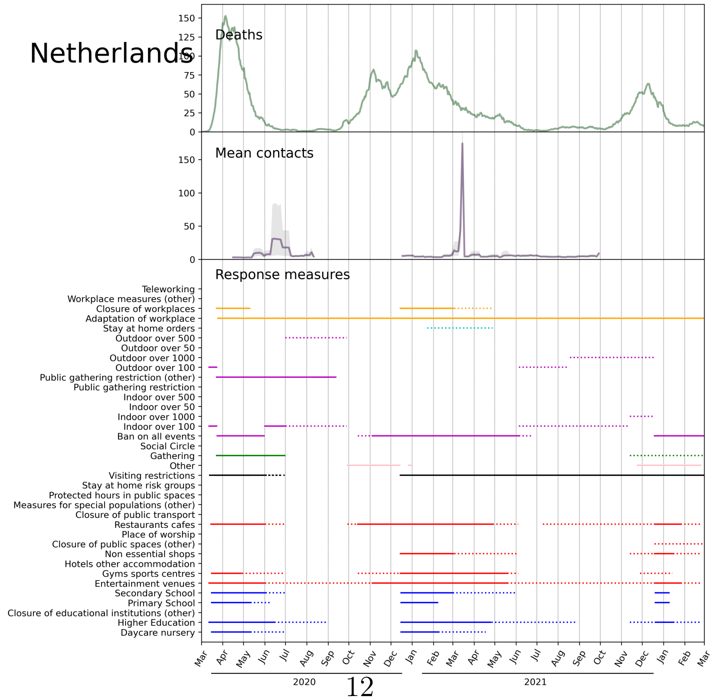




Figure S1: Visualisations of ECDC response measures data (continued)

w

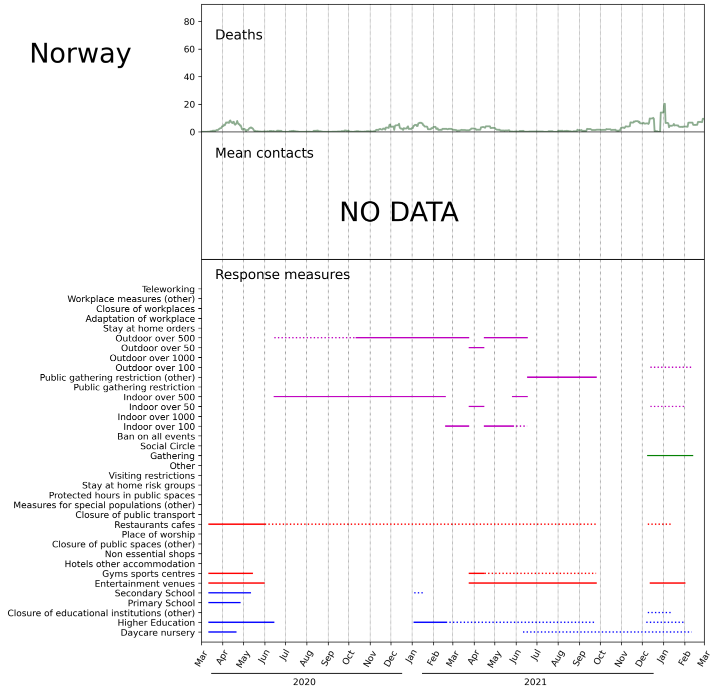


x

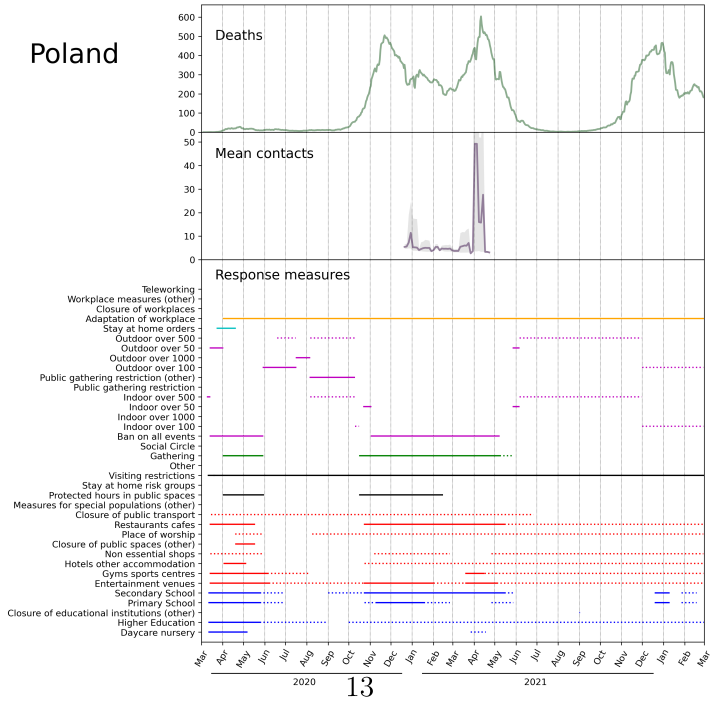




Figure S1: Visualisations of ECDC response measures data (continued)

y

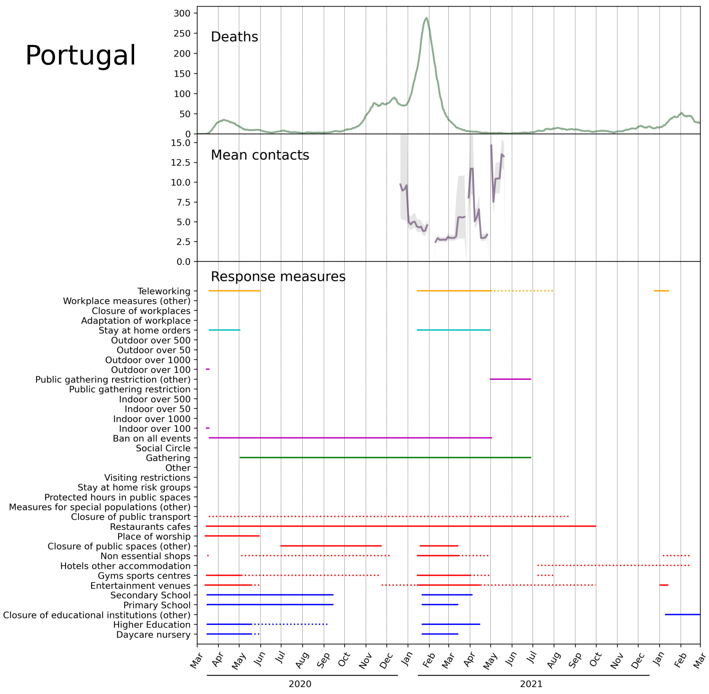


z

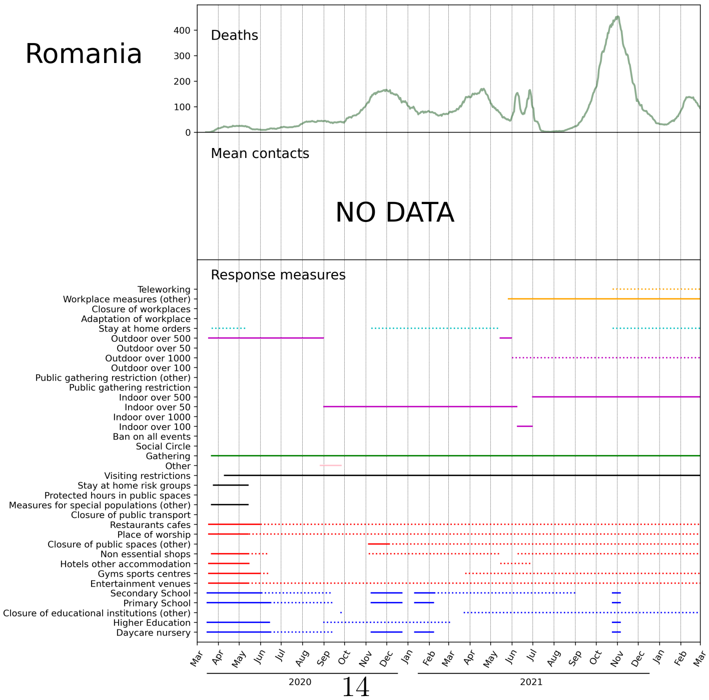




Figure S1: Visualisations of ECDC response measures data (continued)

(aa)

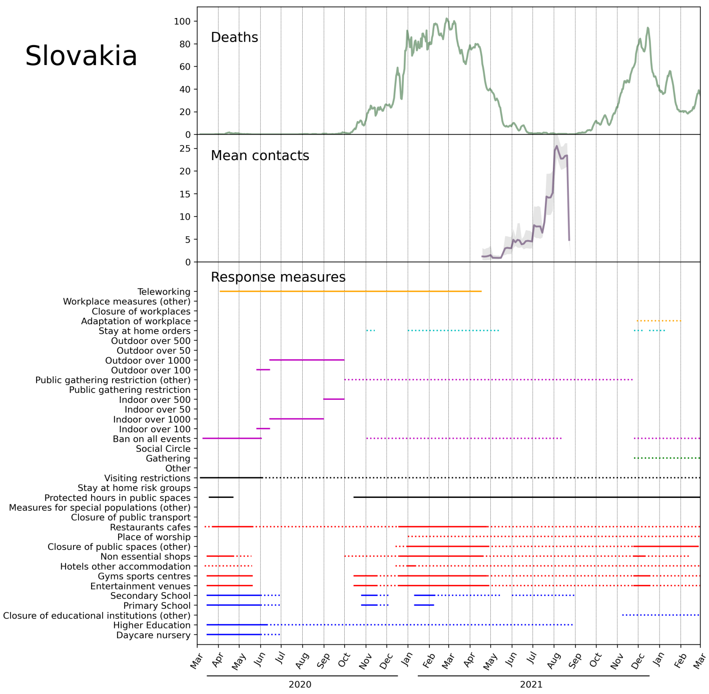


(bb)

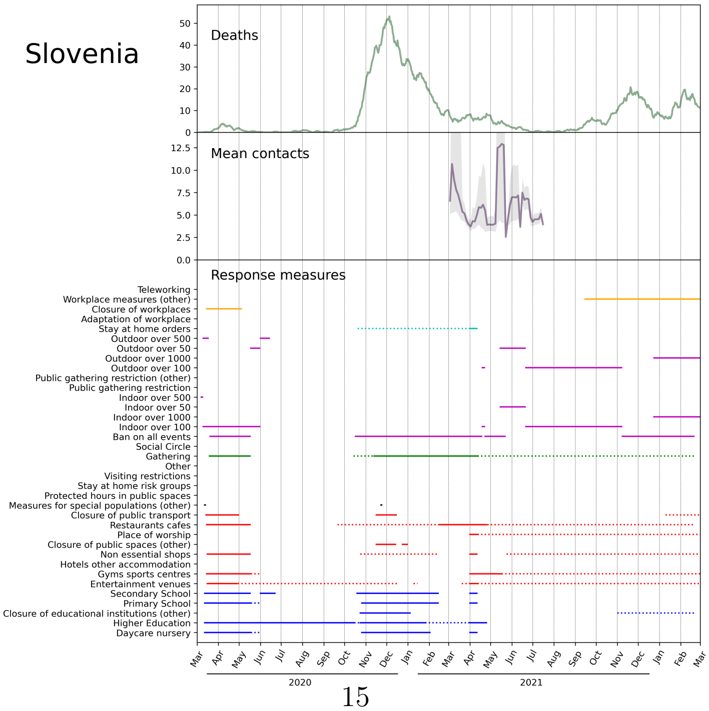




Figure S1: Visualisations of ECDC response measures data (continued)

(cc)

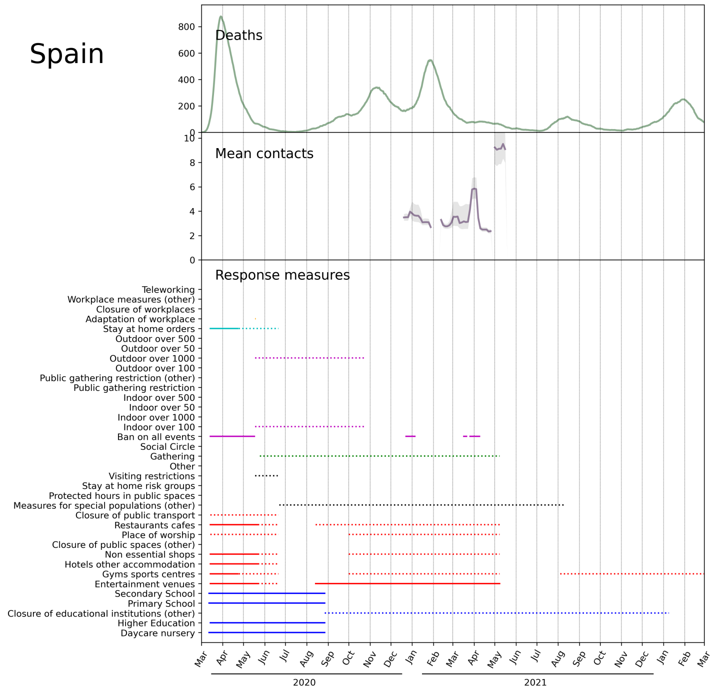


(dd)

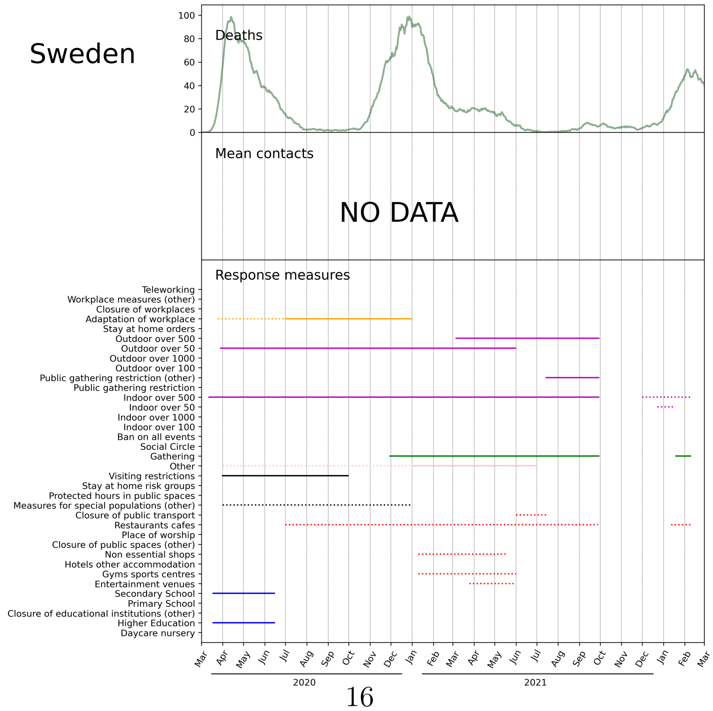




Figure S1: Visualisations of ECDC response measures data (continued)

(Austria)

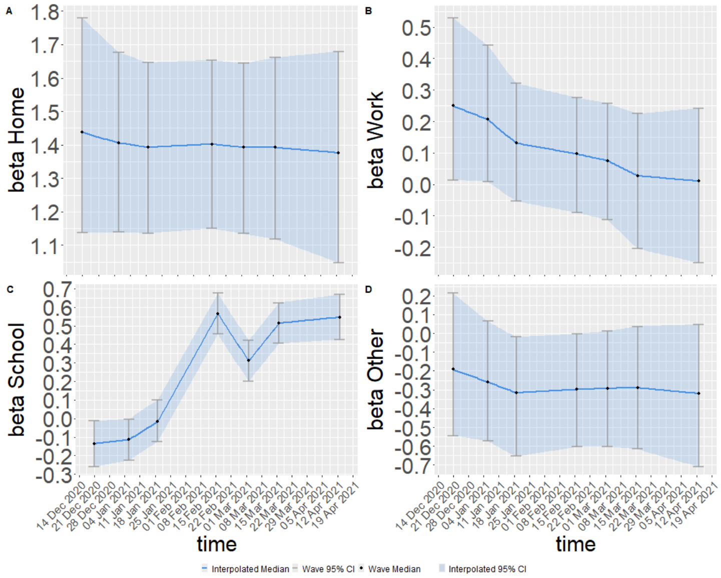


(Belgium)

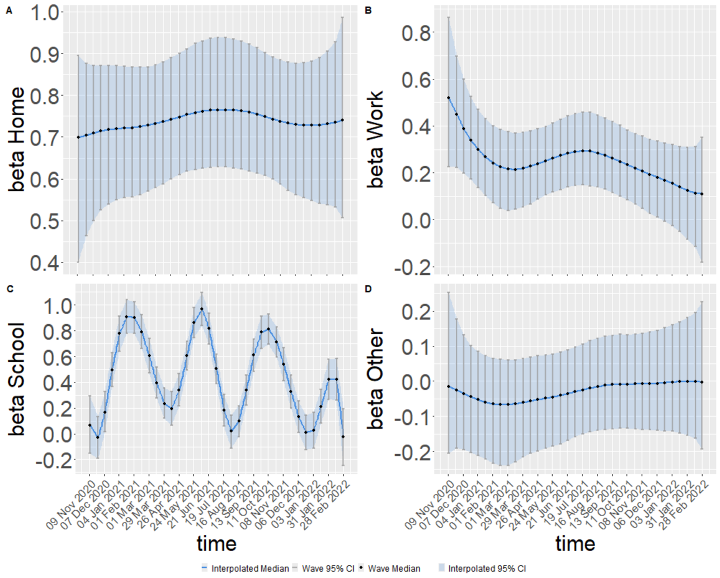


Figure S2: Estimated coefficient functions of the TVEM for the different locations; home, work, school and other places.

(Croatia)

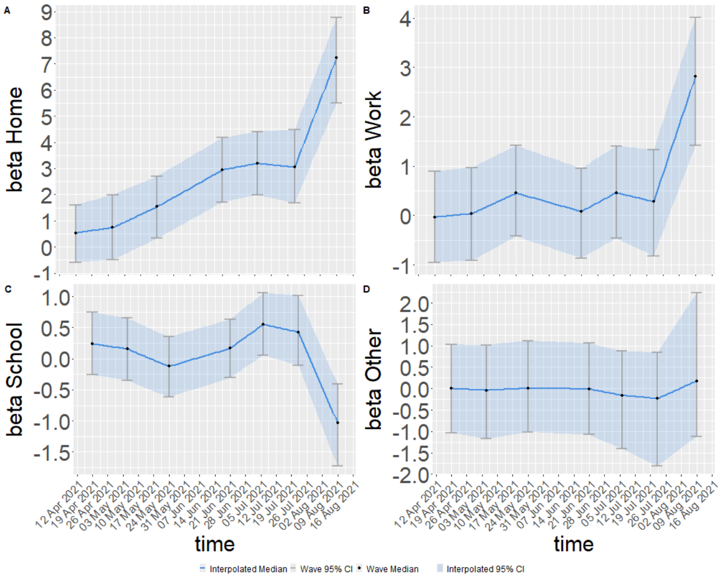


(Denmark)

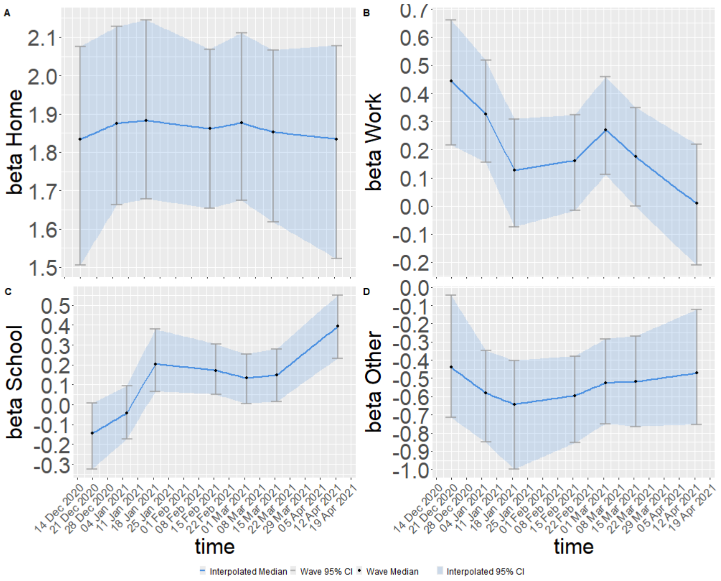


Figure S2: Estimated coefficient functions of the TVEM for the different locations; home, work, school and other places. (continued)

(Estonia)

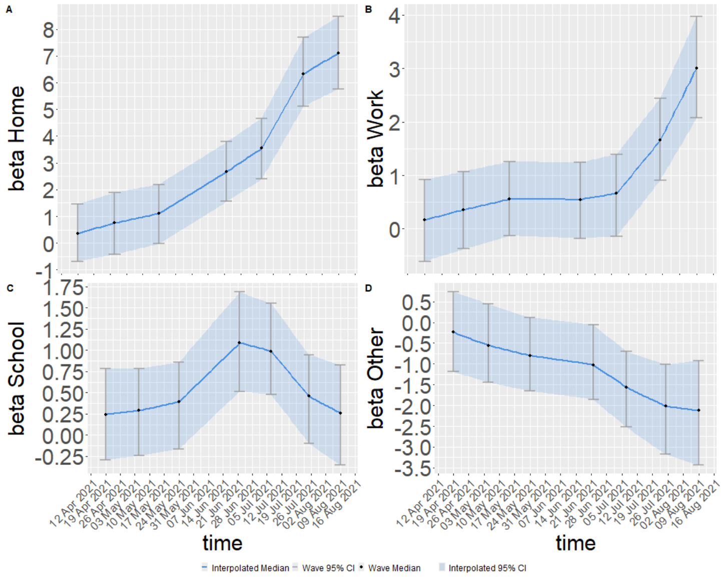


(Finland)

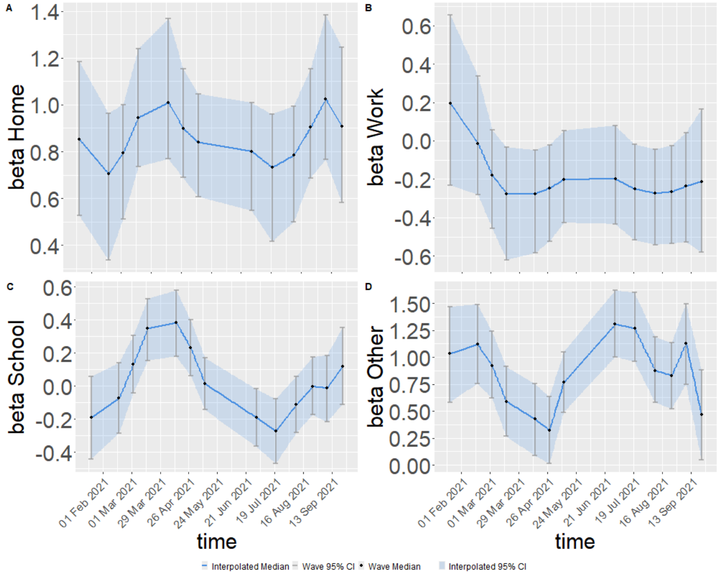


Figure S2: Estimated coefficient functions of the TVEM for the different locations; home, work, school and other places. (continued)

(France)

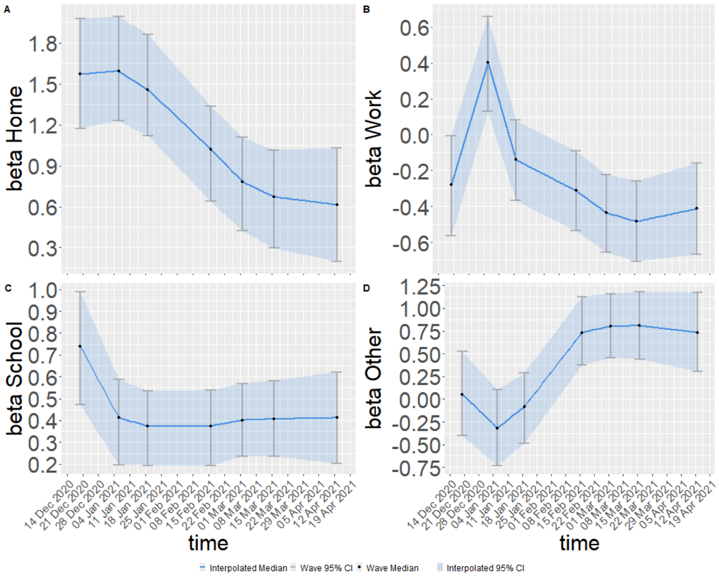


(Greece)

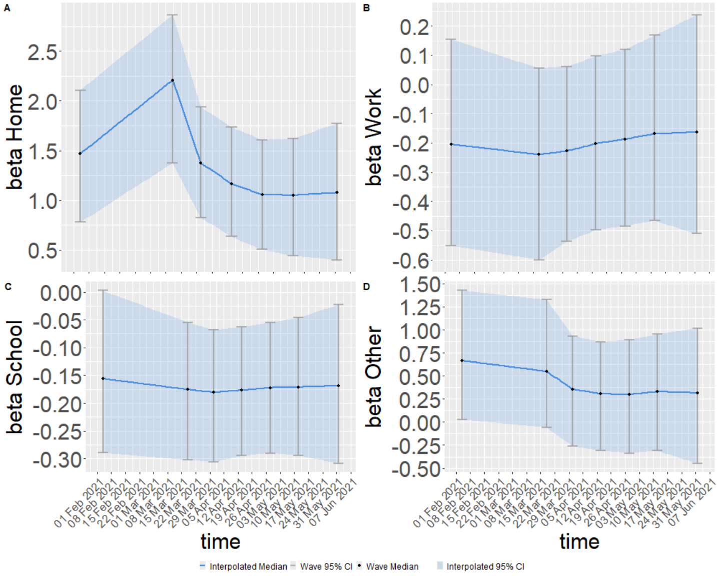


Figure S2: Estimated coefficient functions of the TVEM for the different locations; home, work, school and other places. (continued)

(Hungary)

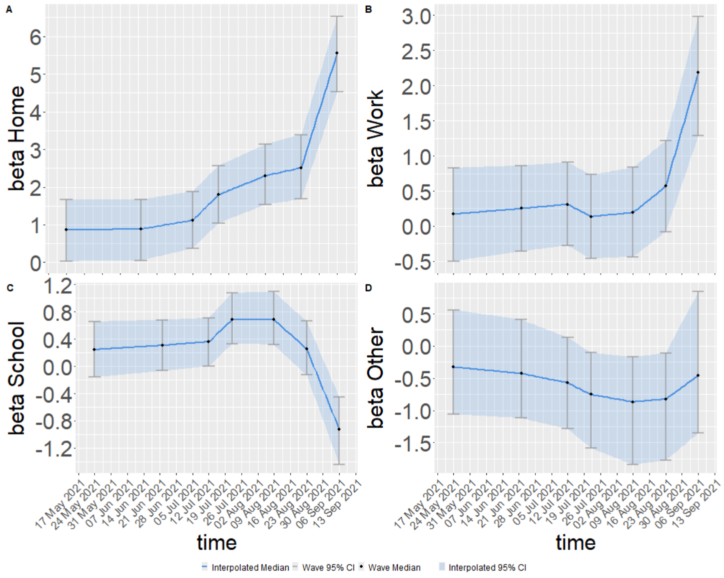


(Italy)

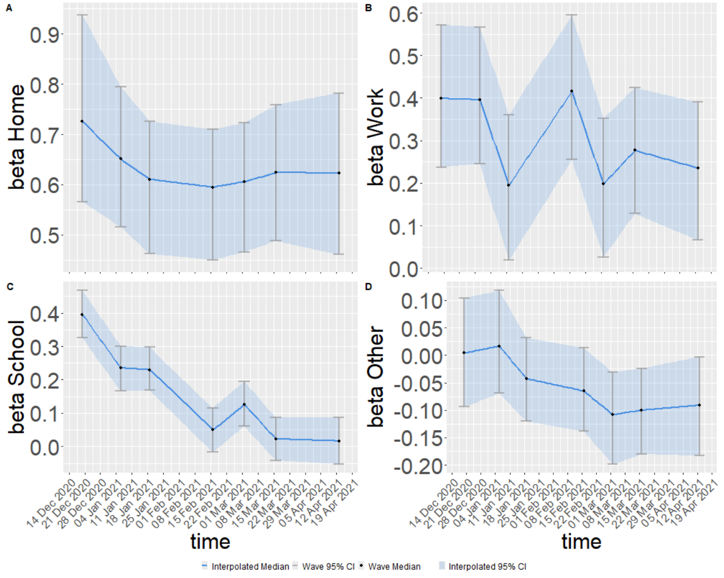


Figure S2: Estimated coefficient functions of the TVEM for the different locations; home, work, school and other places. (continued)

(Lithuania)

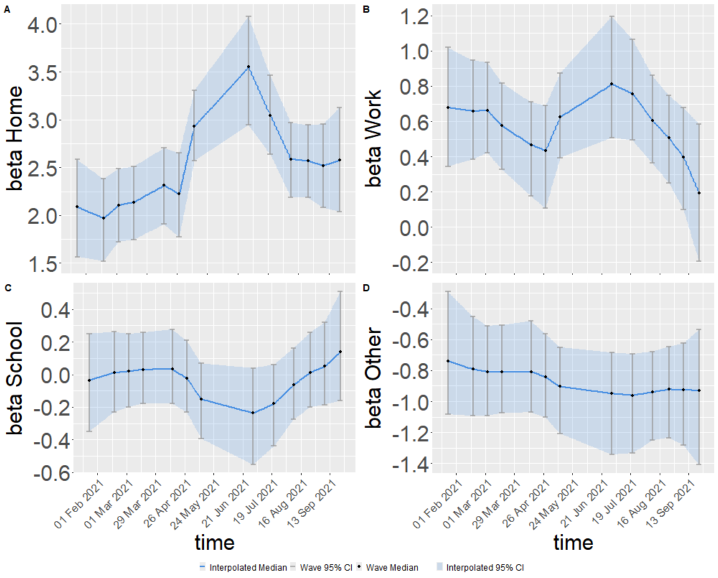


(Netherlands)

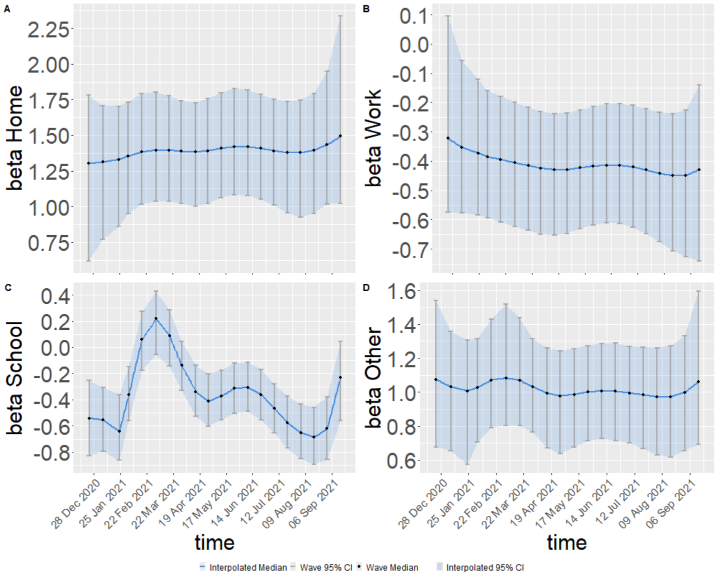


Figure S2: Estimated coefficient functions of the TVEM for the different locations; home, work, school and other places. (continued)

(Poland)

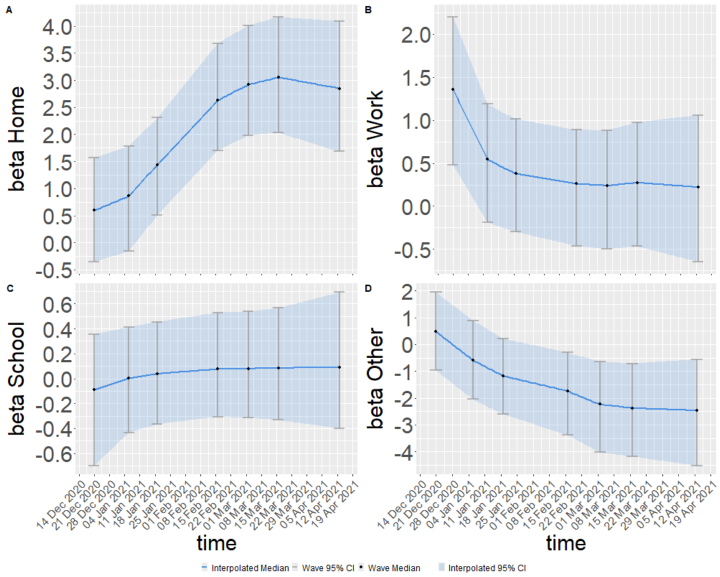


(Portugal)

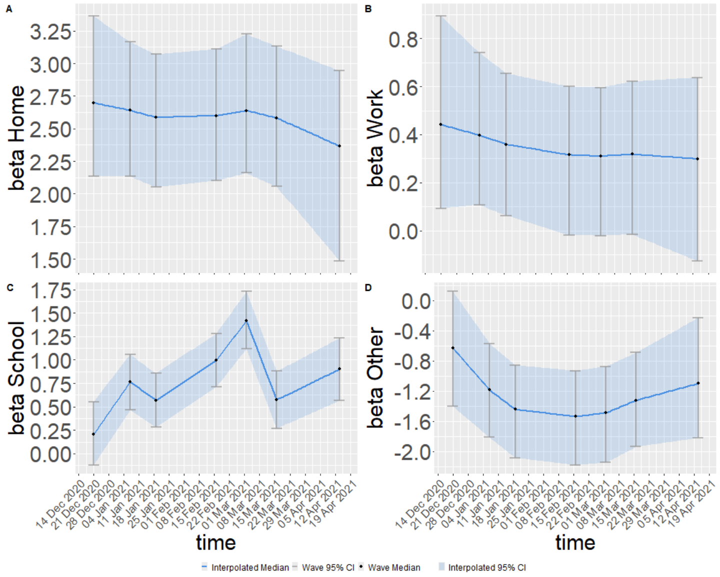


Figure S2: Estimated coefficient functions of the TVEM for the different locations; home, work, school and other places. (continued)

(Slovakia)

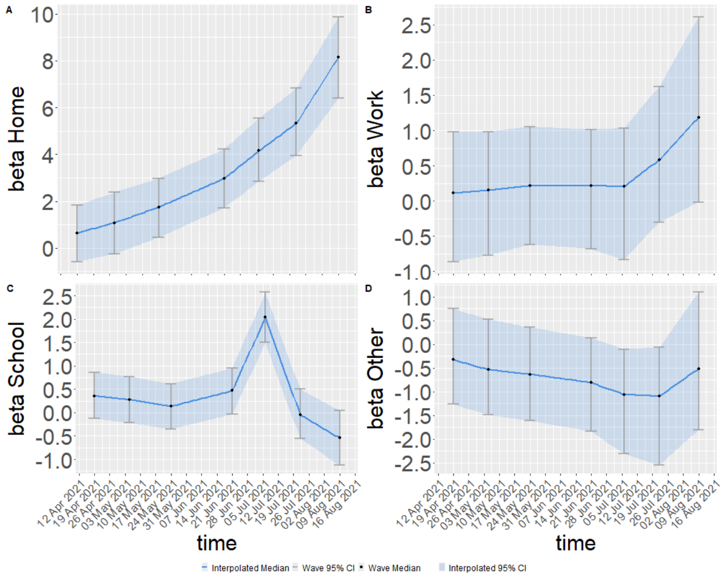


(Slovenia)

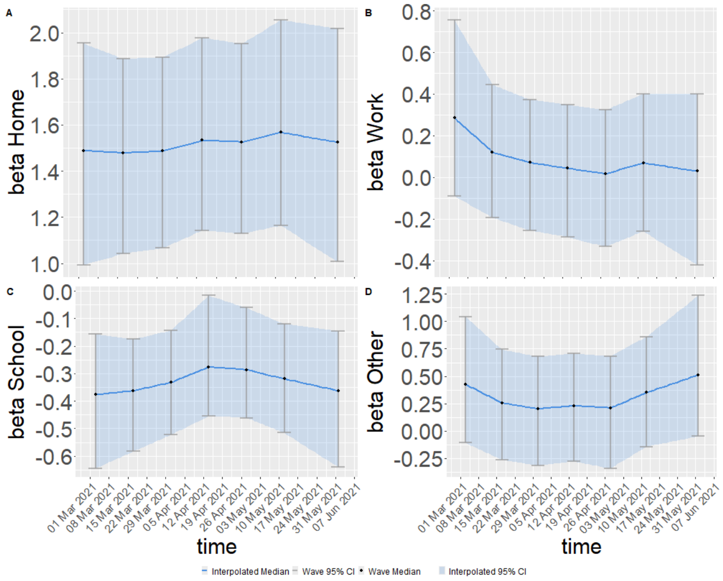


Figure S2: Estimated coefficient functions of the TVEM for the different locations; home, work, school and other places. (continued)

(Spain)

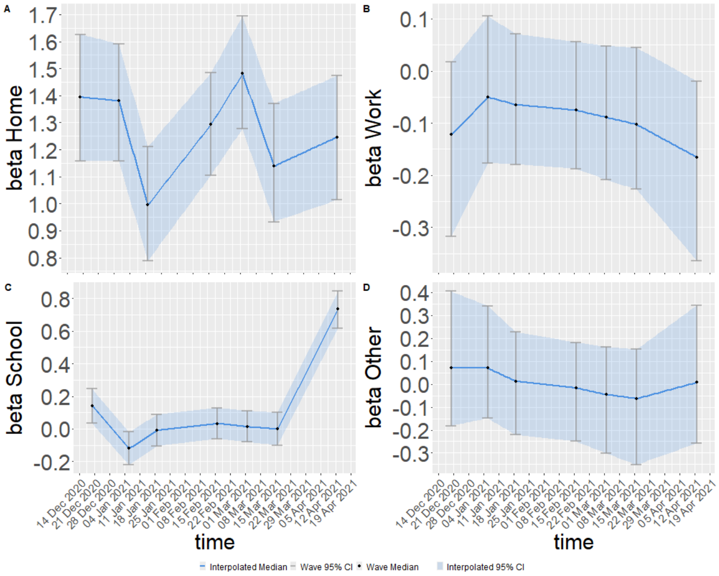


Figure S2: Estimated coefficient functions of the TVEM for the different locations; home, work, school and other places. (continued)

(Austria)

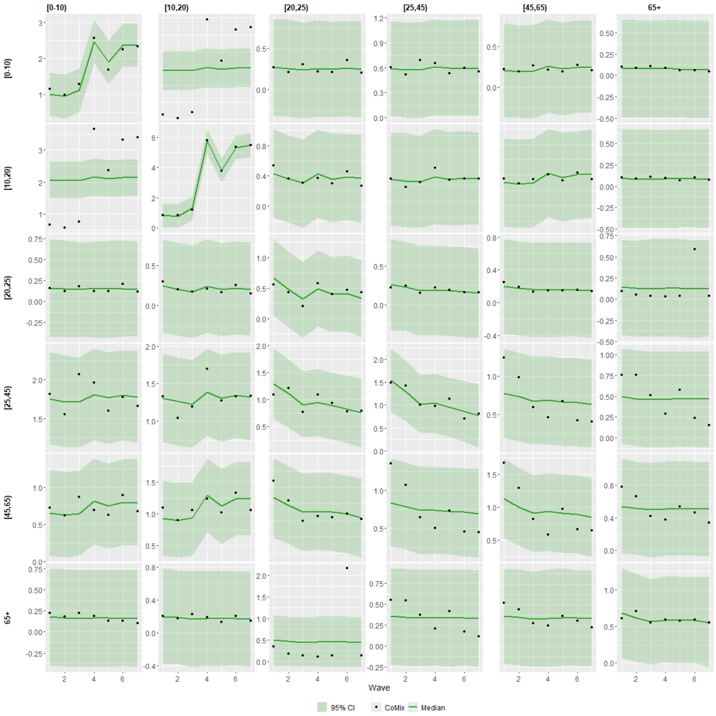

(Belgium)

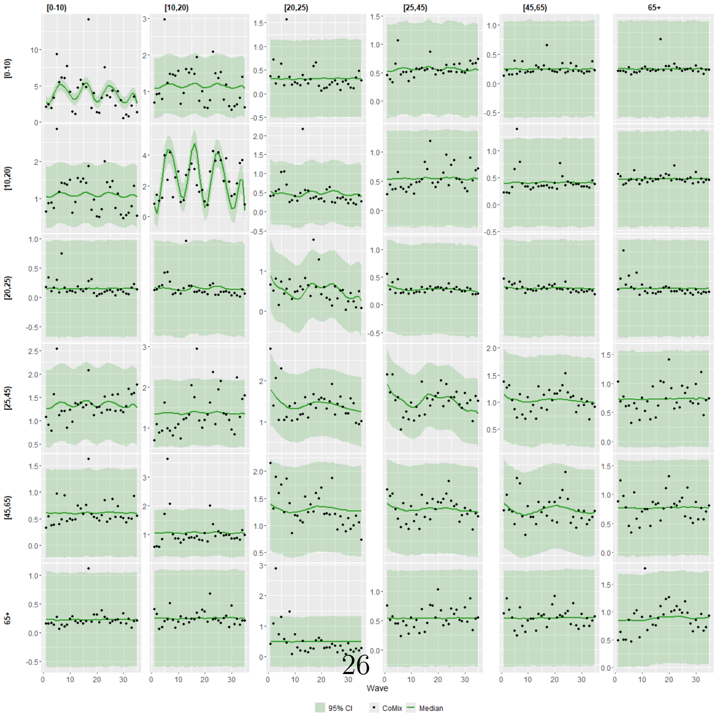

Figure S3: Estimated number of contacts between age groups, medians (lines) and 95% CI (dark shaded areas) along with reported number of contacts from CoMix(dots).

(Croatia)

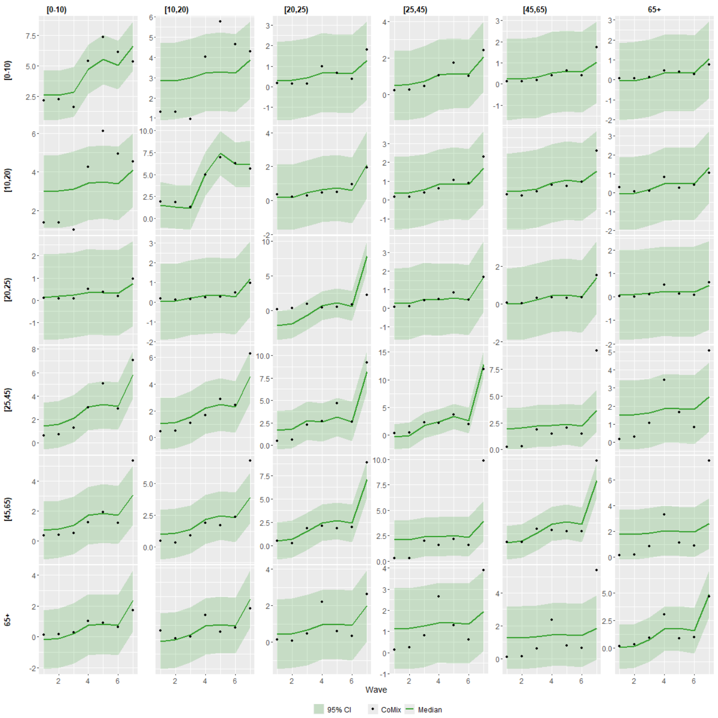


(Denmark)

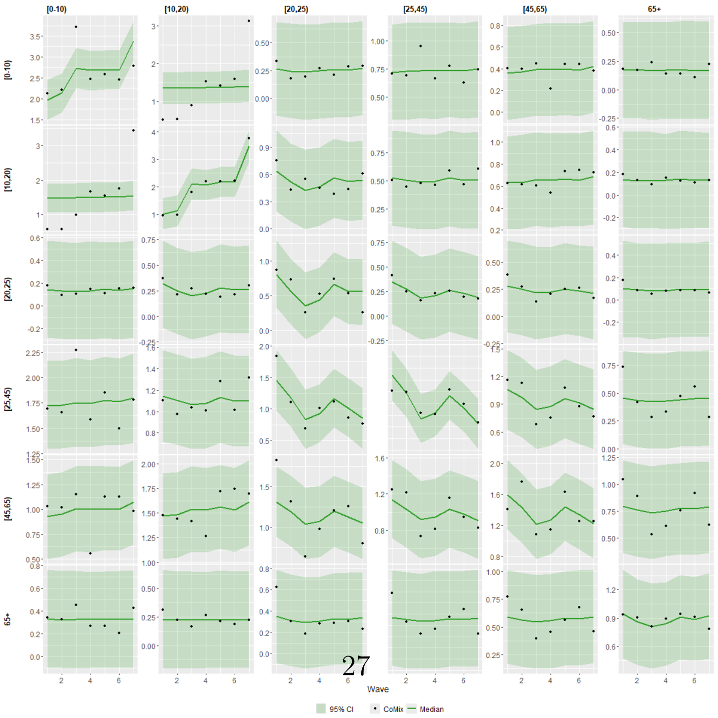


Figure S3: Estimated number of contacts between age groups, medians (lines) and 95% CI (dark shaded areas) along with reported number of contacts from CoMix(dots). (continued)

(Estonia)

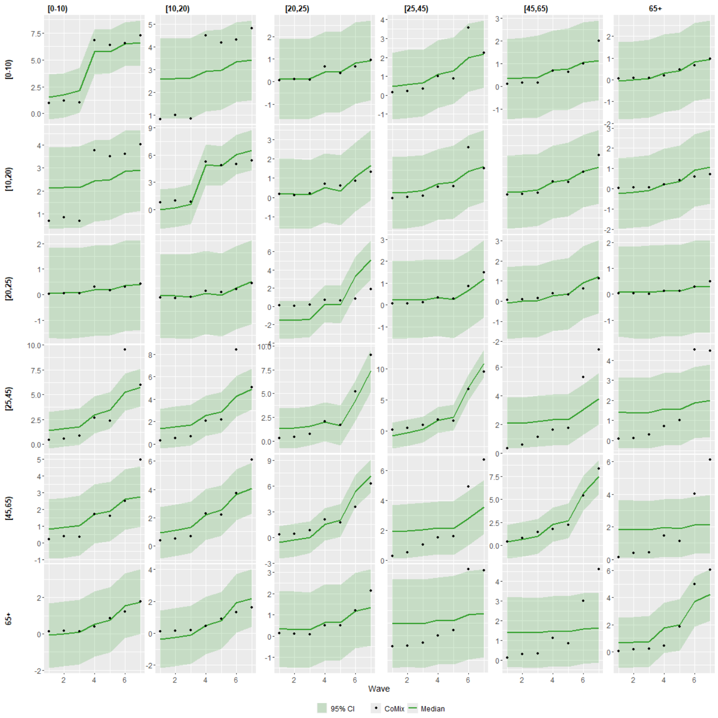


(Finland)

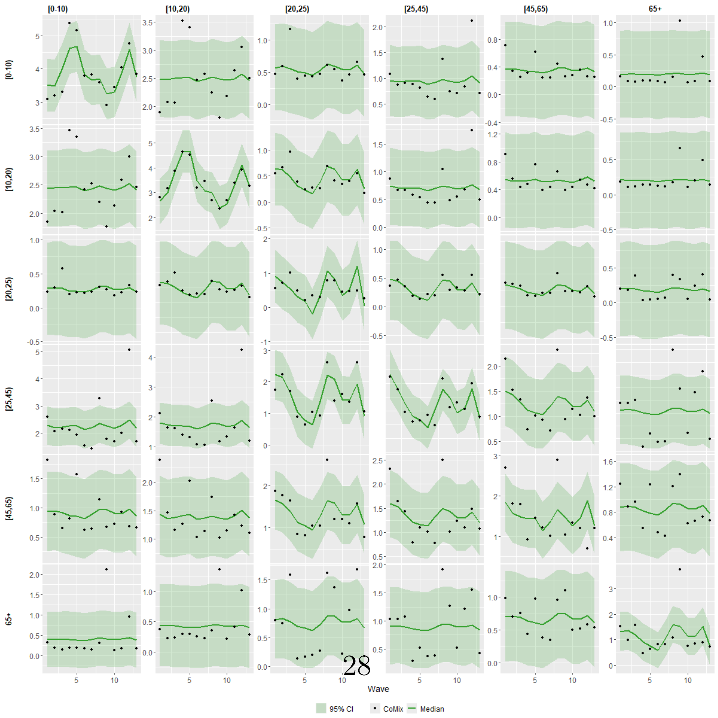




Figure S3: Estimated number of contacts between age groups, medians (lines) and 95% CI (dark shaded areas) along with reported number of contacts from CoMix(dots). (continued)

(France)

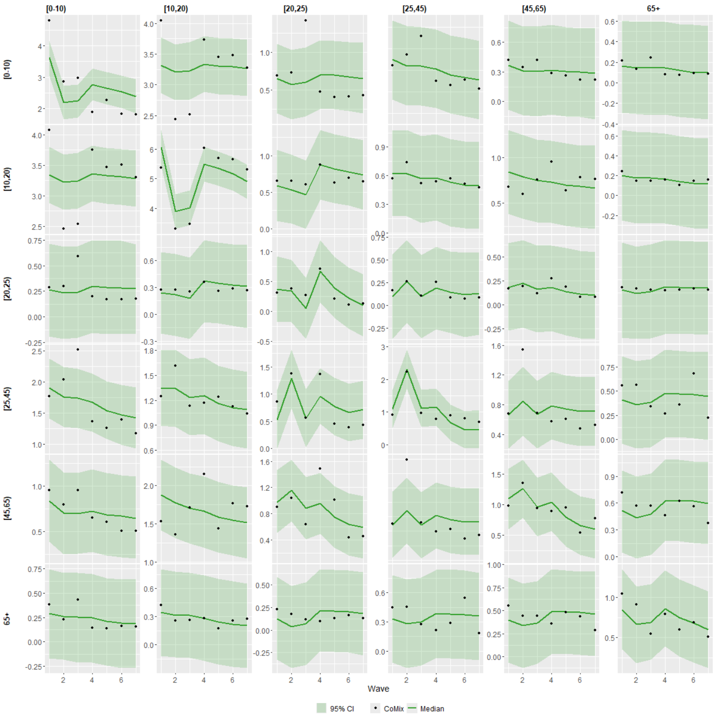


(Greece)

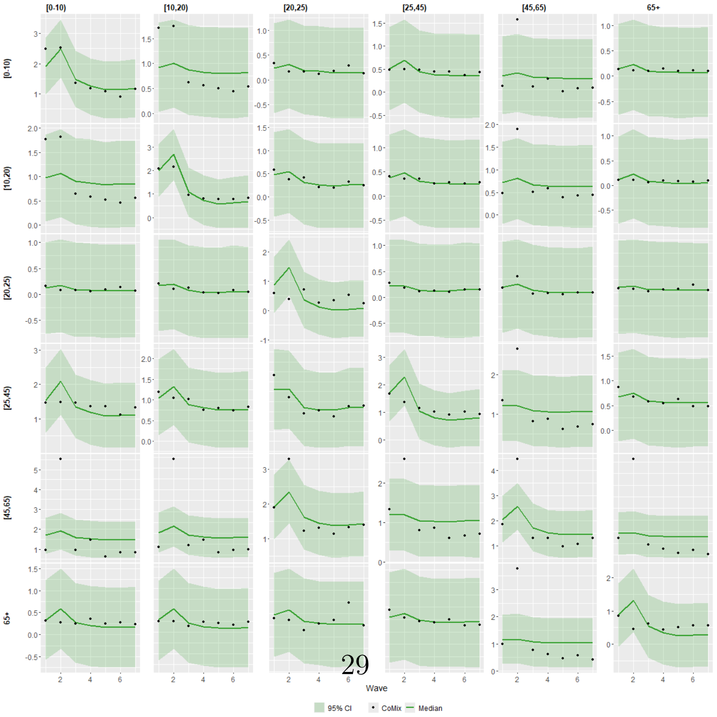




Figure S3: Estimated number of contacts between age groups, medians (lines) and 95% CI (dark shaded areas) along with reported number of contacts from CoMix(dots). (continued)

(Hungary)

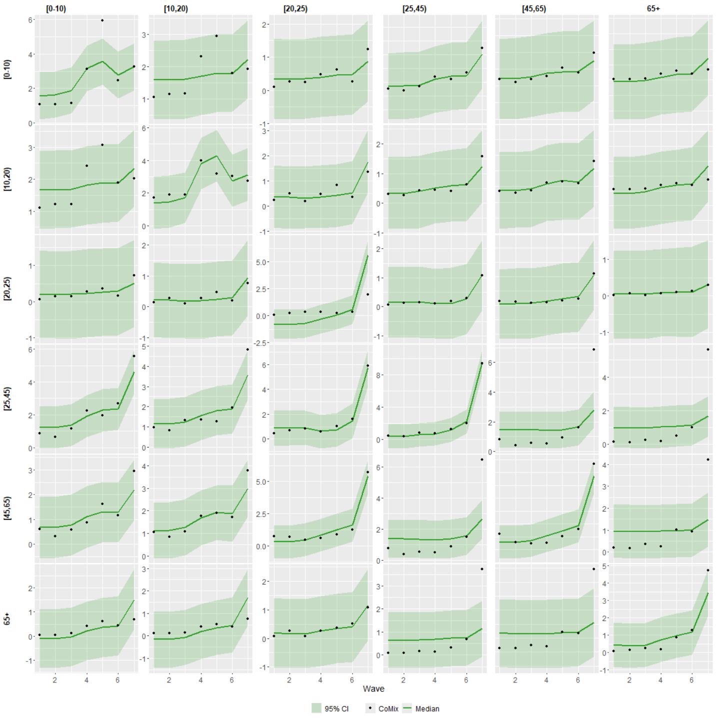


(Italy)

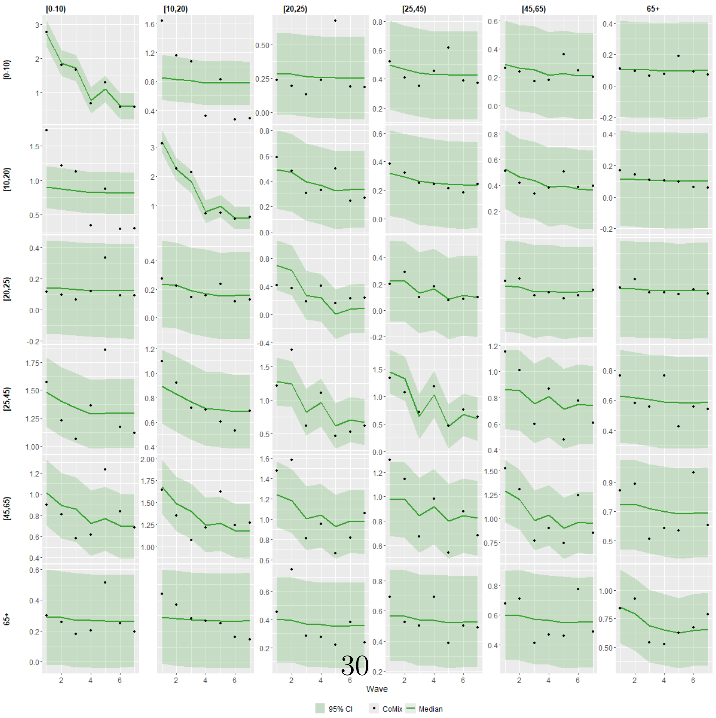


Figure S3: Estimated number of contacts between age groups, medians (lines) and 95% CI (dark shaded areas) along with reported number of contacts from CoMix(dots). (continued)

(Lithuania)

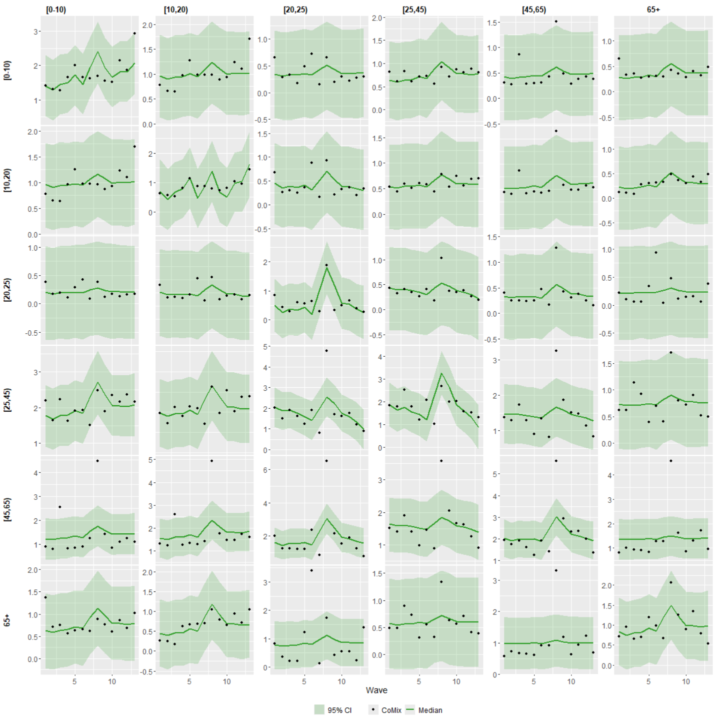


(Netherlands)

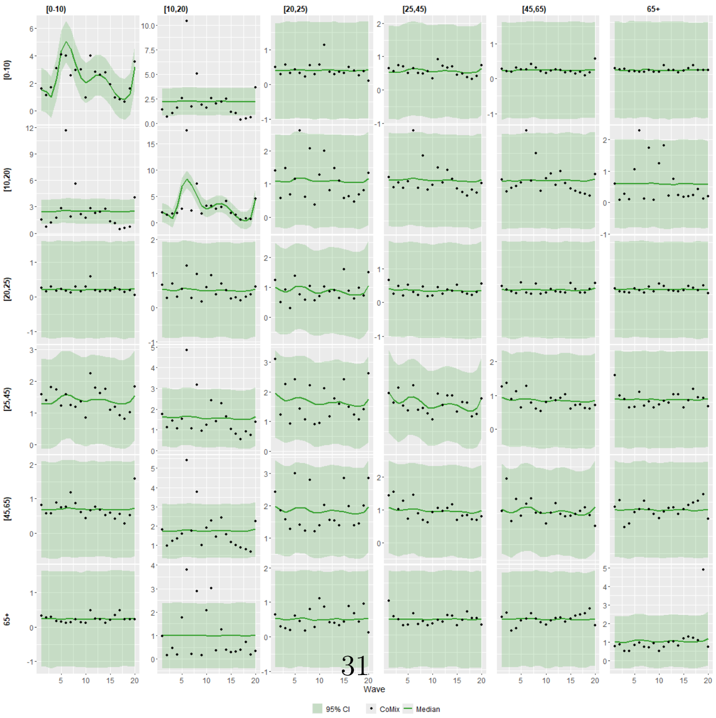




Figure S3: Estimated number of contacts between age groups, medians (lines) and 95% CI (dark shaded areas) along with reported number of contacts from CoMix(dots). (continued)

(Poland)

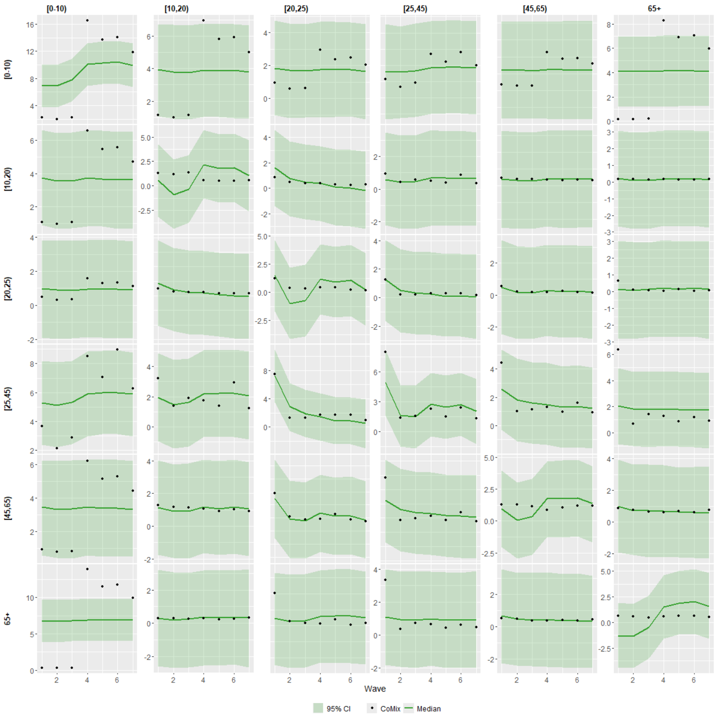


(Portugal)

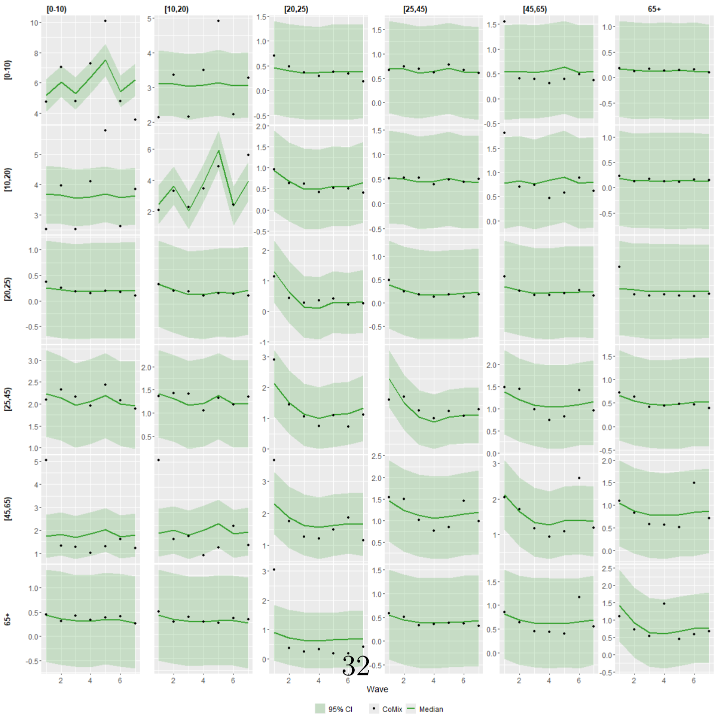




Figure S3: Estimated number of contacts between age groups, medians (lines) and 95% CI (dark shaded areas) along with reported number of contacts from CoMix(dots). (continued)

(Slovakia)

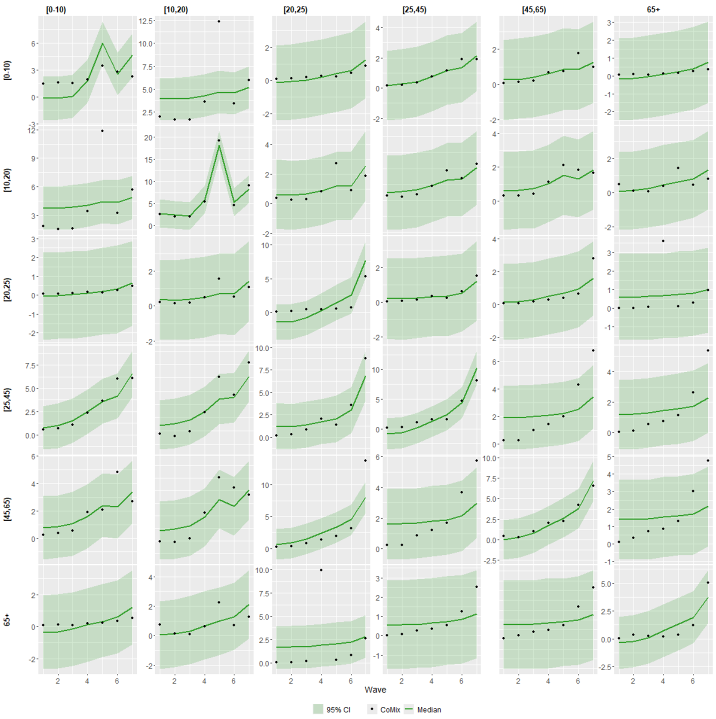


(Slovenia)

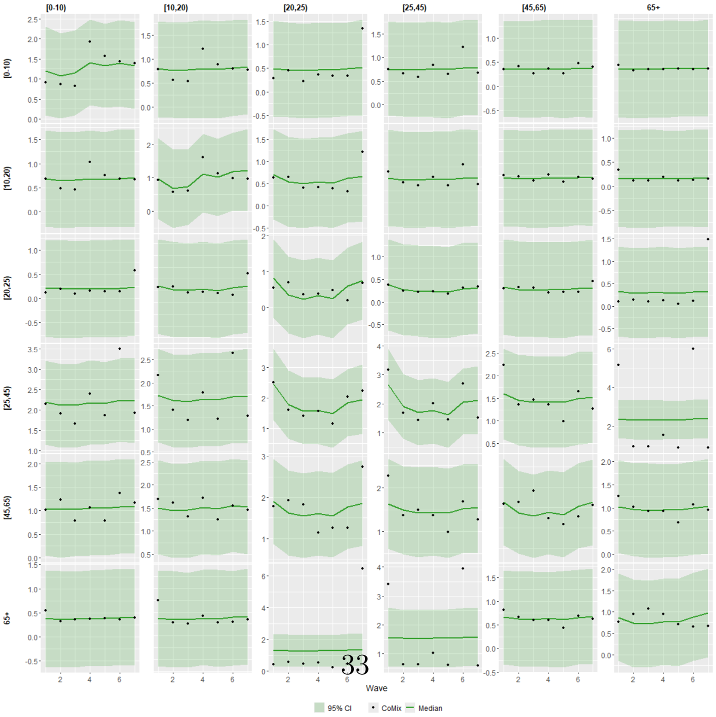




Figure S3: Estimated number of contacts between age groups, medians (lines) and 95% CI (dark shaded areas) along with reported number of contacts from CoMix(dots). (continued)

(Spain)

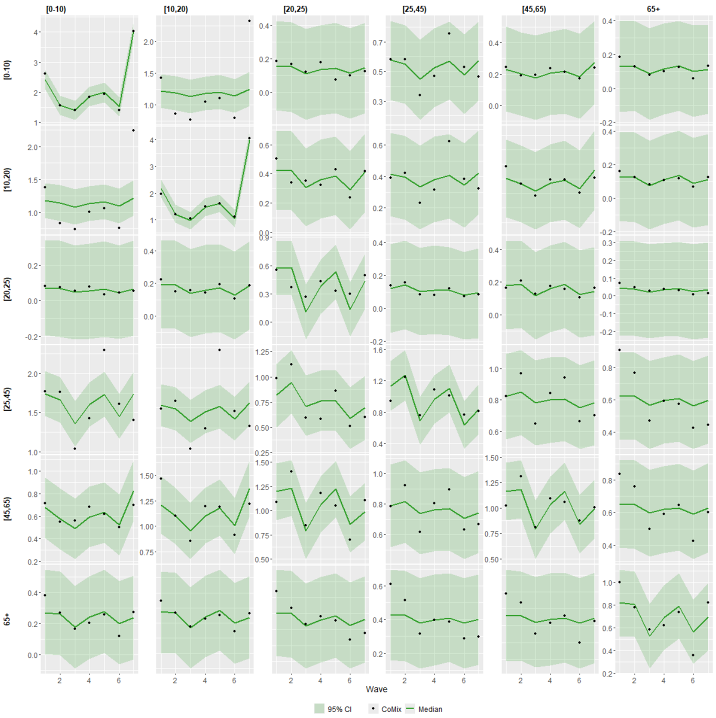


Figure S3: Estimated number of contacts between age groups, medians (lines) and 95% CI (dark shaded areas) along with reported number of contacts from CoMix(dots). (continued)

(Austria)

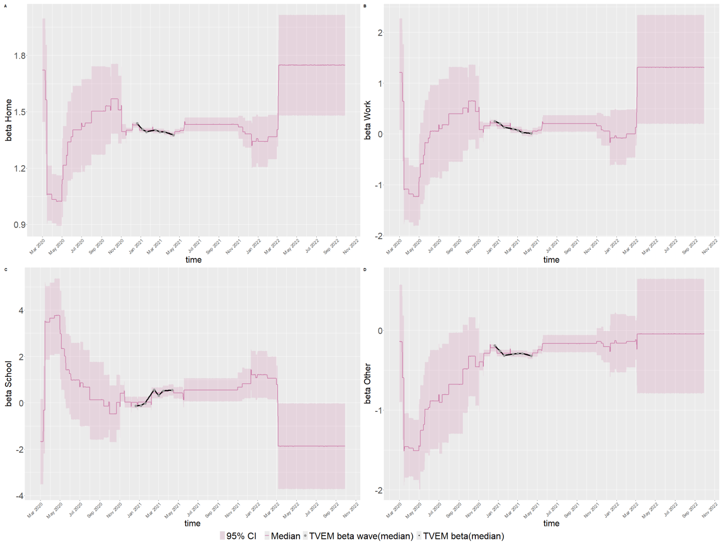


(Belgium)

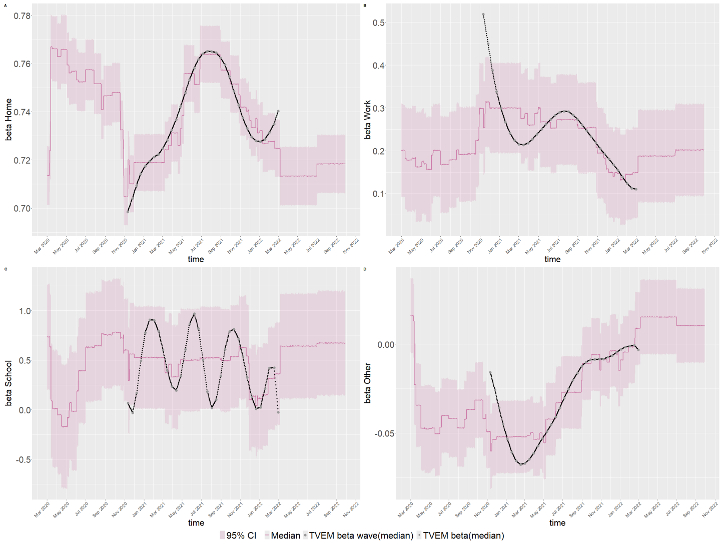


Figure S4: Predicted change in contacts during the pandemic resulting from a change at a specific location before the pandemic across time based on the NPIs.

(Croatia)

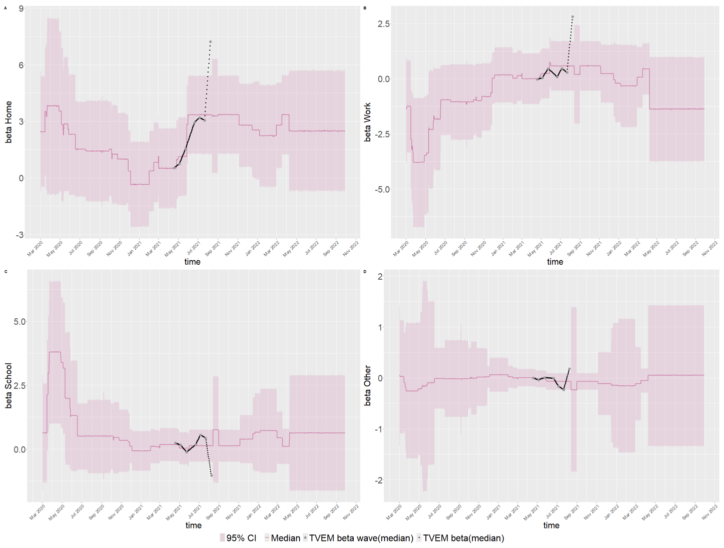


(Denmark)

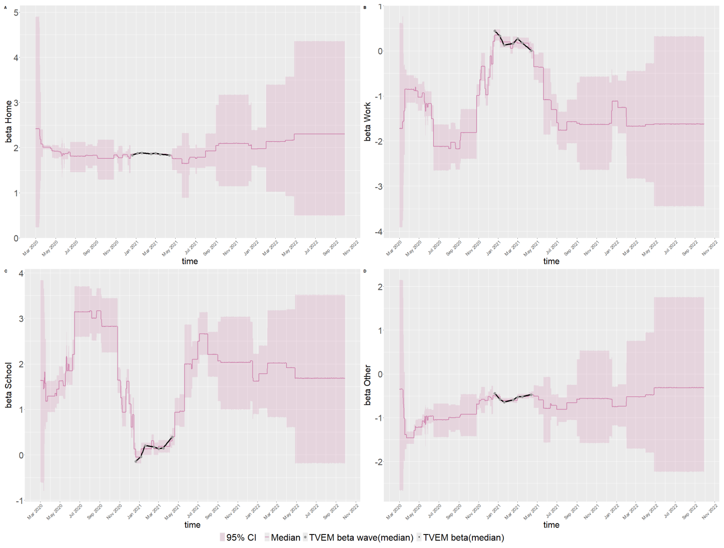


Figure S4: Predicted change in contacts during the pandemic resulting from a change at a specific location before the pandemic across time based on the NPIs. (continued)

(Estonia)

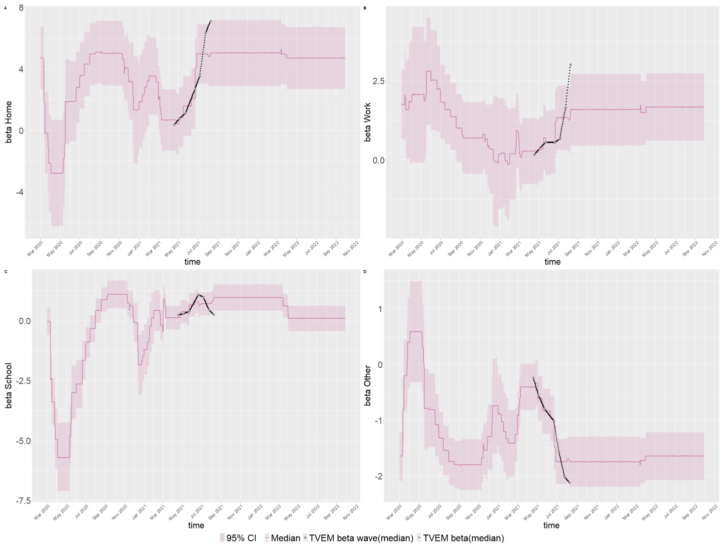


(Finland)

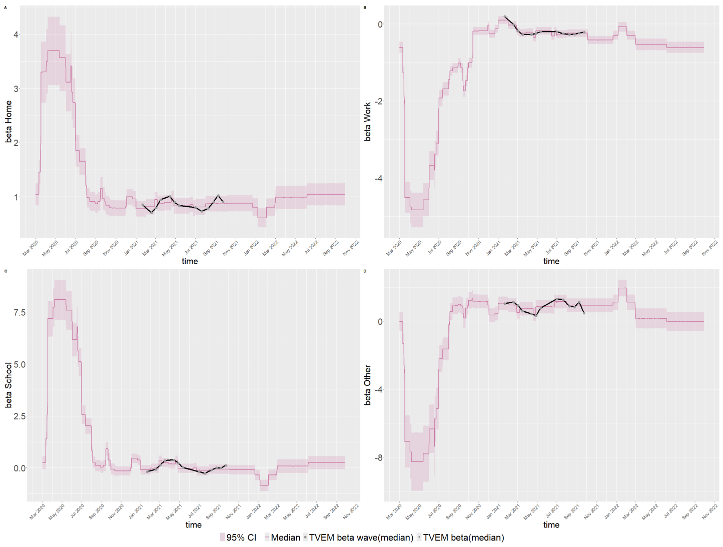


Figure S4: Predicted change in contacts during the pandemic resulting from a change at a specific location before the pandemic across time based on the NPIs. (continued)

(France)

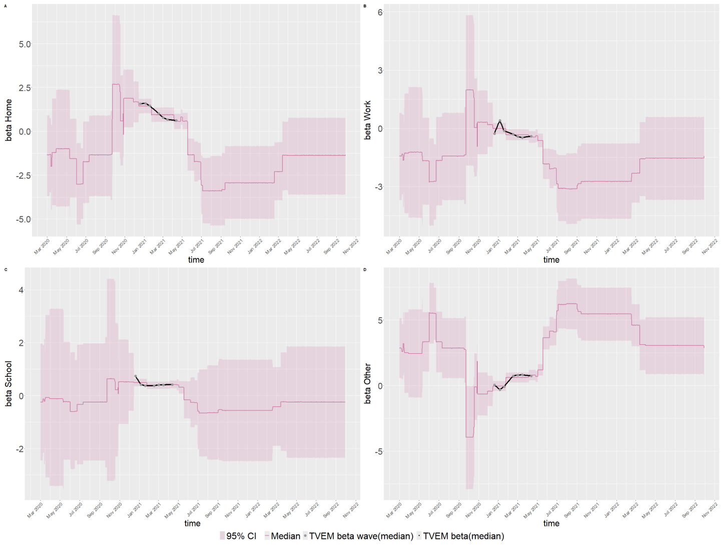


(Greece)

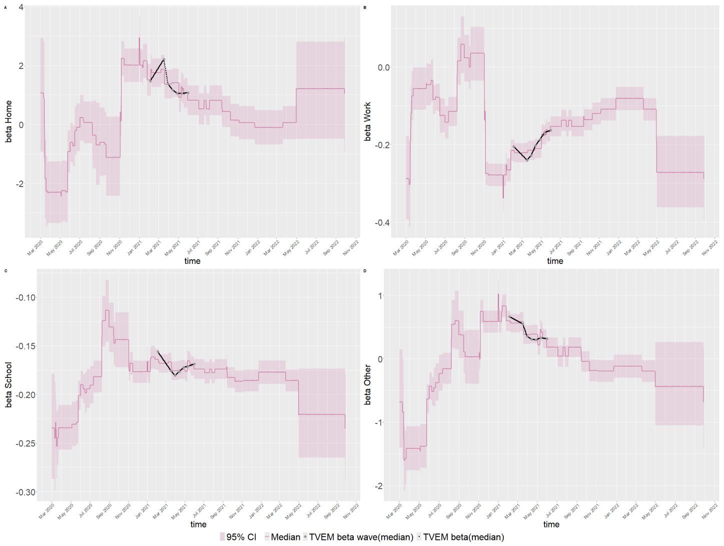


Figure S4: Predicted change in contacts during the pandemic resulting from a change at a specific location before the pandemic across time based on the NPIs. (continued)

(Hungary)

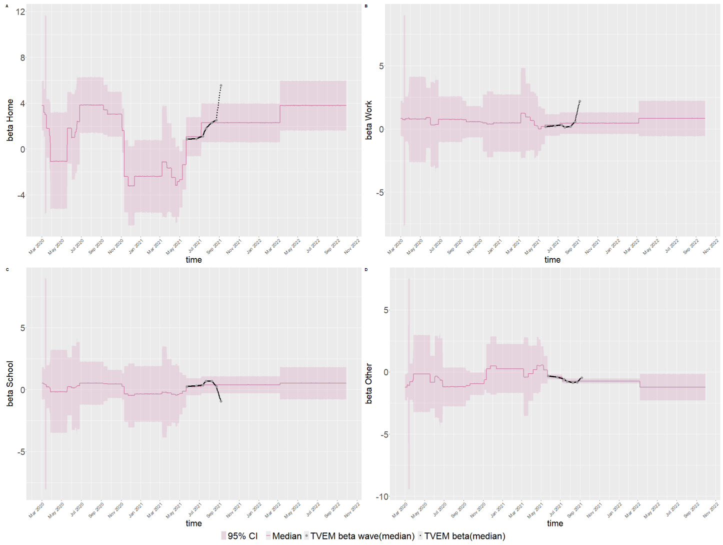


(Italy)

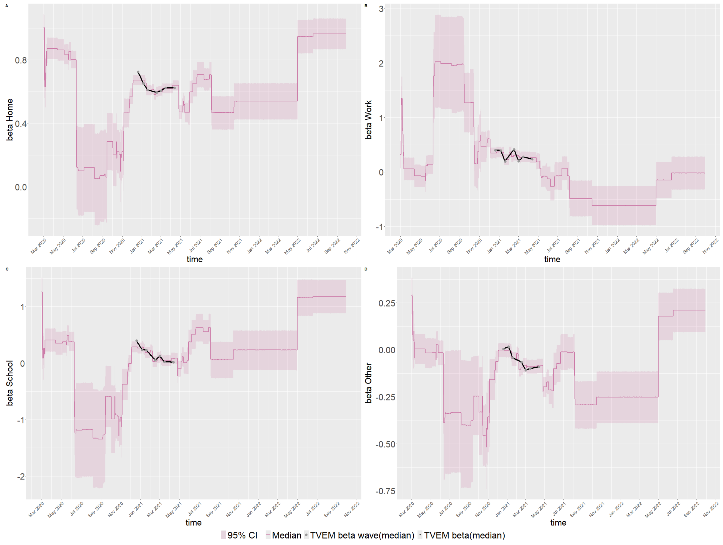


Figure S4: Predicted change in contacts during the pandemic resulting from a change at a specific location before the pandemic across time based on the NPIs. (continued)

(Lithuania)

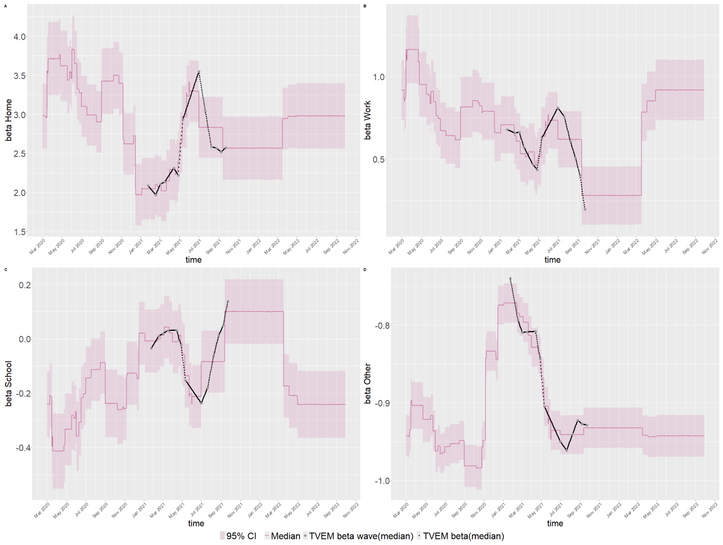


(Netherlands)

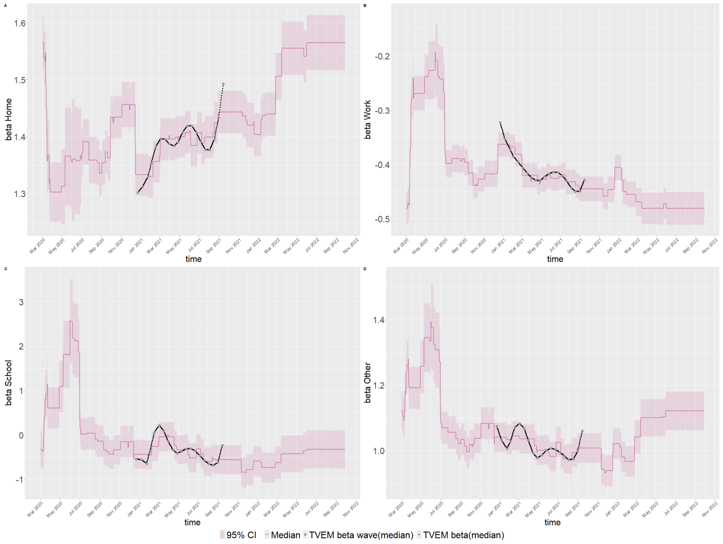


Figure S4: Predicted change in contacts during the pandemic resulting from a change at a specific location before the pandemic across time based on the NPIs. (continued)

(Poland)

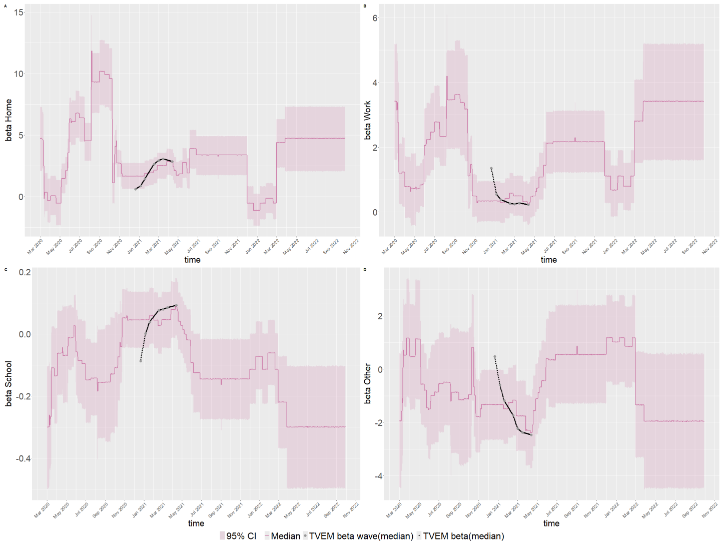


(Portugal)

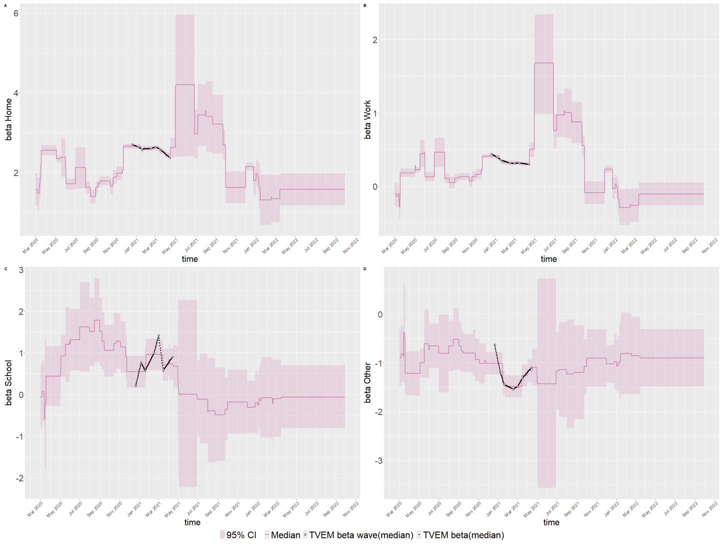


Figure S4: Predicted change in contacts during the pandemic resulting from a change at a specific location before the pandemic across time based on the NPIs. (continued)

(Slovakia)

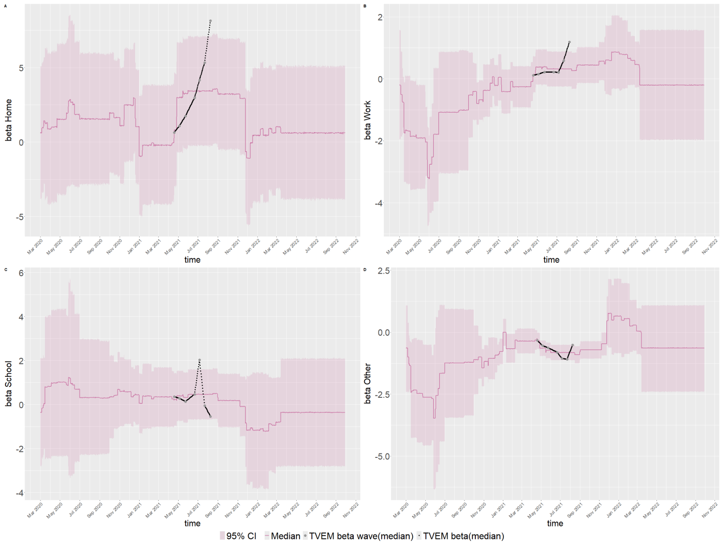


(Slovenia)

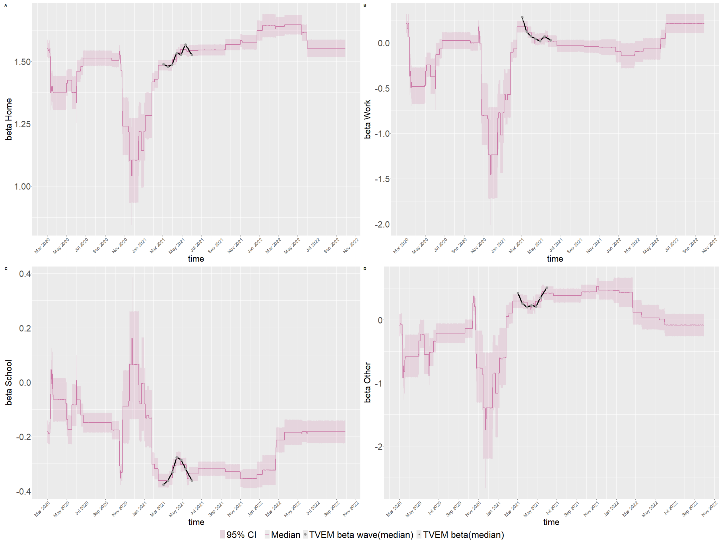


Figure S4: Predicted change in contacts during the pandemic resulting from a change at a specific location before the pandemic across time based on the NPIs. (continued)

(Spain)

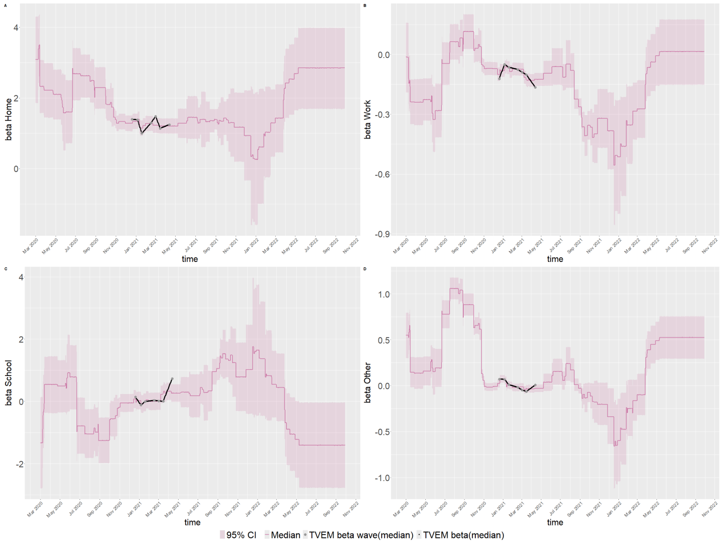


Figure S4: Predicted change in contacts during the pandemic resulting from a change at a specific location before the pandemic across time based on the NPIs. (continued)

(Austria)

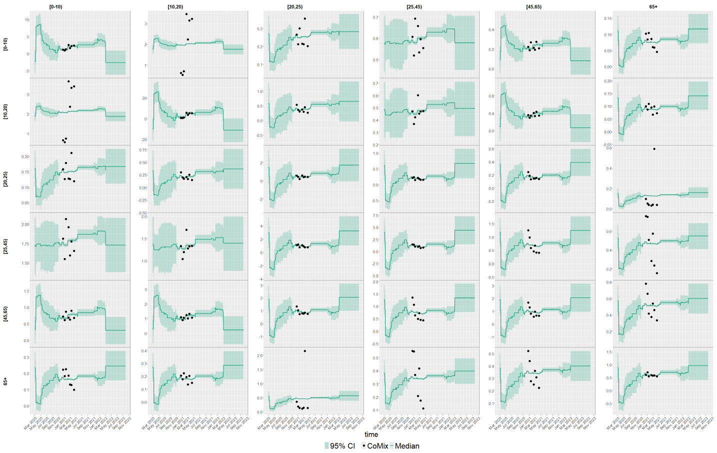


(Belgium)

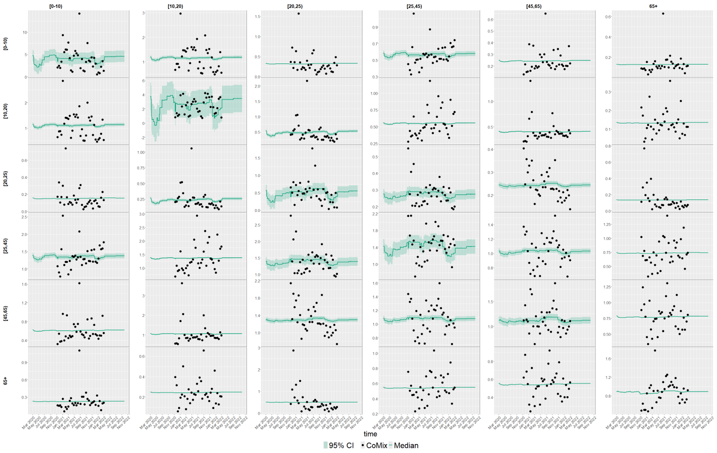


Figure S5: Predicted number of contacts between age groups, medians (lines) and 95% CI (dark shaded areas) based on posterior medians of $\beta_t$ along with reported number of contacts from CoMix(dots).

(Croatia)

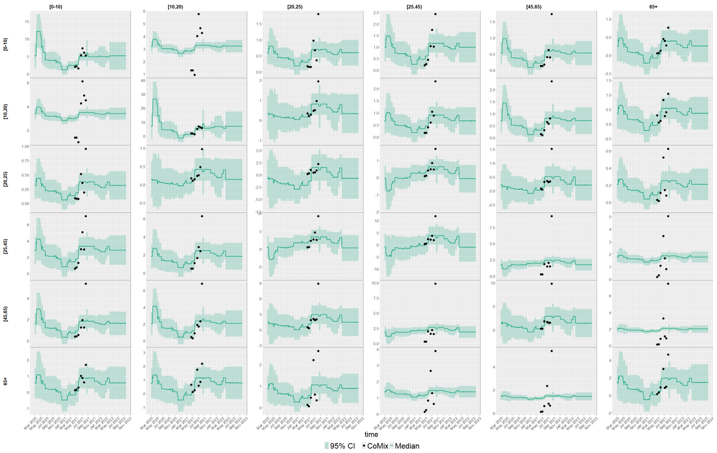


(Denmark)

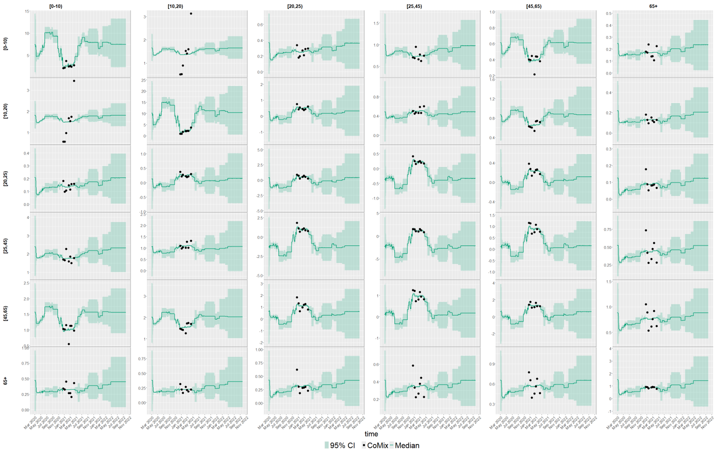


Figure S5: Predicted number of contacts between age groups, medians (lines) and 95% CI (dark shaded areas) based on posterior medians of $\beta_t$ along with reported number of contacts from CoMix(dots). (continued)

(Estonia)

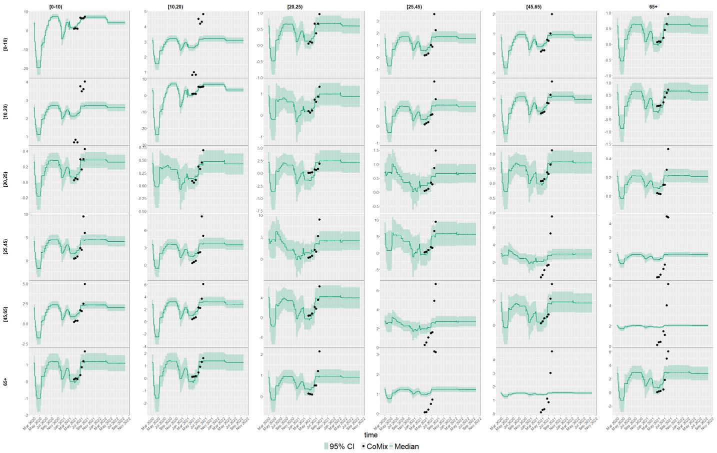


(Finland)

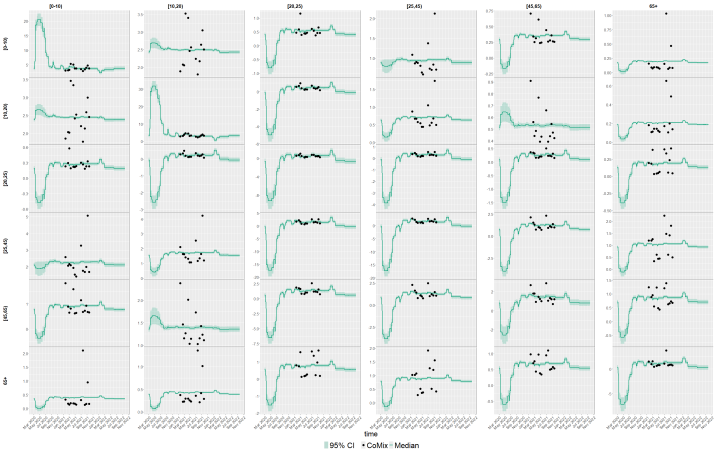


Figure S5: Predicted number of contacts between age groups, medians (lines) and 95% CI (dark shaded areas) based on posterior medians of $\beta_t$ along with reported number of contacts from CoMix(dots). (continued)

(France)

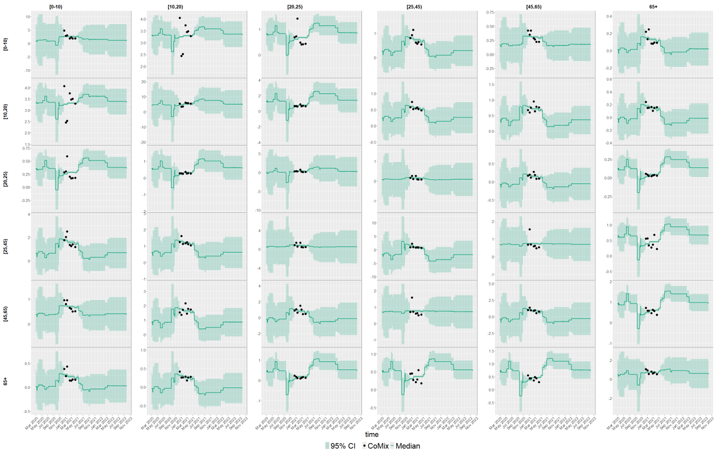


(Greece)

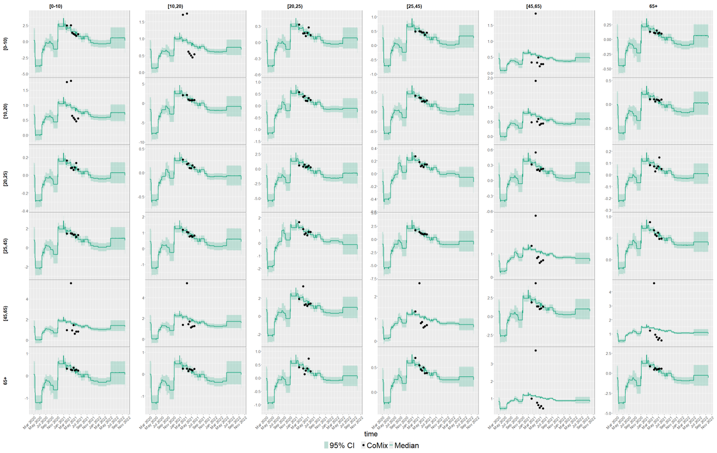


Figure S5: Predicted number of contacts between age groups, medians (lines) and 95% CI (dark shaded areas) based on posterior medians of $\beta_t$ along with reported number of contacts from CoMix(dots). (continued)

(Hungary)

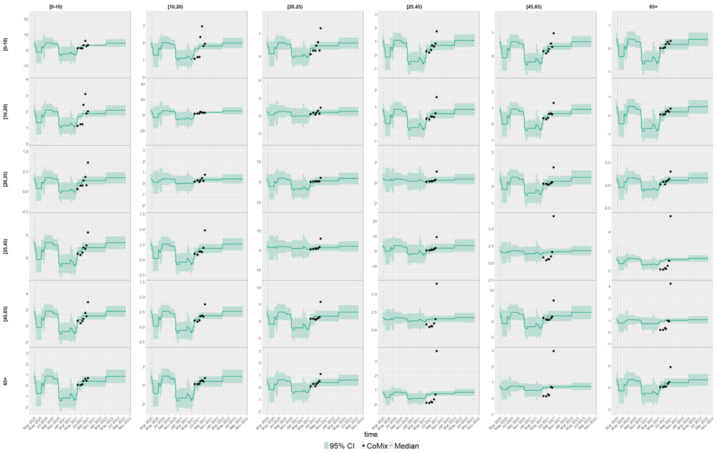

(Italy)

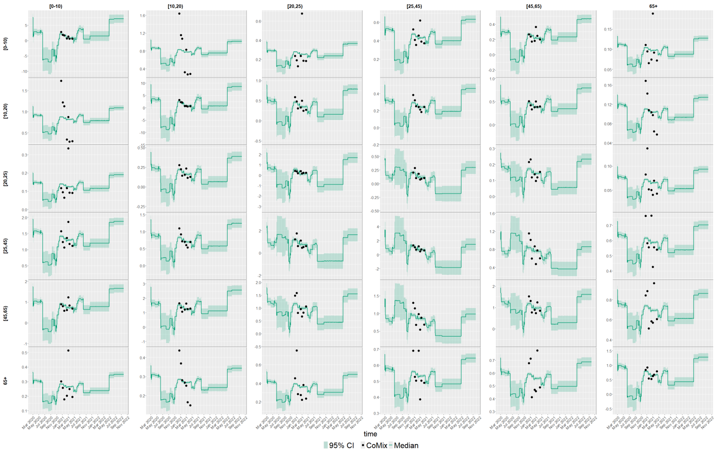

Figure S5: Predicted number of contacts between age groups, medians (lines) and 95% CI (dark shaded areas) based on posterior medians of $\beta_t$ along with reported number of contacts from CoMix(dots). (continued)

(Lithuania)

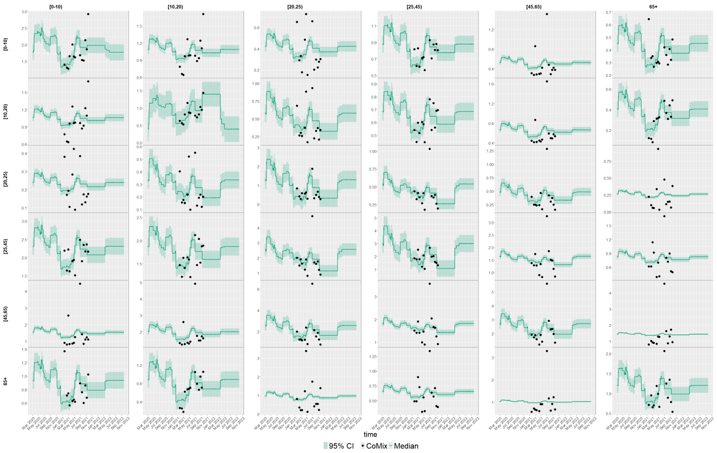


(Netherlands)

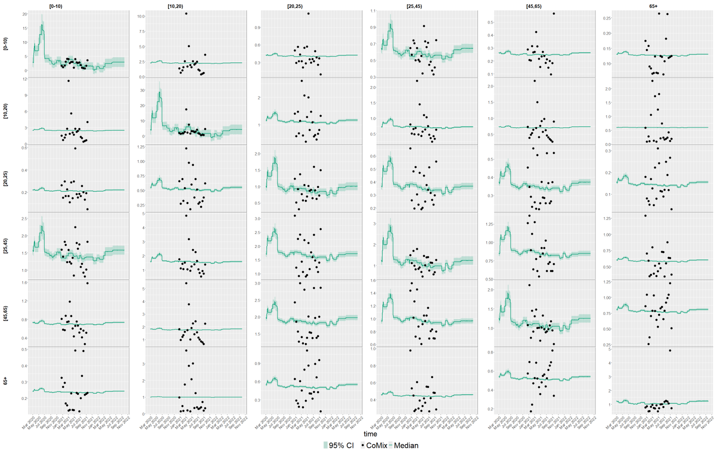


Figure S5: Predicted number of contacts between age groups, medians (lines) and 95% CI (dark shaded areas) based on posterior medians of $\beta_t$ along with reported number of contacts from CoMix(dots). (continued)

(Poland)

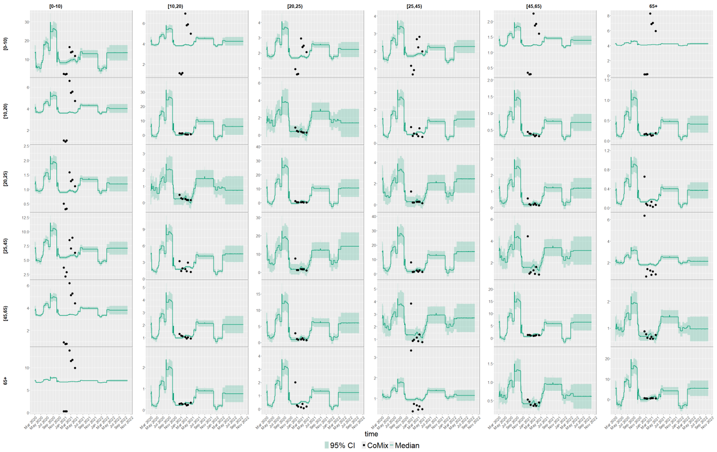


(Portugal)

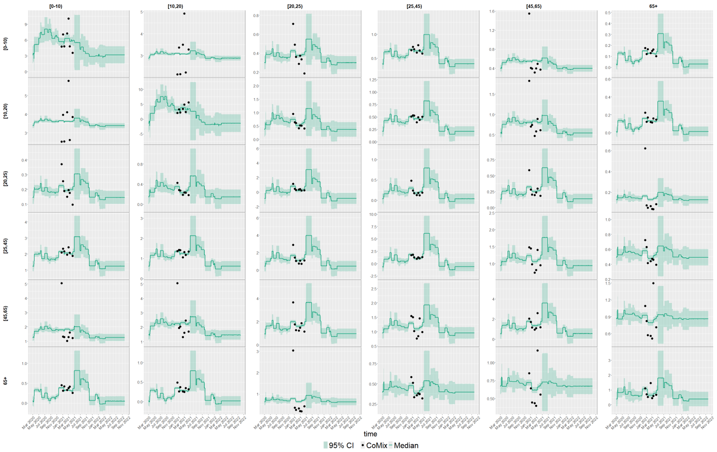


Figure S5: Predicted number of contacts between age groups, medians (lines) and 95% CI (dark shaded areas) based on posterior medians of $\beta_t$ along with reported number of contacts from CoMix(dots). (continued)

(Slovakia)

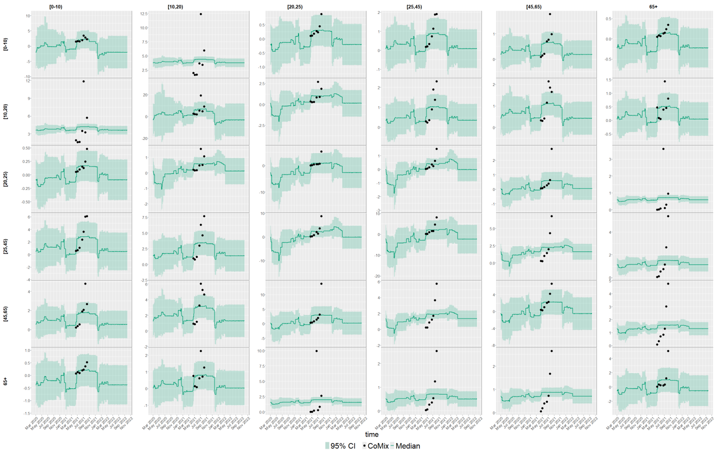


(Slovenia)

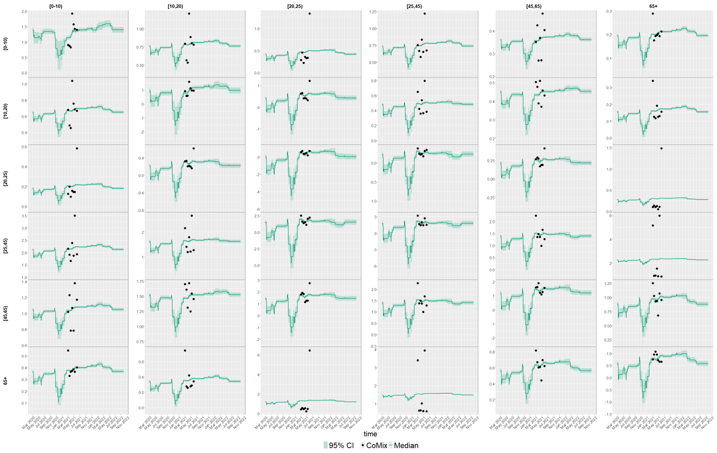


Figure S5: Predicted number of contacts between age groups, medians (lines) and 95% CI (dark shaded areas) based on posterior medians of $\beta_t$ along with reported number of contacts from CoMix(dots). (continued)

(Spain)

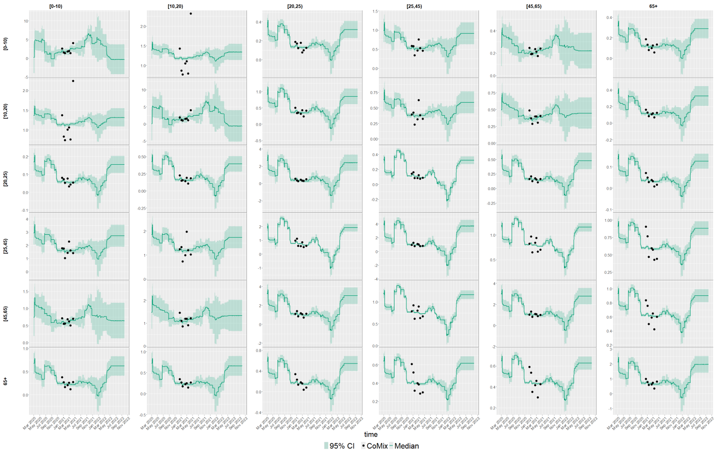


Figure S5: Predicted number of contacts between age groups, medians (lines) and 95% CI (dark shaded areas) based on posterior medians of $\beta_t$ along with reported number of contacts from CoMix(dots). (continued)

(Austria)

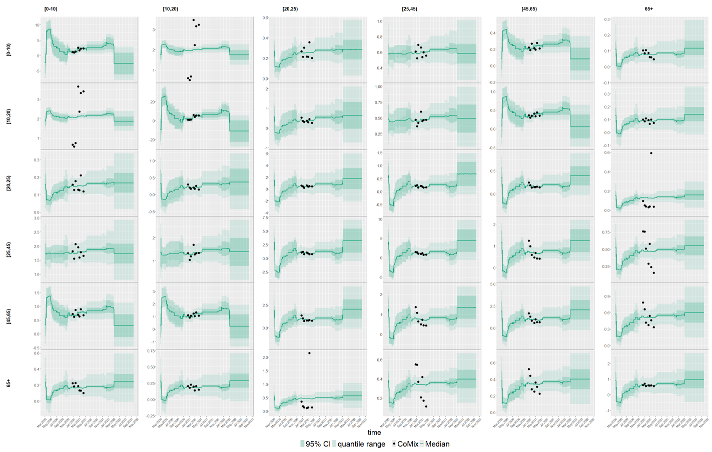


(Belgium)

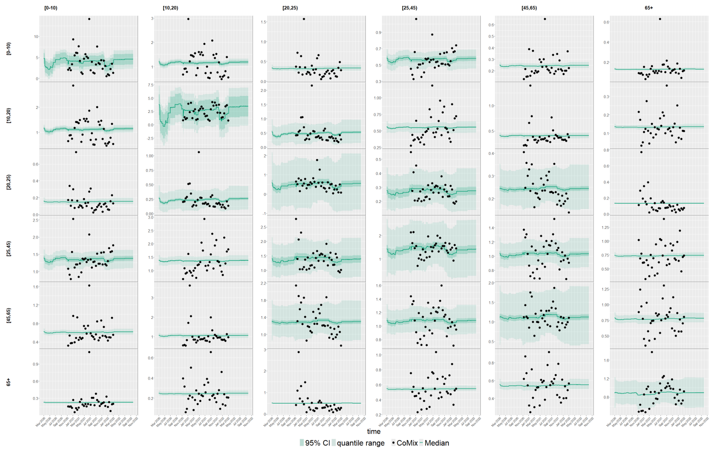


Figure S6: Predicted number of contacts between age groups, medians (lines) and 95% CI (dark shaded areas) based on 2.5%, 50%, and 97.5% percentiles of the posterior distribution of $\beta_t$ along with reported number of contacts from CoMix(dots).

(Croatia)

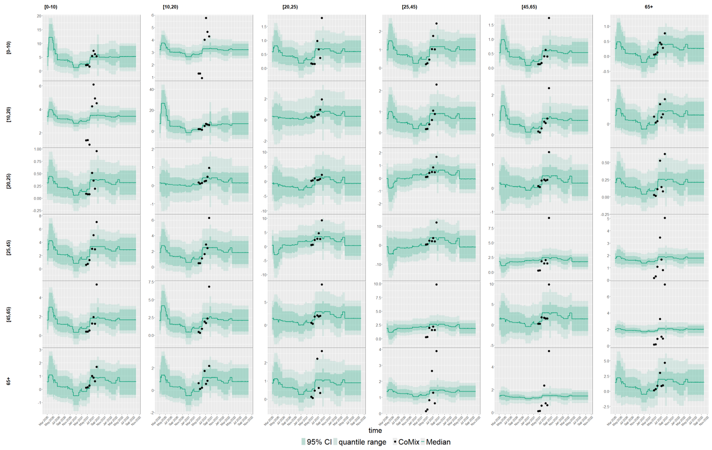


(Denmark)

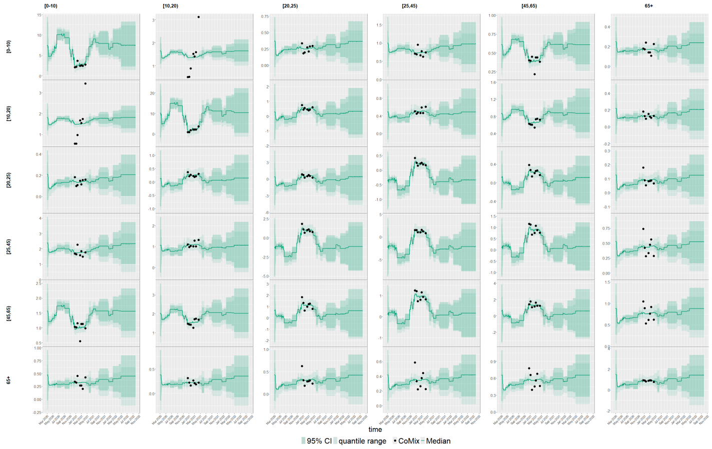


Figure S6: Predicted number of contacts between age groups, medians (lines) and 95% CI (dark shaded areas) based on 2.5%, 50%, and 97.5% percentiles of the posterior distribution of $\beta_t$ along with reported number of contacts from CoMix(dots). (continued)

(Estonia)

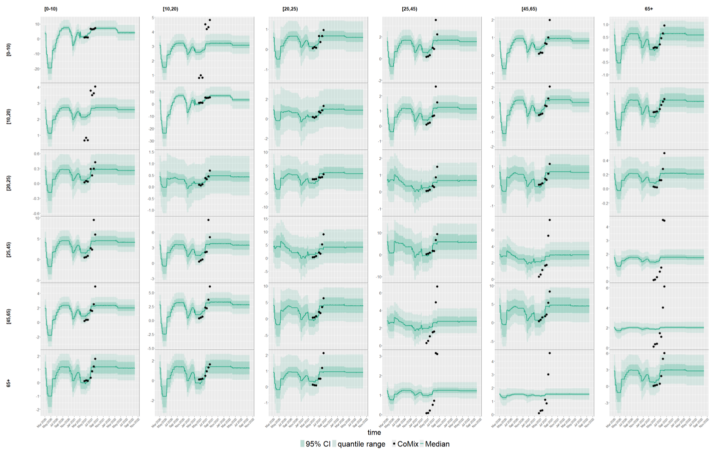


(Finland)

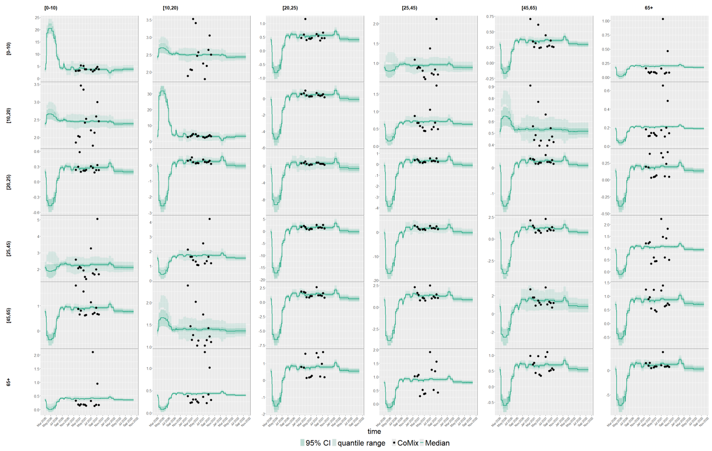


Figure S6: Predicted number of contacts between age groups, medians (lines) and 95% CI (dark shaded areas) based on 2.5%, 50%, and 97.5% percentiles of the posterior distribution of $\beta_t$ along with reported number of contacts from CoMix(dots). (continued)

(France)

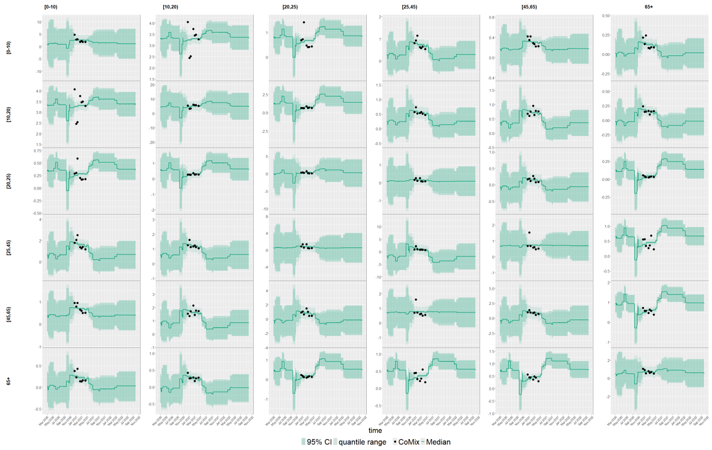

(Greece)

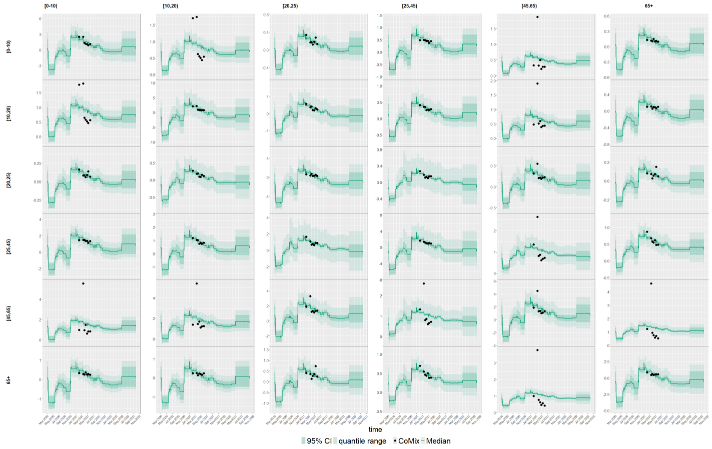

Figure S6: Predicted number of contacts between age groups, medians (lines) and 95% CI (dark shaded areas) based on 2.5%, 50%, and 97.5% percentiles of the posterior distribution of $\beta_t$ along with reported number of contacts from CoMix(dots). (continued)

(Hungary)

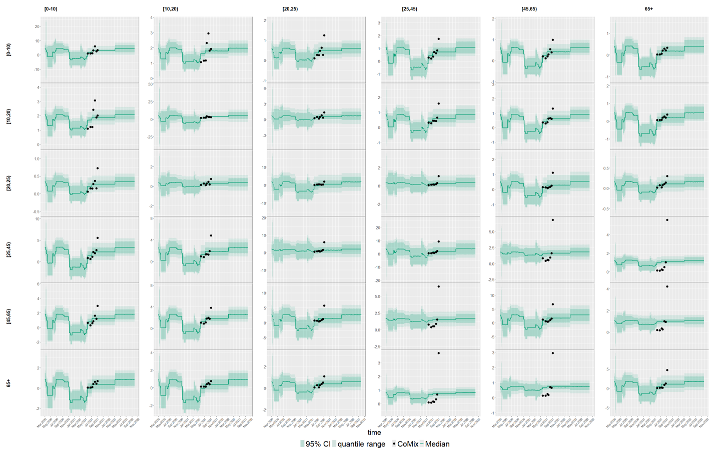


(Italy)

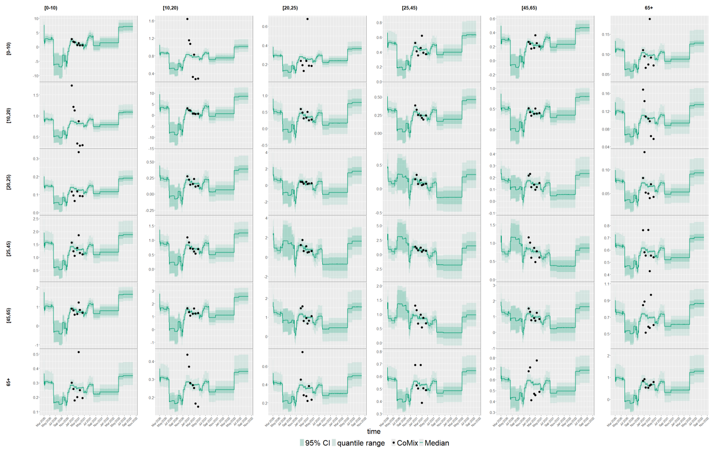


Figure S6: Predicted number of contacts between age groups, medians (lines) and 95% CI (dark shaded areas) based on 2.5%, 50%, and 97.5% percentiles of the posterior distribution of $\beta_t$ along with reported number of contacts from CoMix(dots). (continued)

(Lithuania)

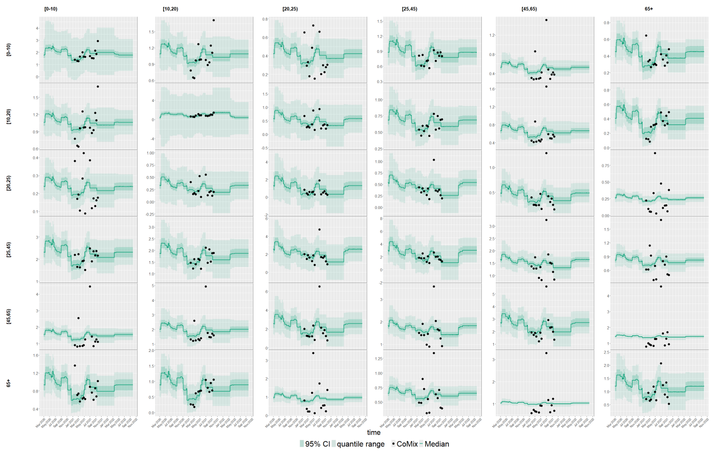


(Netherlands)

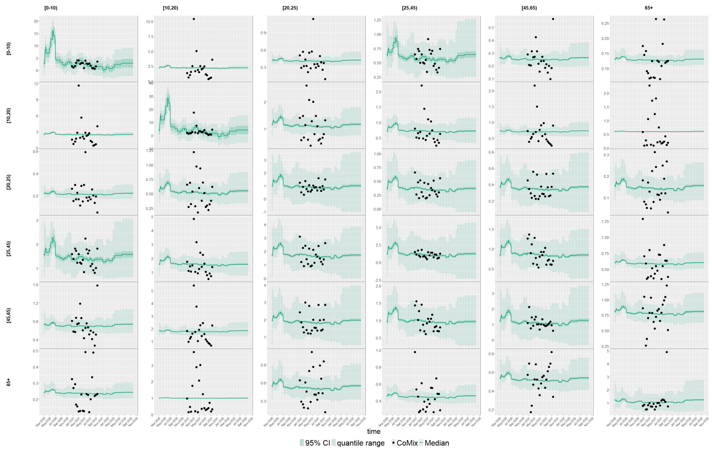


Figure S6: Predicted number of contacts between age groups, medians (lines) and 95% CI (dark shaded areas) based on 2.5%, 50%, and 97.5% percentiles of the posterior distribution of $\beta_t$ along with reported number of contacts from CoMix(dots). (continued)

(Poland)

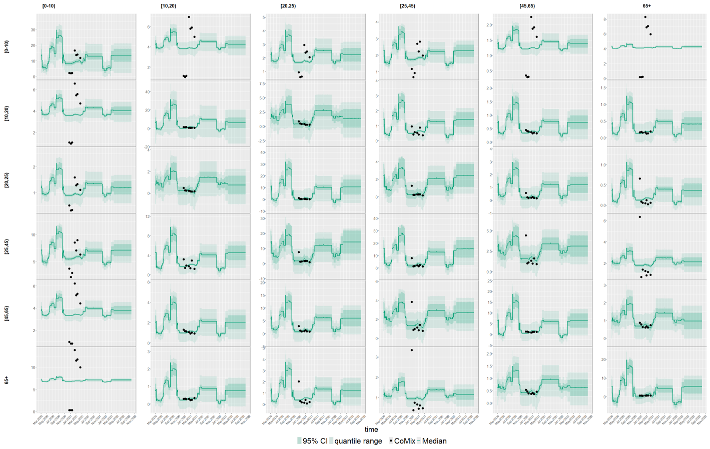


(Portugal)

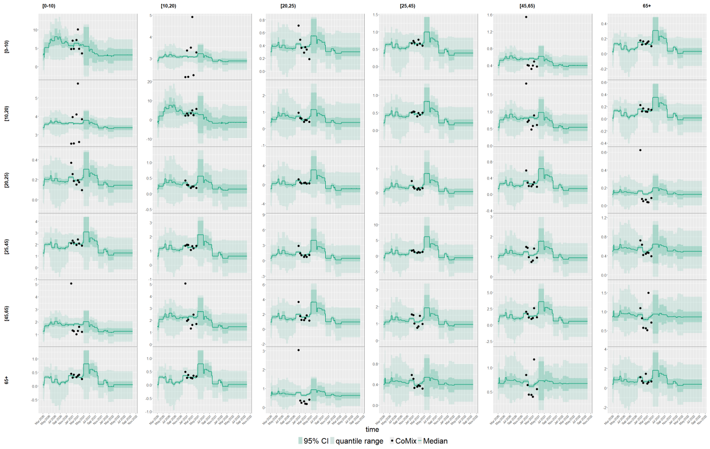


Figure S6: Predicted number of contacts between age groups, medians (lines) and 95% CI (dark shaded areas) based on 2.5%, 50%, and 97.5% percentiles of the posterior distribution of $\beta_t$ along with reported number of contacts from CoMix(dots). (continued)

(Slovakia)

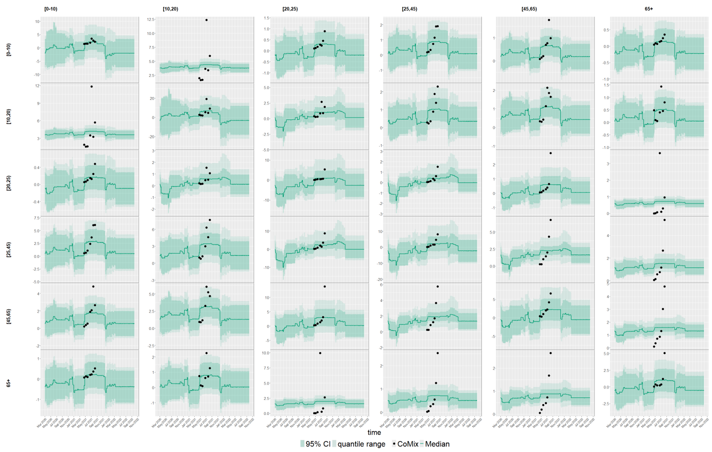


(Slovenia)

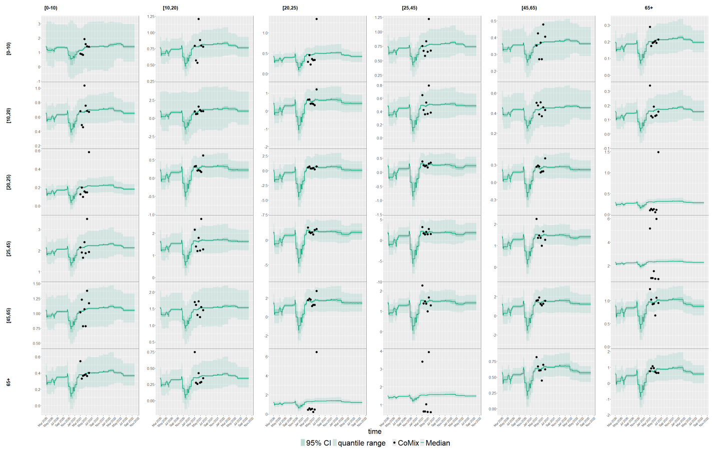


Figure S6: Predicted number of contacts between age groups, medians (lines) and 95% CI (dark shaded areas) based on 2.5%, 50%, and 97.5% percentiles of the posterior distribution of $\beta_t$ along with reported number of contacts from CoMix(dots). (continued)

(Spain)

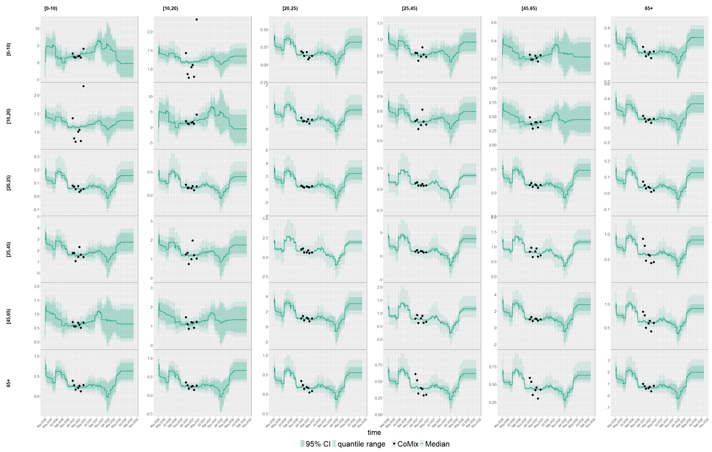


Figure S6: Predicted number of contacts between age groups, medians (lines) and 95% CI (dark shaded areas) based on 2.5%, 50%, and 97.5% percentiles of the posterior distribution of $\beta_t$ along with reported number of contacts from CoMix(dots). (continued)

(A)

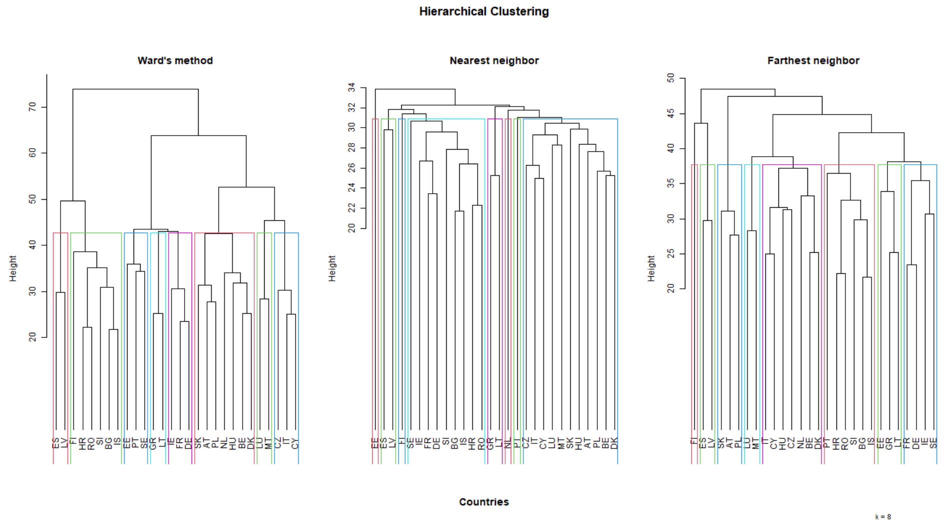


(B)

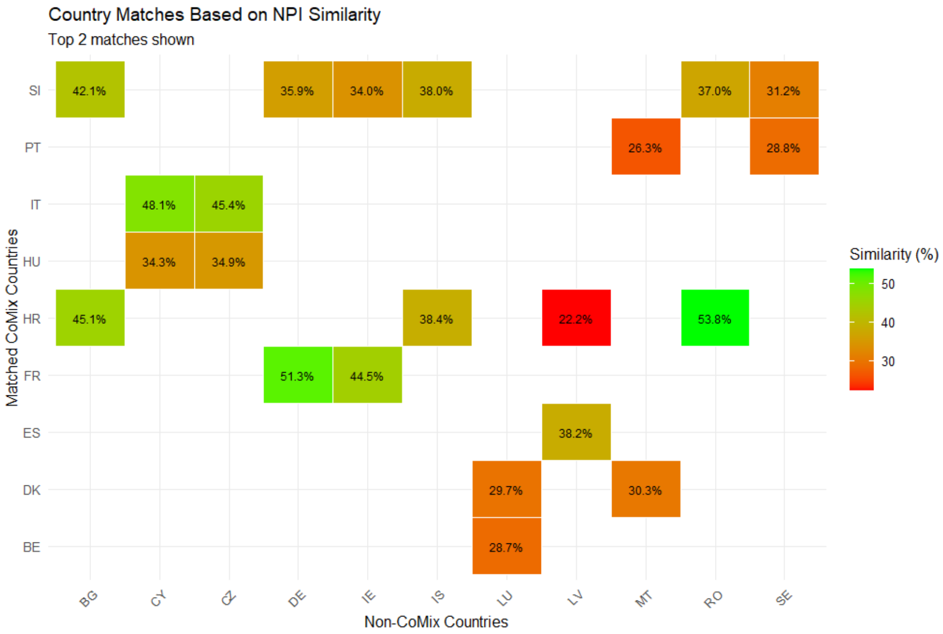


Figure S7: Hierarchical clustering (Panel (A)) and distance-based matching (Panel (B)).

(Bulgaria)

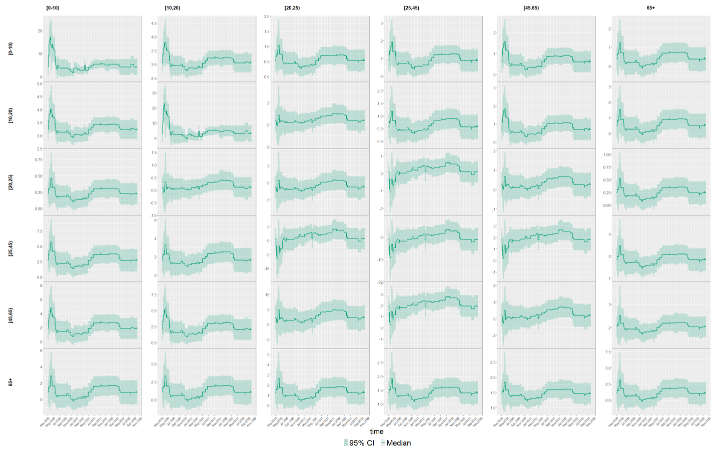


(Cyprus)

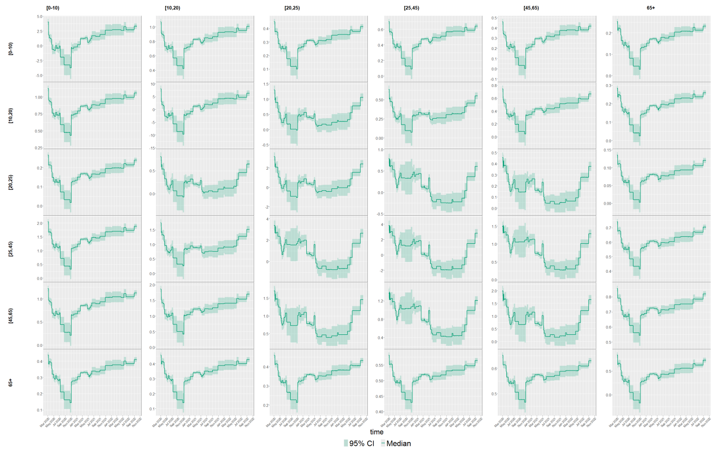


Figure S8: Predicted number of contacts between age groups, medians (lines) and 95% CI (dark shaded areas).

(Czech Republic)

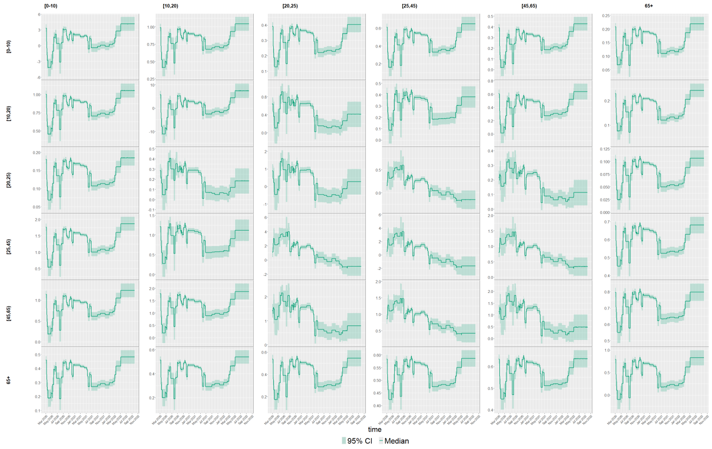


(Germany)

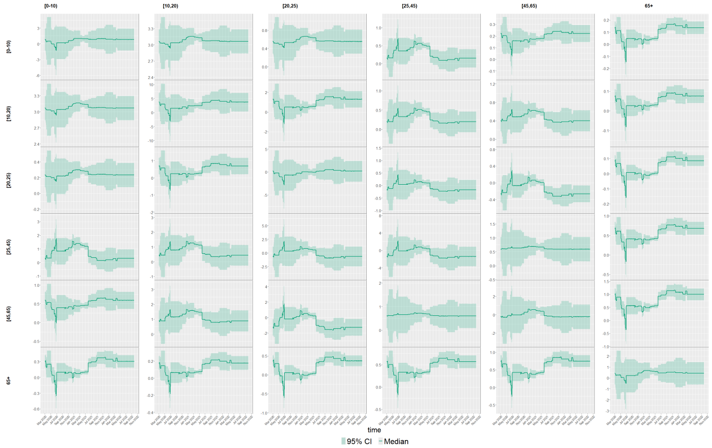


Figure S8: Predicted number of contacts between age groups, medians (lines) and 95% CI (dark shaded areas). (continued)

(Ireland)

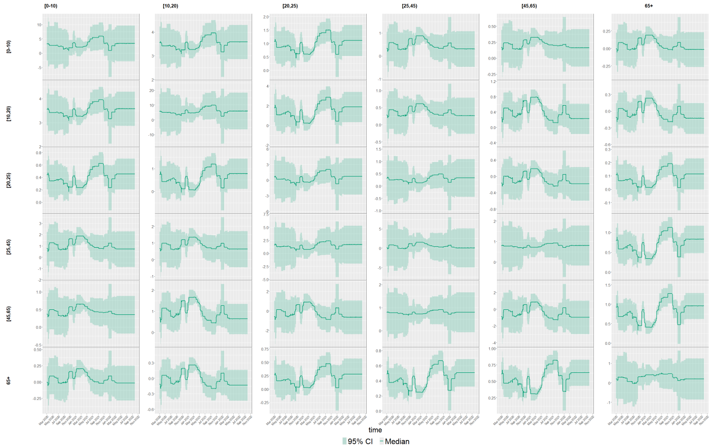

(Iceland)

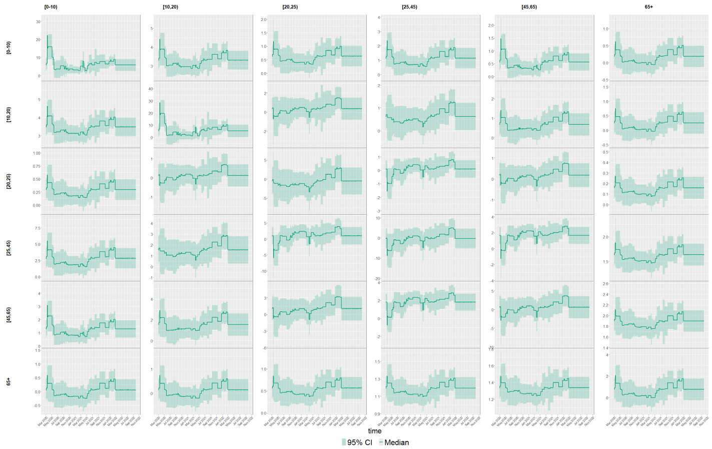

Figure S8: Predicted number of contacts between age groups, medians (lines) and 95% CI (dark shaded areas). (continued)

(Luxembourg)

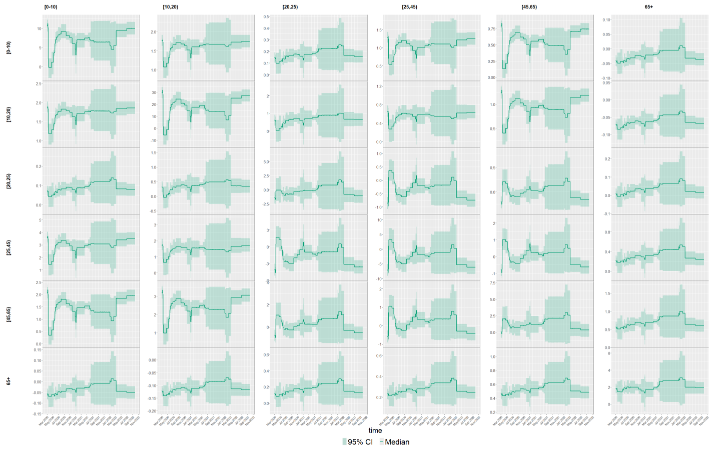


(Latvia)

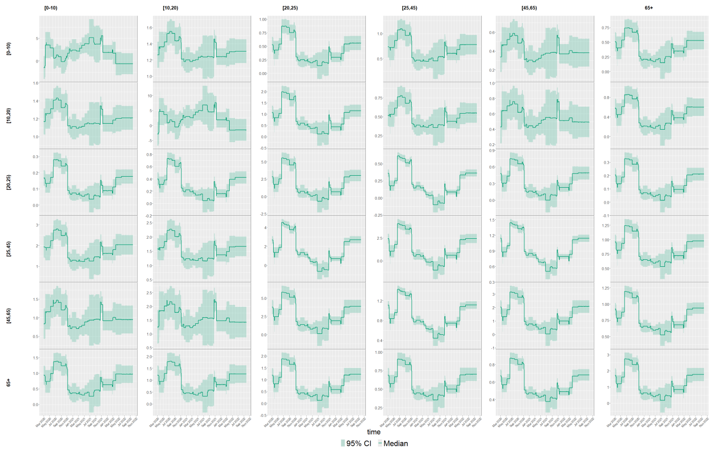


Figure S8: Predicted number of contacts between age groups, medians (lines) and 95% CI (dark shaded areas). (continued)

(Malta)

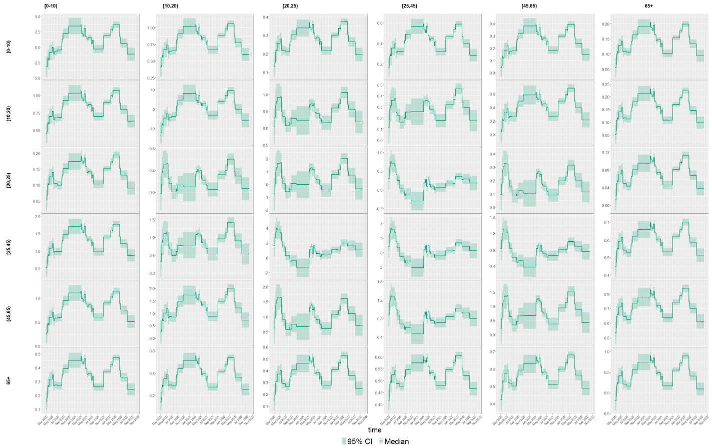


(Romania)

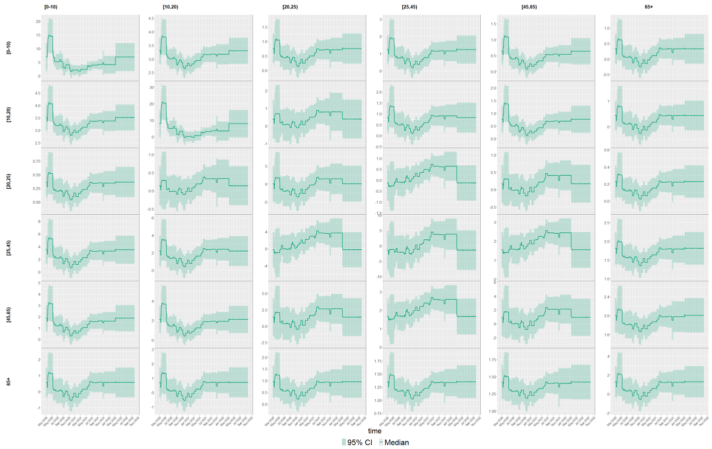


Figure S8: Predicted number of contacts between age groups, medians (lines) and 95% CI (dark shaded areas). (continued)

(Sweden)

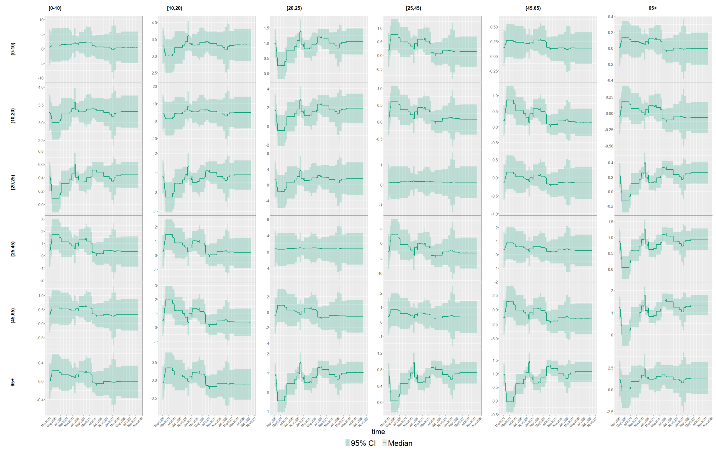

Figure S8: Predicted number of contacts between age groups, medians (lines) and 95% CI (dark shaded areas). (continued)
